# Bio-psycho-social factors’ associations with brain age: a large-scale UK Biobank diffusion study of 35,749 participants

**DOI:** 10.1101/2022.12.12.22283360

**Authors:** Max Korbmacher, Tiril P. Gurholt, Ann Marie de Lange, Dennis van der Meer, Dani Beck, Eli Eikefjord, Arvid Lundervold, Ole A. Andreassen, Lars T. Westlye, Ivan I. Maximov

## Abstract

Brain age refers to age predicted by brain features. Brain age has previously been associated with various health and disease outcomes and suggested as a potential biomarker of general health. Few previous studies have systematically assessed brain age variability derived from single and multi-shell diffusion magnetic resonance imaging data. Here, we present multivariate models of brain age derived from various diffusion approaches and how they relate to bio-psycho-social variables within the domains of sociodemographic, cognitive, life-satisfaction, as well as health and lifestyle factors in midlife to old age (*N* = 35,749, 44.6 to 82.8 years of age). Bio-psycho-social factors could uniquely explain a small proportion of the brain age variance, in a similar pattern across diffusion approaches: cognitve scores, life satisfaction, health and lifestyle factors adding to the variance explained, but not socio-demographics. Consistent brain age associations across models were found for waist-to-hip ratio, diabetes, hypertension, smoking, matrix puzzles solving, and job and health satisfaction and perception. Furthermore, we found large variability in sex and ethnicity group differences in brain age. Our results show that brain age cannot be sufficiently explained by bio-psycho-social variables alone. However, the observed associations suggest to adjust for sex, ethnicity, cognitive factors, as well as health and lifestyle factors, and to observe bio-psycho-social factor interactions’ influence on brain age in future studies.

## 1 Introduction

Developmental trajectories of brain morphology are informative signaling markers of health. For example, significant deviations from normative morphology values can signify the presence or development of disease (Marquand et al., 2019; Remiszewski et al., 2022). Based on the idea that a general normative pattern could describe brain trajectories, the concept of brain age has been introduced. Here, different brain features are used to predict individuals’ age. The difference between such predicted age and chronological age, the brain age gap (BAG), has the potential as a general health biomarker, sensitive to various neurological, neuropsychiatric, and neurodegenerative disorders (Kaufmann et al., 2019; Cole, 2020; Rokicki et al., 2021). Brain age can be derived using different imaging modalities. Structural and diffusion MRI (dMRI) have shown high prediction accuracy (e.g., Beck et al., 2022b; Chen et al., 2022; Cole, 2020; Leonardsen et al., 2022; Sone et al., 2022). Different dMRI-derived parameters allow one to describe multiple changes in WM micro-structure using various diffusion-weighted approaches. Such dMRI measures provide invaluable information about WM architecture at the micrometer scale and can be associated with macroscopic outcomes. The most popular dMRI approach, diffusion tensor imaging (DTI), is often used to describe WM organization (Basser et al., 1994). However, methodological advances and newer diffusion approaches may provide more meaningful bio-physical information (Novikov et al., 2019), thereby increasing the power of cross-validation of findings and their comparability with other clinical markers (Beck et al., 2021; Billiet et al., 2015; Kamagata et al., 2020; Wood et al., 2022).

The bio-psycho-social model (Engel, 1977) strives for a holistic perspective on medical research to understand health and disease by integrating information on biological, psychological, and social factors (e.g., Ghaemi, 2018; Wade & Halligan, 2017). Brain age can be utilized in this context as an indicator of general health (Kaufmann et al., 2019), using the different levels of observation (bio-psycho-social) to describe brain age relationships with different phenotypes. While there are some studies providing evidence for brain age associations with bio-psycho-social factors, including demographic, biomedical, lifestyle, cognitive, and behavioral factors (Beck et al., 2022b; Cole, 2020; de Lange et al., 2021; Leonardsen et al., 2022; Sone et al., 2022), it remains unclear whether brain age derived from different diffusion approaches relates differentially to sociodemographic, health, life-satisfaction, and cognitive factors (**Figure 1**), and what the qualities of such relationships are.

**Figure 1.**
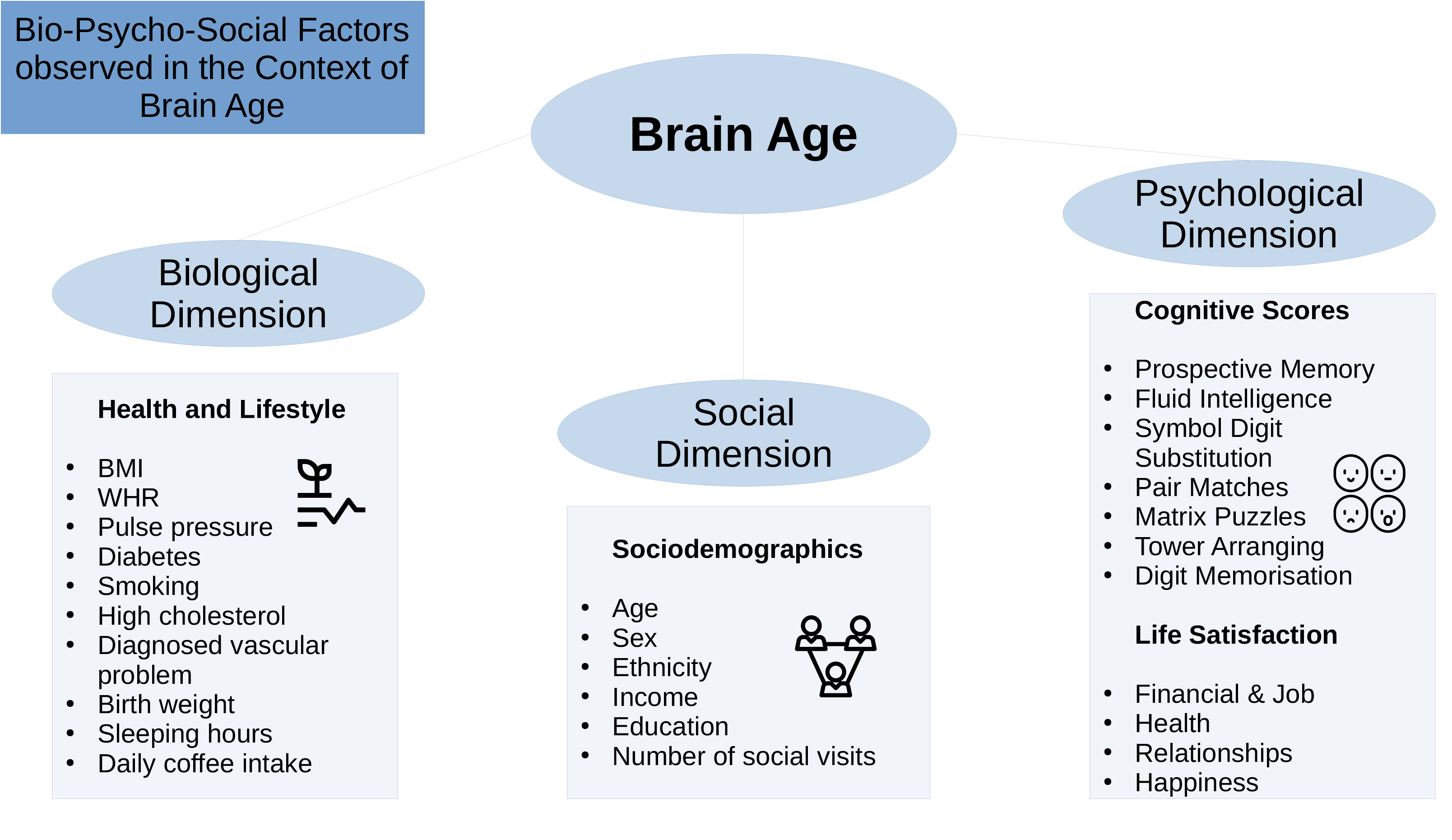
Overview of the Variables Used

It is necessary to increase our understanding and the interpretability of brain age by observing the associations of common phenotypes with brain age. There are large differences in the usage of underlying data and machine learning approaches applied to the data for brain age predictions (Franke & Gaser, 2019). Practical effects of such differences, for example, on phenotype associations, have yet to be systematized to better interpret brain age and BAG. The bio-psycho-social approach (Engel, 1977) lends itself to categorizing phenotypes into concrete groups. Within the groups, phenotype associations with brain age in general can be considered in addition to differences in underlying data used to calculate brain age. Here, we limit our investigations to dMRI-derived brain age to examine brain age relationships with bio-psycho-social factors specific to WM. WM has repeatedly been shown to change throughout ageing and to relate to different bio-psycho-social variables (Beck et al., 2021, 2022a, 2022b; Le Bihan & Iima 2015). However, phenotype-WM-brain-age relationships require still further examination. Using different diffusion approaches in this context will not only help extend commonly used diffusion tensor imaging by giving reference values to other brain age derived from other WM metrics but also provide a clearer understanding of WM-phenotype associations.

DMRI can describe various biological processes by providing markers of brain tissue changes across the lifespan (Beck et al., 2021). These markers are not only heritable (Elliott et al., 2018) but also indicative of health, for example, by being associated with psychiatric and neurological disorders, addiction, stroke (Le Bihan & Iima 2015), or cardiovascular health (Beck et al., 2022b). Various diffusion metrics that have previously been related to cognitive and mental health traits have also shared genetic underpinnings with cognitive and mental health phenotypes (Zhao et al., 2021). The biological underpinnings of dMRI markers become particularly apparent in WM abnormalities observed in severe mental disorders, including schizophrenia (Cetin-Karayumak et al., 2020) or bipolar disorder (Houenou et al., 2007). Furthermore, dMRI-derived brain age is higher in people showing accumulations of cardiometabolic risk and markers of adipose tissue distribution (Beck et al., 2022a, 2022b). Such associations between phenotypes and brain age can also be observed when comparing high to low socioeconomic status (SES) groups, where low SES individuals have lower WM integrity (Pavlakis et al., 2014; Shaked et al., 2019).

Furthermore, dMRI offers both single and multi-shell approaches and various meaningful metrics describing white matter microstructure (Fieremans et al., 2011; Jensen et al., 2005; Kaden et al., 2016a, 2016b; Reisert et al., 2017), which serve as a good basis for brain age estimations (Beck et al., 2021; Korbmacher et al., 2022) exploiting biophysically meaningful parameters of brain tissue in contrast to general measures such as grey/white volume or thickness. Differences in brain age-phenotype relationships can be expected when varying dMRI approaches, as varying underlying dMRI approaches will also produce variability in brain age predictions (see Beck et al., 2021; Korbmacher et al., 2022), potentially due to measuring different bio-physical processes (Fieremans et al., 2011; Jensen et al., 2005; Kaden et al., 2016a, 2016b; Reisert et al., 2017). These potential differences become important when attempting to generalize findings on brain age across the literature and setting standards for brain age predictions. However, to what extent age predictions based on single and multi-shell dMRI approaches relate differentially to phenotypes requires further investigation. Hence, comparing dMRI-based brain age predictions can be fruitful, not only when expanding current efforts of examining brain age associations with phenotypes but also by investigating whether differences in the underlying data can influence relationships of brain age with bio-psycho-social variables.

State-of-the-art conceptualizations of health, such as the bio-psycho-social model (Engel, 1977), recommend considering various domains or levels of explanation when assessing health outcomes, such as brain age. To date, brain age is usually calculated from a large range of MRI features. The resulting brain age estimate is then usually predicted from single variables of interest while controlling for sex and age (e.g., Cole, 2020; Leonardsen et al., 2022). However, cumulative and synergy effects can be expected to partly explain health, which has, for example, been shown for cardiometabolic risk factors explaining brain age (see Beck et al., 2022a, 2022b). Hence, we group available phenotypes that have previously been found influential for health (**Figure 1**) into health and lifestyle factors, representing the biological dimension of the bio-psycho-social model, respectively (Beck et al., 2022a, 2022b; Erhardt, 2009; Gill et al., 2021; Ning et al., 2020; Pham et al., 2021; Vidal-Pineiro et al., 2021). Life satisfaction factors and cognitive factors represent the psychological dimension, and sociodemographic factors the social dimension of the bio-psycho-social model, respectively.

Generally, explaining brain age variance is required to further our understanding of brain age and its multivariate relationship with different phenotypes influencing physiology directly or indirectly. We, therefore, extend previous work by explaining variance in brain age by combining sets of bio-psycho-social variables into domains of sociodemographic, health, life satisfaction, and cognitive factors (**Figure 1**) to assess their associations with brain age. In addition to exploring associations of bio-psycho-social variables with dMRI-based brain age, we differentiate between diffusion approaches used for brain age predictions and exame the consistency across diffusion approaches. Previous findings revealed weak associations of various phenotypes with brain age in the UK Biobank (e.g., Cole, 2020; Smith et al., 2019). Hence, we expect only small proportions of the variance in brain age to be predicted by bio-psycho-social variables. We also hypothesize that factors directly representing or impacting physiology are more predictive of brain age than those which impact physiology only indirectly. Thus, health factors are presumed to be more predictive of brain age than sociodemographic, cognitive, and life satisfaction factors. Finally, we expect some variability in these associations to be due to the underlying diffusion approach, as different WM properties are also expected to be differentially related to phenotypes. We may move brain age closer to the clinical utility by furthering our understanding of brain age.

## 2 Methods

### 2.1 Sample characteristics

The sample used has been described elsewhere (Korbmacher et al., 2022). In brief, the UK Biobank (UKB) (Sudlow et al., 2015) diffusion MRI data consisted of N = 42,208 participants. We excluded subjects who withdrew their informed consent (up to 22^nd^ of February 2022) or with an ICD-10 diagnosis from categories F, G, I, or stroke from the general health assessment (Field 42006; excluded: N = 3,521). We also excluded data that did not pass our quality control (N = 2,938) using the YTTRIUM method (Maximov et al. 2021). In brief, YTTRIUM converts diffusion scalar metric into 2D format using a structural similarity extension (Wang et al., 2004) of each scalar map to their mean image to create a 2D distribution of image and diffusion parameters. Quality check is based on 2 step clustering algorithm in order to identify subjects out of the main distribution. Our final sample consisted of 35,749 healthy adults. For an overview of demographics and the bio-psycho-social variables included in this study and their relationship with brain age.

### 2.2 MRI acquisition, diffusion post-processing, and TBSS analysis

UKB MRI data acquisition procedures are described elsewhere (Alfaro-Almagro et al., 2018; Miller et al., 2016; Sudlow et al., 2015). Briefly, single and multi-shell data were acquired at four different locations using identical scanners: 3T Siemens Skyra, with a standard 32-channel head coil and key diffusion parameters being MB = 3, R = 1, TE/TR = 92/3600 ms, PF 6/8, fat sat, b = 0 s/mm^2^ (5x + 3× phase-encoding reversed), b = 1,000 s/mm^2^ (50×), b = 2,000 s/mm^2^ (50×) (Alfaro-Almagro et al., 2018).

We obtained access to the raw diffusion data and pre-processed the data using an optimized pipeline as described by Maximov et al. (2019). The pipeline includes corrections for noise (Veraart et al., 2016), Gibbs ringing (Kellner et al., 2017), susceptibility-induced and motion distortions, and eddy current artifacts (Andersson & Sotiropoulos, 2016). Isotropic 1 mm^3^ Gaussian smoothing was carried out using FSL’s (Smith et al., 2004; Jenkins et al., 2012) *fslmaths*. Employing the multi-shell data, Diffusion Tensor Imaging (DTI), Diffusion Kurtosis Imaging (DKI) (Jensen et al., 2005) and White Matter Tract Integrity (WMTI) (Fieremans et al., 2011) metrics were estimated using Matlab 2017b code (https://github.com/NYU-DiffusionMRI/DESIGNER). Spherical mean technique SMT (Kaden et al., 2016b), and multi-compartment spherical mean technique (mcSMT) (Kaden et al., 2016a) metrics were estimated using original code (https://github.com/ekaden/smt) (Kaden et al., 2016a, 2016b). Estimates from the Bayesian Rotational Invariant Approach (BRIA) were evaluated by the original Matlab code (https://bitbucket.org/reisert/baydiff/src/master/) (Reisert et al., 2017).

Previous advances observing age-dependent WM changes have largely focused on single-shell diffusion, such as DTI with DTI-derived metrics being fractional anisotropy (FA), and axial (AD), mean (MD), and radial (RD) diffusivity, all being highly sensitive to age (Beck et al., 2021; Cox, 2016; Westlye et al., 2010). More recently developed multi-shell diffusion approaches which extend the space of derivable diffusion metrics appear more sensitive to brain changes and sex differences (Lawrence et al., 2021), and at the same time less sensitive to motion artefacts than single-shell models (Pines et al., 2020). Newer approaches are 1) BRIA, as an alternative to not rely on fiber orientation but rotation invariant feature (Reisert et al., 2017), 2) DKI, a method tackling the problem of non-Gaussian diffusion (Jensen et al., 2005); 3) WMTI, which extends DKI by calculating inter and extra-axonal features (Fieremans et al., 2011); and 4) SMT (Kaden et al., 2016b) and 5) mcSMT, which factor out neurite orientation to give a better estimate of microscopic diffusion anisotropy (Kaden et al., 2016a). The selection of diffusion models was dictated by a few practical reasons. There are two conventional approaches (DTI and DKI) describing the general WM changes. As a result, these approaches are expected to be sensitive to a broad range of aging-related effects associated with WM maturation (Westlye et al., 2010; Yap et al., 2013). Advanced dMRI approaches enable more detailed quantification associated with age in a different manner (Beck et al., 2021; Cox et al., 2016). Diffusion modelling relies on biophysically motivated assumptions such as the axon bundle distribution (WMTI) or attempts to suppress such kind of parameters (SMT and SMT mc). Another modelling option are Bayesian rotation invariants (BRIA), providing multiple measures of WM but depending on efficacy of initial Bayesian simulations. All together, these approaches allow us to indirectly verify the stability and reliability of diffusion assumptions in brain-age prediction on their own and in comparison to each other, or to determine similarity among scalar metrics appearing in several diffusion approaches.

In total, we obtained 28 metrics (**Supplementary Table 1**) from six diffusion modeling approaches (DTI, DKI, WMTI, SMT, mcSMT, and BRIA). To normalize all metrics, we used tract-based spatial statistics (TBSS) (Smith et al., 2006) as part of FSL (Smith et al., 2004; Jenkins et al., 2012). In brief, initially, all FSL BET-extracted (Smith, 2002) FA images were aligned to MNI space using non-linear transformation (FNIRT) (Jenkins et al., 2012). Subsequently, we derived the mean FA image and the related mean FA skeleton. Each diffusion scalar map was projected onto the mean FA skeleton using the standard TBSS procedure. To provide a quantitative description of diffusion metrics we evaluated averaged values over the skeleton and two WM atlases, namely the Johns Hopkins University (JHU) atlas (Mori et al., 2005) and the JHU tractography atlas (Hua et al., 2008; see **Supplementary Table 2** for an overview). Finally, we obtained 20 WM tracts and 48 regions of interest (ROIs) based on a probabilistic WM atlas (JHU) (Hua et al., 2008) for each of the 28 metrics, including the mean skeleton values. Altogether, we derived 1,932 features per individual (28 metrics * (48 ROIs + 1 skeleton mean + 20 tracts)); see **Supplementary Table 1** for metrics, **Supplementary Table 2** for regions and tracts.

### 2.3 Brain Age Predictions

We computed brain age predictions derived from 8 different models including the six diffusion approaches, their whole-brain average scores (mean multimodal), and a model combining the six diffusion approaches and their whole-brian average scores (full multimodal). Each of the six diffusion approaches details WM features based on differing modelling assumptions and were assumed to provide unique brain age scores. Whole-brain average scores for each of the six diffusion approaches’metrics were investigated on their own to further our understanding of spacial specificity. Finally, previous results (Beck et al., 2021, 2022b; de Lange et al., 2020b) provide clear evidence of strong age prediction performance when combining diffusion metrics. We hence included a model combining all diffusion approaches’ metrics and their whole-brain average scores to compare whether there are differences in multimodal to single diffusion approaches’ brain-age-phenotype associations.

Brain age was predicted using the XGBoost algorithm implemented in Python (v3.7.1), being a highly effective algorithm for tabular data (Chen & Guestrin, 2016). From the total included sample (N = 35,749), we used 10% (N = 3,575) for hyperparameter tuning on a data set containing data from all diffusion approaches (i.e., full multimodal data with 1,932 features/parameters) using 5-fold cross-validation (after estimating an optimal hyperparameter tuning set size (Korbmacher et al., 2022)). The considered hyperparameters for the randomized grid search were learning rate with a range of 0.01 to 0.3 and steps of 0.05, maximum layers/depth with a range of 3 to 6 and steps of 1, and number of trees with a range of 100 to 1000 and steps of 50. The resulting hyperparameters (learning rate = 0.05, max layers/depth = 3, and the number of trees = 750) were then used in a 10-fold cross-validation applied to the test set (N = 32,174). Cross-validaton was used to leverage the full sample size and to calculate the uncertainty around the estimates (for such see Korbmacher et al., 2022). The cross-validation procedure was executed using each of the six diffusion approaches’ metrics, whole-brain averaged metrics for all approaches (mean multimodal model), and finally a combination of all approaches and the whole-brain average scores (full multimodal model), resulting in eight brain age models (see **Supplementary Table 1** for dMRI approach-specific metrics). Each of these brain ages were used in the analyses. See **Supplementary Figure 1** for an overview of the brain age models and the following modelling of these predictions from the bio-psycho-social models.

### 2.4 Statistical Analyses

All statistical analyses were carried out using Python, version 3.7.1 and R, version 4.2.0 (www.r-project.org/) using test data set (N = 32,174). These analyses focused on the associations between brain age and (1) demographics, (2) social factors, (3) cognitive test scores, (4) life satisfaction, and (5) health and lifestyle factors (with weight on cardiometabolic factors). For detailed information on how variables were extracted and coded see **Supplementary Table 3**. First, we examined bio-psycho-social factors’ principal components associations with brain age. Second, we examined to which extend multivariate models explain brain age from the factors of the five bio-psycho-social domains. Finally, we tested whether our findings would be influenced by analyzing data separately for males and females, and present bi-variate relationships between multimodal brain age and single bio-psycho-social variables.

For bi-variate relationships between bio-psycho-social factors and full multimodal brain age, we adjusted *p*-values for multiple comparison using Bonferroni correction, dividing the alpha-level by twenty-five (α/25), the number of bi-variate associations observed. For multivariate relationships we divided alpha by eight (α/8), the number of brain age models used. Furthermore, the coefficient of determination describing the proportion of variance explained (R^2^) will be presented as marginal R^2^, referring to variance explained by fixed effects, and conditional R^2^, referring to both fixed and random effects variance explained.

#### 2.4.1 Bio-psycho-social models explaining brain age

We used linear mixed effects models with the random intercepts at the level of scanner site to explain changes in brain age from socio-demographics, cognitive test scores, life satisfaction (self-assessment), and health and lifestyle factors. The presented models were used in two different ways: first, with the principal component of the model-specific bio-psycho-social factors replacing the respective bio-psycho-social factors, and second using all eight brain ages from the different diffusion approaches on with the models. For an overview of the predictors in the multivariate bio-psycho-social models used see **Table 1**.

**Table 1:** Overview of the predictors used in the bio-psycho-social models

| Model | Predictors |
| --- | --- |
| Model 1.<br>Baseline | Age<br>Sex<br>Scanner Site* |
| Model 2.<br>Socio-demographics | Age<br>Sex<br>Scanner Site*<br>Ethnicity<br>Income<br>Education<br>Nb of Social Visits |
| Model 3.<br>Cognitive<br>Scores | Age<br>Sex<br>Scanner Site*<br>Prospective Memory<br>Fluid Intelligence<br>Symbol Digit Substitution<br>Pair Matches<br>Matrix Puzzles<br>Tower Arranging<br>Digit Memorization |
| Model 4. Life<br>Satisfaction | Age<br>Sex<br>Scanner Site*<br>Financial & Job Satisfaction<br>Friend & Family Relation Satisfaction<br>Happiness |
| Model 5.<br>Health and<br>Lifestyle | Age<br>Sex<br>Scanner Site*<br>BMI<br>WHR<br>Pulse Pressure<br>Diabetes<br>Smoking<br>High Cholesterol<br>Diagnosed Vascular Disorder<br>Birth Weight<br>Sleeping Hours<br>Daily Coffee Intake<br>Alcohol Drinker |

\* Random effect

##### We established the following models to compare

1. A baseline model capturing the relationship of age, sex, the age-sex interaction, and scanner site with brain age. This baseline model was selected as predicted age is expected to be largely reflected by chronological age. However, also sex (e.g., Rokicki et al., 2021), and scanner site (here, Bristol, Cheadle, Newcastle, Reading) and prediction bias (e.g., Jirsaraie et al., 2022) have been shown to be influential for brain age. Using a baseline model and additional models for comparison had the goal to estimate added variance explained by the bio-psycho-social models above and beyond the baseline mode (Bollen, 1989). Additionally, predictors within these bio-psycho-social models were observed. Model comparison to a baseline model (instead of a null model) is important in this context as brain age is sensitive to age, sex and scanner site (de Lange & Cole, 2020; Jirsaraie et al., 2022; Rokicki et al., 2021). Hence, instead of using a null model which does not contain much information, we used the following model as a reference point for further model comparison:

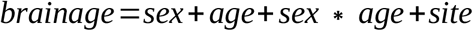
2. A sociodemographic model additionally included ethnic ancestry (binary yes/no self-reported white European; for additional information sample groupings by ethnicity see **Supplementary Table 4**), average annual total household income before tax (coded as continuous variable 1-5, with low <£18,000 to high income >£100,000), and higher education (binary yes/no self-report of having obtained higher education) relative to the baseline model.

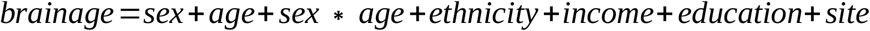
3. A cognitive model testing how non-verbal cognitive abilities add to the baseline model (overview: Fawns-Ritchie & Deary, 2020). Namely, the number of matrix puzzles solved (matrixS) testing non-verbal reasoning using COGNITO Matrices, tower arranging correctly solved (towerS) testing executive function using the Delis-Kaplan Executive Function System Tower Test, prospective memory (memory) assessed with the Rivermead Behavioural Memory Test, fluid intelligence (intel) from the UKB own Fluid IQ test, digits remembered (digits) from the Symbol Digit Modalities Tests, and the mean number of incorrect pair matches (IPM) across trail A and B assessing visual declarative memory using the Wechsler Memory Scale IV Designs I and Designs II. Correlations were small to moderate (*r*_*max*_ = 0.41) with the variance inflation factor (VIF) indicating low levels of multicollinearity (**Supplementary Figure 2**).

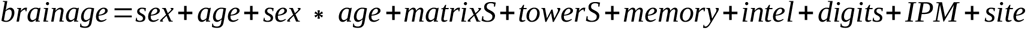
4. A life satisfaction model that additionally included job satisfaction (jobS), financial satisfaction (financeS), overall health rating (healthR), health satisfaction (healthS), family relation satisfaction (famS), happiness, friend relationship satisfaction (friendS) relative to the baseline model. Some of the model features were highly correlated (*r*_*max*_ = 0.65), yet VIF values indicated low levels of multicollinearity (**Supplementary Figure 3**).

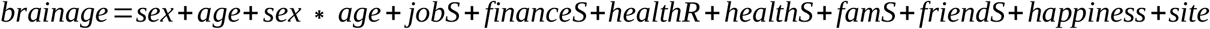
5. A health and lifestyle model testing how body mass index (BMI), pulse pressure (Ppressure: the difference between systolic and diastolic blood pressure), waist-to-hip-ratio (WHR), binary smoking status, binary diabetes diagnosis (both type I and II), binary high cholesterol (chol), binary diagnosed vascular problem (DVP), birth weight (Bweight), sleeping hours, and daily coffee intake (coffee) add to the baseline model, with only BMI and WHR showing a moderate correlation *r* = 0.42, but all other correlations being small *rs* < 0.16, with VIF values indicating only low levels of multicollinearity (**Supplementary Figure 4**).

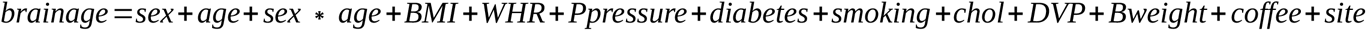

#### 2.4.2 Follow-up and quality control analyses and single bio-psycho-social factor associations with multimodal brain age

Previous research showed sex differences in brain age, suggesting sex separate analyses (Rokicki et al., 2021). Hence, we conducted the analyses described in 2.4.1 separately for males and females.

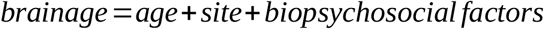

To examine the contributions of single bio-psycho-social variables to explaining WM brain age, linear mixed models were used to observe bio-psycho-social variable associations with brain age when controlling for age and sex with scanner site as a random factor. In other words, different from 2.4.1, we applied one model per bio-psycho-social factor. For simplicity, this analysis step only considered the best brain age predictions from the multimodal model including the metrics of all diffusion approaches (Korbmacher et al., 2022).

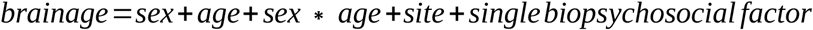

Each model was then compared to a model not including the respective bio psycho social variable:

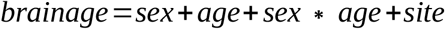

## 3 Results

### 3.1 Linear mixed effect models explaining brain age gap from bio-psycho-social factors

We ran the proposed five baseline and bio-psycho-social models with the first principal component (PC) of the numeric predictors from each of the models showing a small proportion of the variance in brain age uniquely explained by the principal components (R^2^ < 1%; **Supplementary Table 5**), with differences between these models and respective baseline models yet being highly significant (**Supplementary Table 6**).

When including bio-psycho-social factors instead of their PCs and comparing baseline to models 2-5, a larger proportion of both marginal or conditional variance in brain age could be uniquely explained by bio-psycho-social variables (marginal and conditional R^2^ < .03; **Figure 3, Supplementary Table 7**). Model comparisons showed that, with the exception of socio-demographic factors, bio-psycho-social models explained significantly more variance in brain age than the baseline model (with age, sex, and age-by-sex interaction as fixed and scanner site as random effect), irrespective of the diffusion approach used to calculate brain age (*ps* < .01; **Figure 2**). Differences between this uniquely explained variance were small across diffusion approaches (**Figure 3**).

**Figure 2.**
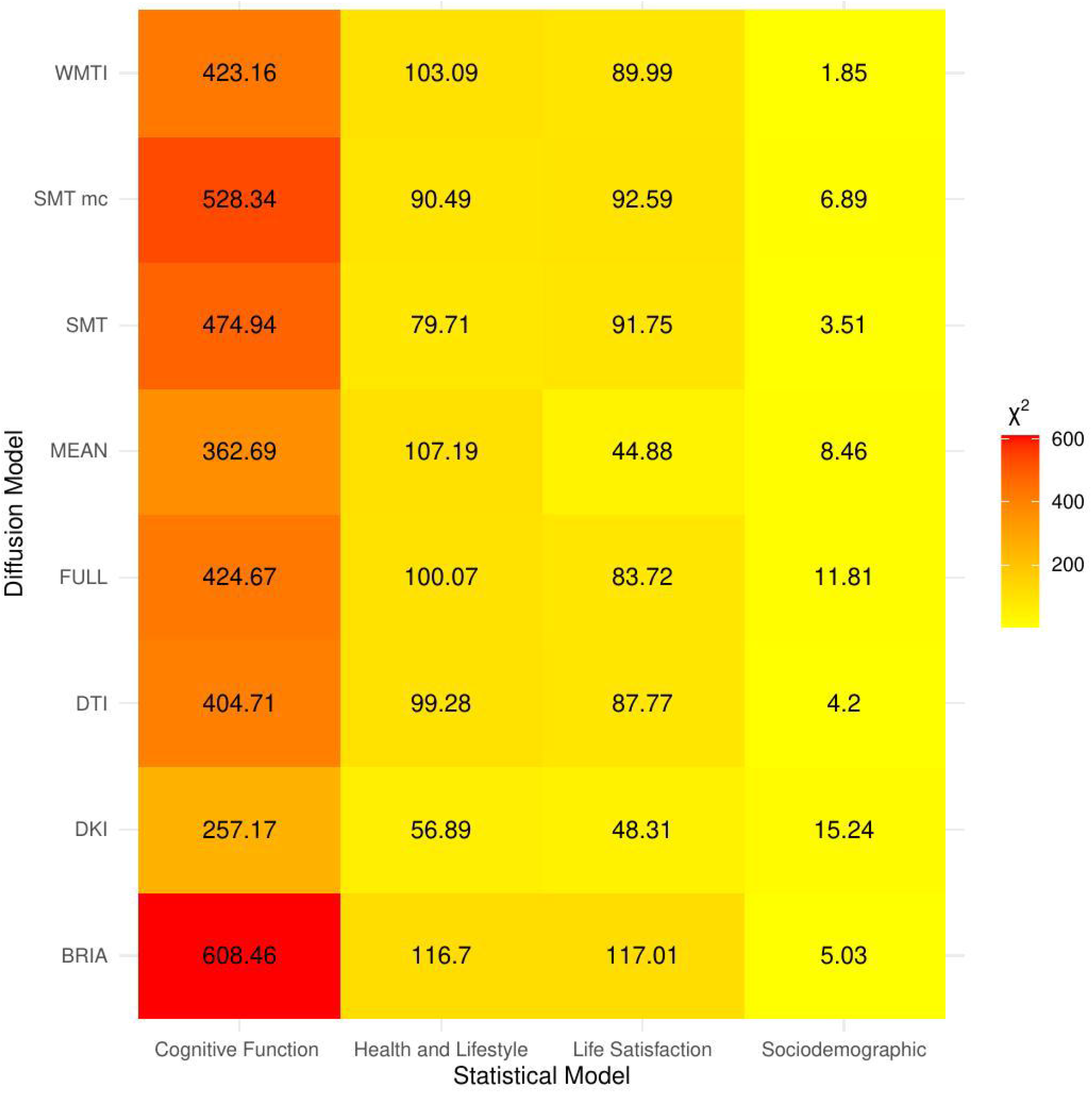
Overview of Comparison of Bio-Psycho-Social Statistical Models with Baseline Models The table presents χ^2^ values for each of the bio-psycho-social statistical models for each diffusion approach tested against the baseline model. Note that only values of χ^2^ > 11 were significant (*p* < .05).

**Figure 3.**
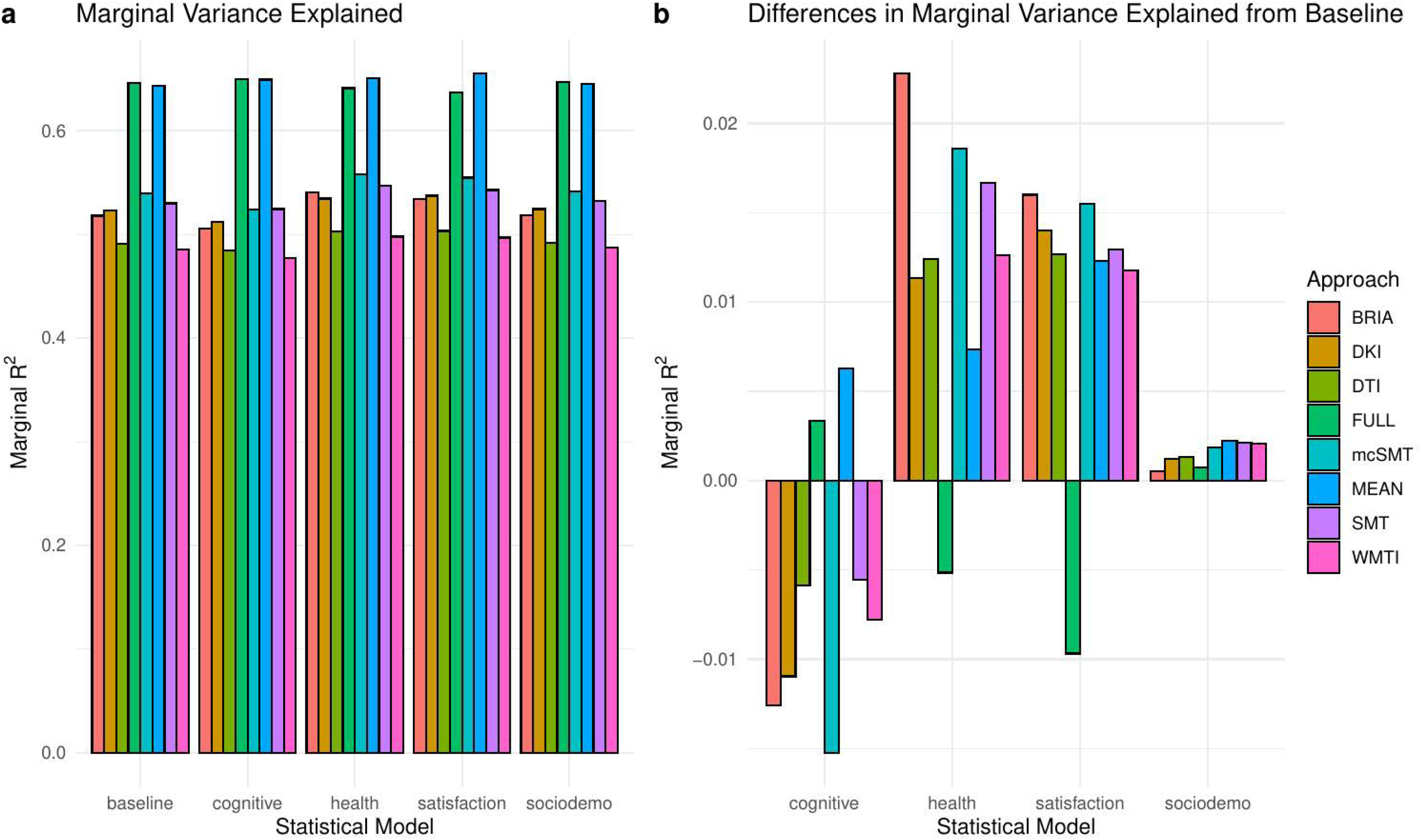
Marginal R^2^ Values for Statistical Models across Diffusion Approaches

**Figure 3a** shows the total marginal variance in brain age explained from the different models from (1) baseline, (2) social factors, (3) cognitive test scores, (4) life satisfaction, and (5) health and lifestyle factors (with weight on cardiometabolic factors). **Figure 3b** shows the marginal variance of brain age explained by other models above that of the baseline model.

Across statistical and diffusion models, age was used as a control variable to correct for the mere reflection of age by brain age producing stable associations across models (1-5) for multimodal brain age (**Figure 4-7**). However, except for the life-satisfaction model, in contrast to the full multimodal model, the other diffusion approaches’ brain ages were negatively associated with age, giving another indication of overall poor model fit. Even more so, the effect of sex was dependent on the model, producing mixed effects with large uncertainty surrounding *β-*values, also in the age-by-sex interactions’ associations with brain age. Overall, bio-psycho-social factors were consistently associated with brain ages from different diffusion approaches, with the exception for sex (**Figure 4-7**).

**Figure 4.**
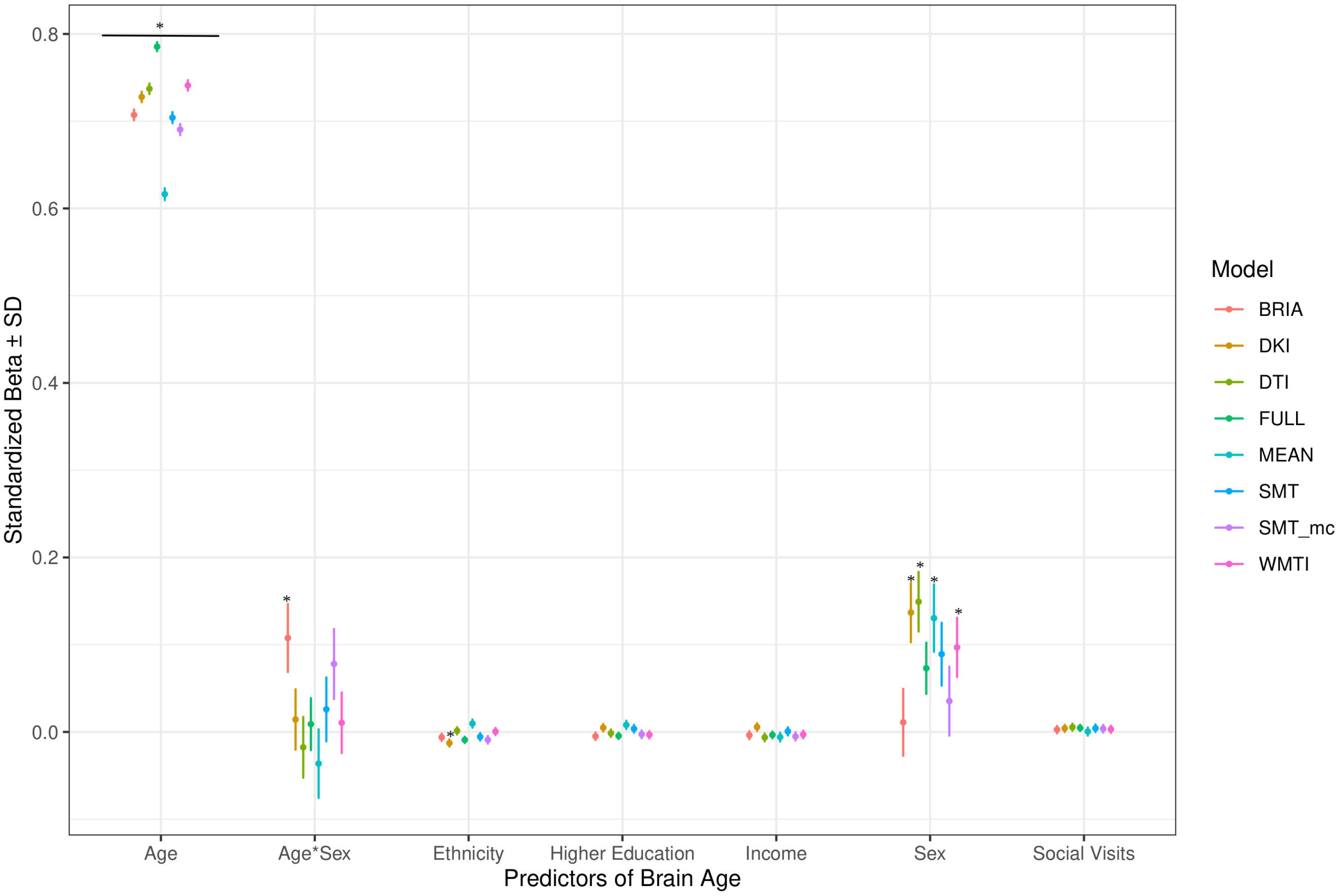
Sociodemographic Model Predictors’ Standardized Beta-Values with Standard Deviation indicates Bonferroni-corrected *p* < .05

#### 3.1.1 Sociodemographic factors’ associations with brain age

In the model including sociodemographic factors explaining brain age (see **Figure 4** for the predictors), results were mixed for the significant predictors. Sex was a significant predictor for mean DKI, DTI, and WMTI (*ps* < .05), the age-by-sex interaction only for BRIA (*p* = .045), and ethnicity only for DKI (*p* = .012; **Figure 4**). Overall, only 95% confidence intervals of *β-*values for age and ethnicity were not consistently overlapping, indicating differential effects of these variables on brain age based on the underlying data. All other 95% confidence intervals surrounding coefficients’ *β-*values were overlapping across diffusion approaches, with expected strong age contributions predicting brain age.

#### 3.1.2 Health and lifestyle factors’ associations with brain age

Similarly, in the model including health and lifestyle factors explaining brain age (see **Figure 5** for the predictors), significant health factors leading to higher brain age were WHR (*ps* < .001), pulse pressure (*ps* < .001), and hypertension (*ps* < .001). Evidence across diffusion approaches was mixed for the other predictors with smoking predicting brain age derived from BRIA, DTI, mean scores, SNT, and WMTI (*ps* < .05), diabetes diagnosis for all models except DKI and DTI(*ps* < .05), the diagnosis of at least one vascular disease for BRIA, mean scores, and mcSMT (*ps* < .02), and average daily cups of coffee for brain age estimates except the one based on BRIA (*ps* < .01), and the age-by-sex interaction for BRIA and mcSMT (*ps* < .04).

**Figure 5.**
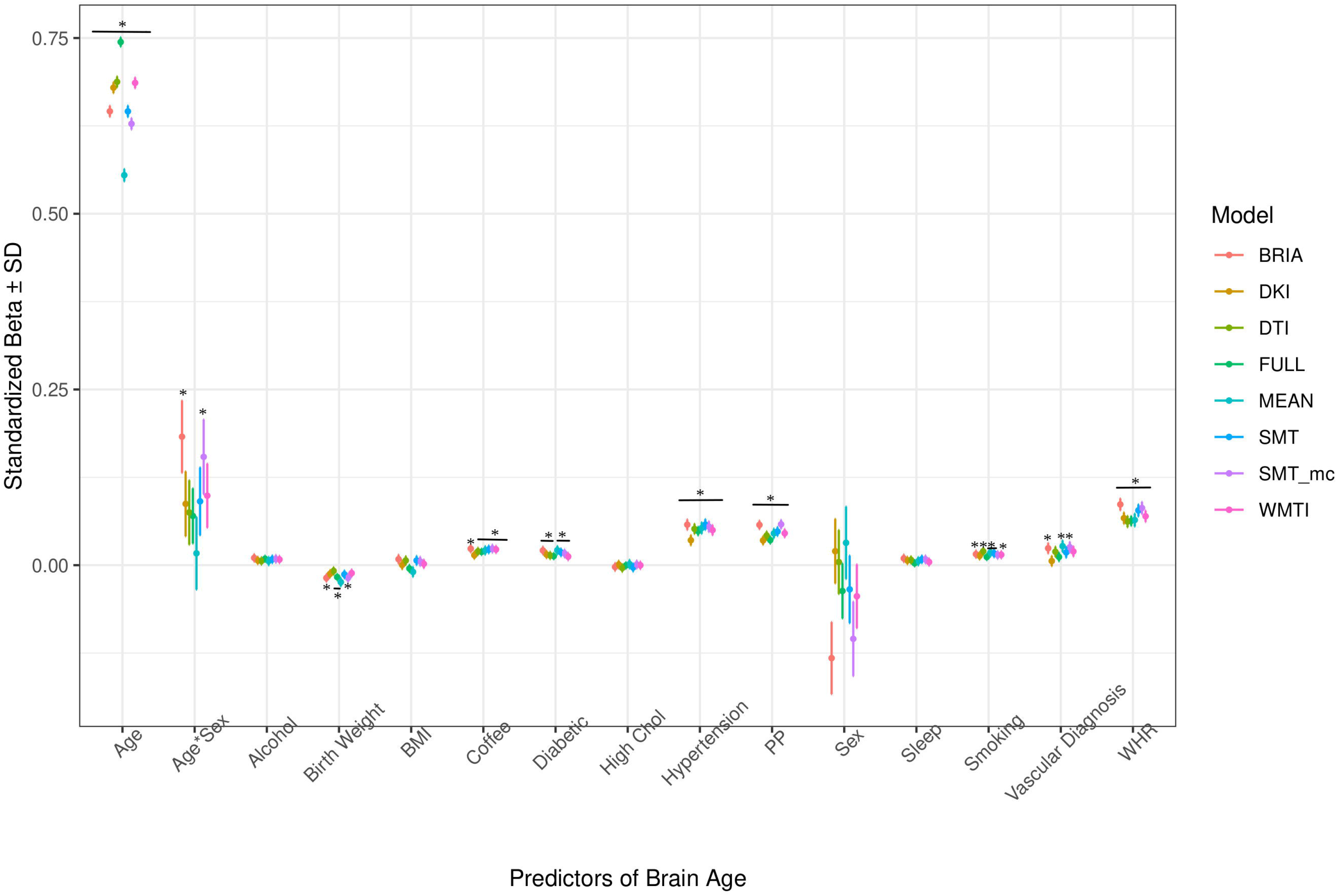
Health Model Predictors’ Standardized Beta-Values with Standard Deviation indicates Bonferroni-corrected *p* < .05

Interestingly, WHR was a stronger predictor of brain age in males than in females (**Supplementary Figure 5**). Practically, a WHR *β*_*unstd*_-value of, for example *β* = 4 would mean that for every 0.1 step change in WHR, the brain age can be expected to increase by 0.4 years (see Supplementary Figure 6-9 for *β*_*unstd*_). Importantly, this association was controlled for age, as age is correlated with WHR at *r* = 0.14 and brain age at *r* = 0.80. Mean population values for WHR were found to be WHR < 1 (Molarius et al., 1999), with our sample corresponding with these estimates (M_WHR_ = 0.871±0.088, min = 0.534, max = 1.472) with males having a higher WHR (M_WHR_ = 0.923±0.064) than females (M_WHR_ = 0.817±0.069).

BMI was potentially non-significant due to the model construction as the highly correlated WHR (**Supplementary Figure 4**) was a significant predictor of brain age, and BMI alone being a significant predictor of brain age (**Table 2**). Finally, higher birth weight was associated with lower brain age estimated from full and mean models, as well as BRIA and WMTI (*ps* < .02).

**Table 2:**
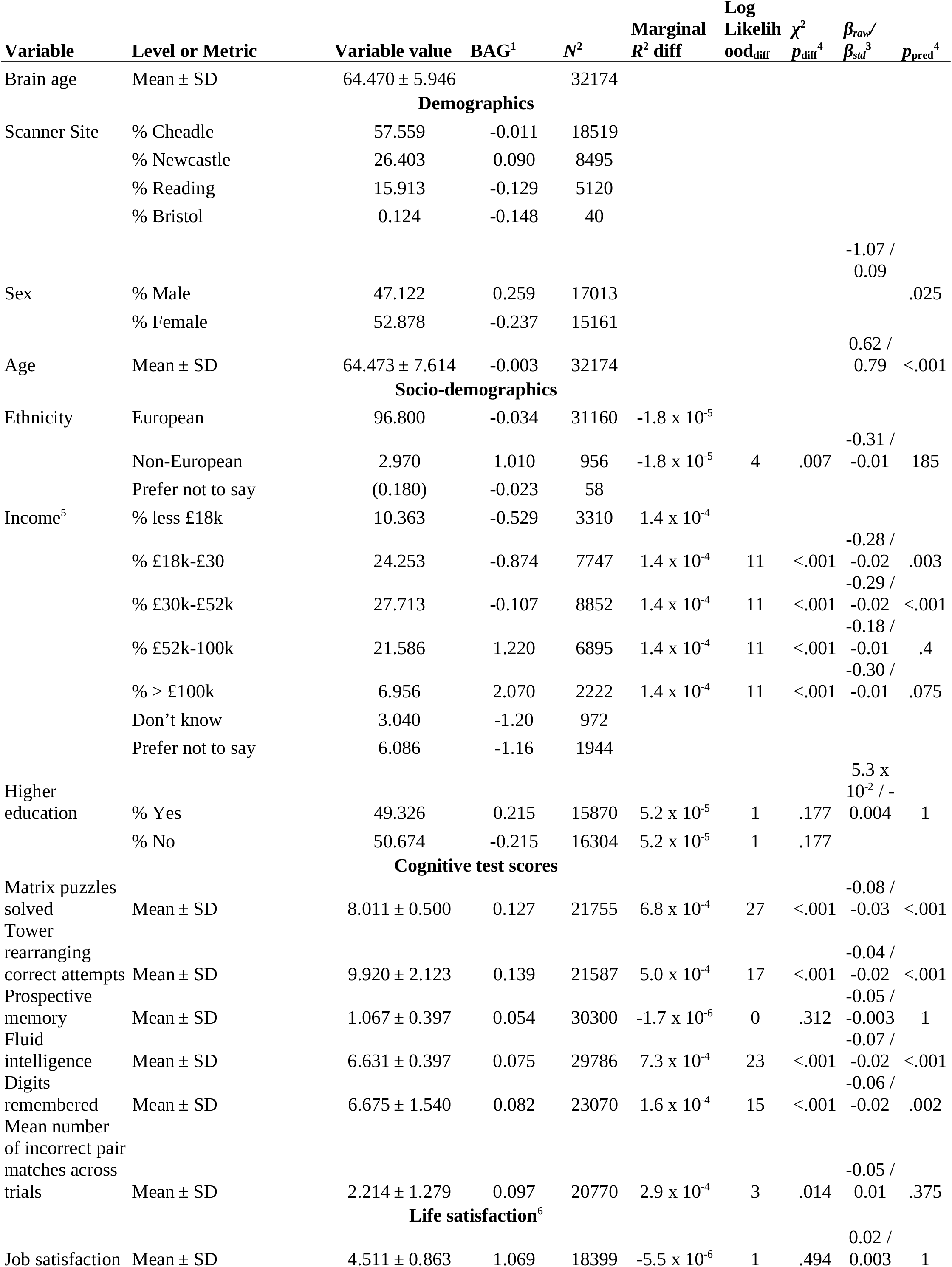
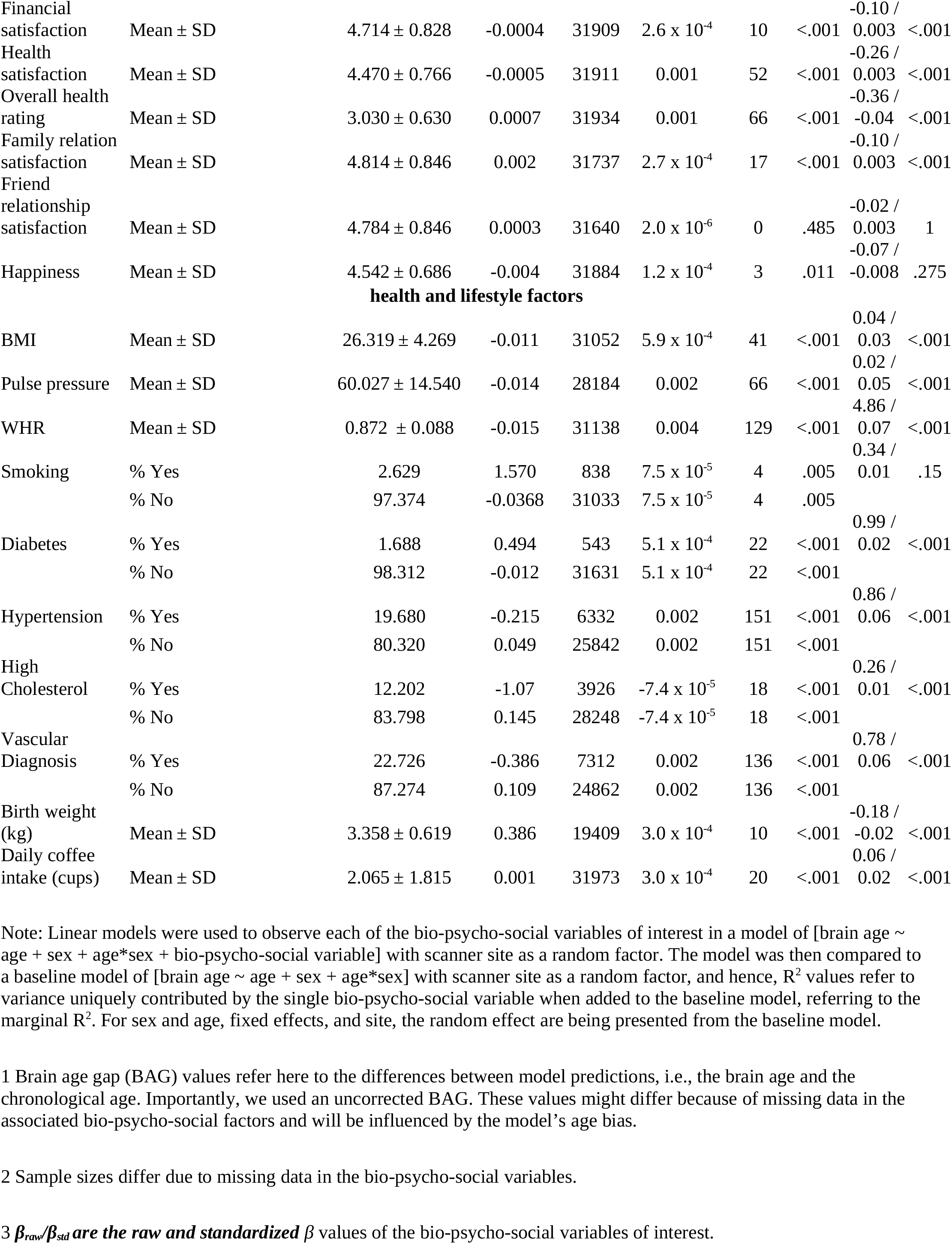
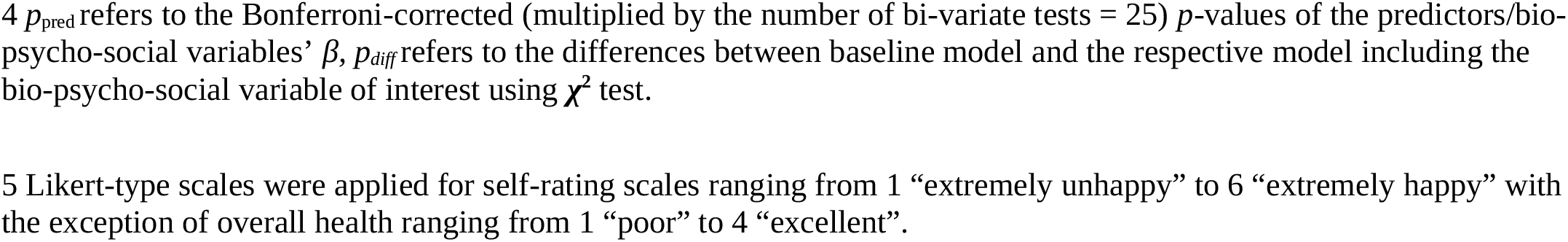
Linear models relating multimodal brain age to bio-psycho-social factors

Generally, 95% confidence intervals around coefficients’ β-values were overlapping across models indicating no significant differences in β-values across diffusion approaches. As a control, we ran the same model without WHR as predictor, due to its high correlation with BMI, rendering BMI as significant predictor across diffusion approaches’ brain ages except the mean model (βs > 0.01 ps < .004), also showing now clearer evidence for higher brain age when smoking (ps < .05), with other predictors unchanged (**Supplementary Figure 11**). Furthermore, leaving out hypertension, being a substrate of blood pressure, did not lead to changes in the model (**Supplementary Figure 12**). For both models, variance explained is slightly reduced compared to the models including the respective variables, making the reduced models significantly different (ps < .001) from the full health models (**Supplementary Table 8-9)**.

#### 3.1.3 Life satisfaction factors’ associations with brain age

When modeling brain age from life satisfaction (see **Figure 6** for the predictors), self-rated health was a significant predictor of all brain age estimates except for DKI brain age (*ps* < .05) and health satisfaction for all brain age estimates except the mean model’s brain age (*ps* < .02). Only the 95% confidence intervals of *β-*values for age do not overlap across models (with the mean model having the largest *β* and full model the smallest *β-*value for age). All other 95% confidence intervals around coefficients’ *β-*values overlap across models indicating no significant differences in *β-*values across diffusion approaches.

**Figure 6.**
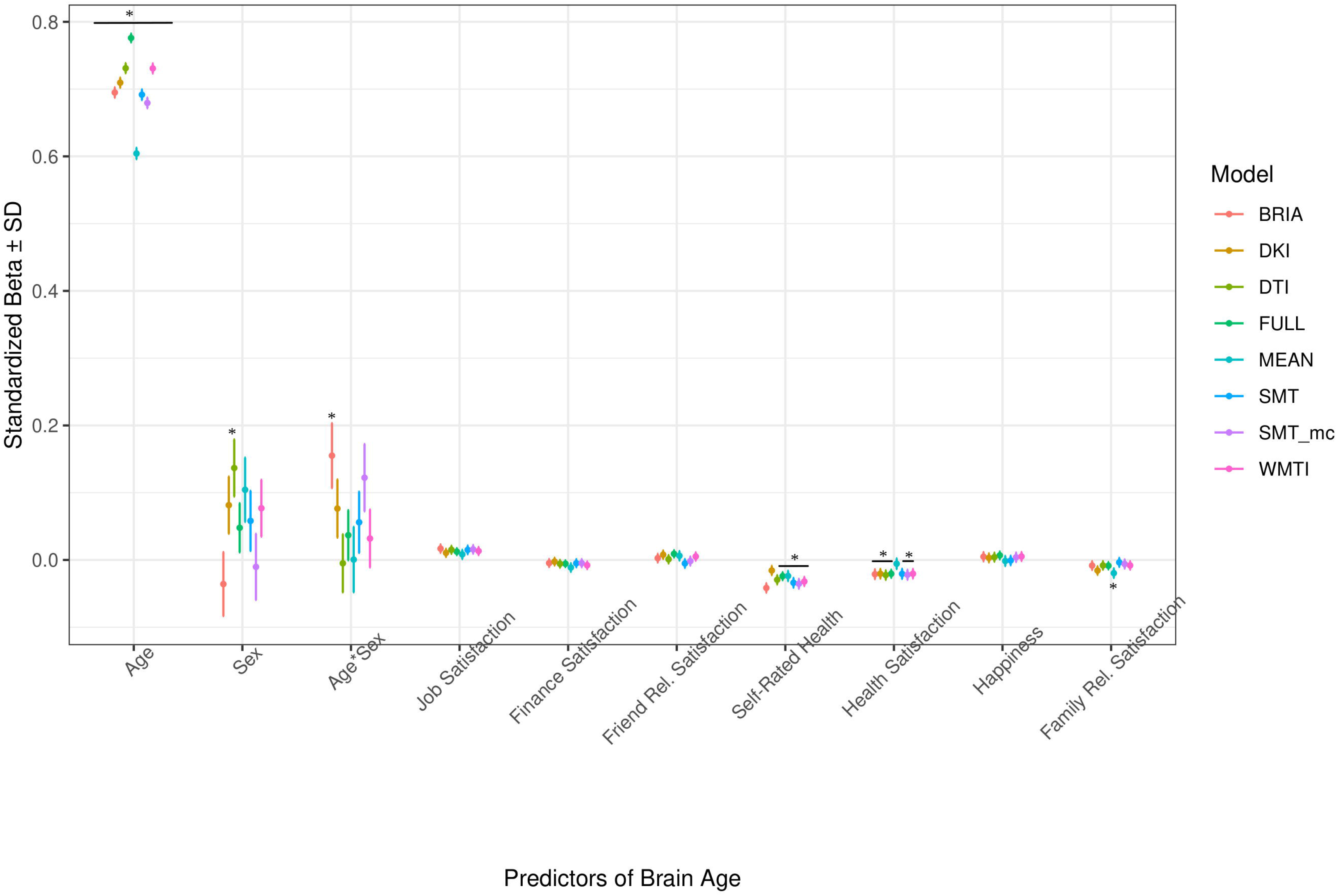
Well-being Model Predictors’ Standardized Beta-Values with Standard Deviation indicates Bonferroni-corrected *p* < .05

Perceived health is moderately correlated with health satisfaction and was left out in a control model resulting in a slightly stronger effect of health satisfaction and significantly worse performing model (*ps* < .001; **Supplementary Figure 12, Supplementary Table 8-9**). Differently, when leaving out happiness as being correlated with several variables the model remains unaffected (*ps* > .23; **Supplementary Figure 13, Supplementary Table 8-9**).

#### 3.1.4 Cognitive factors’ associations with brain age

The only cognitive factor explaining brain age across all models was symbol digit substitution (*ps* < .001; **Figure 7**). Matrix puzzles solved was only a significant predictor for the full multimodal brain age (*p* = .014), and sex only for DTI and WMTI (*ps* < .02). Confidence intervals around coefficients’ *β-*values are overlapping across models indicating no significant differences in *β-*values across diffusion approaches. Fluid intelligence and matrix puzzles are highly correlated and hence, matrix puzzles were left out in a quality control model, not significantly affecting the structure of most models (**Supplementary Figure 14, Supplementary Table 8-9**).

**Figure 7.**
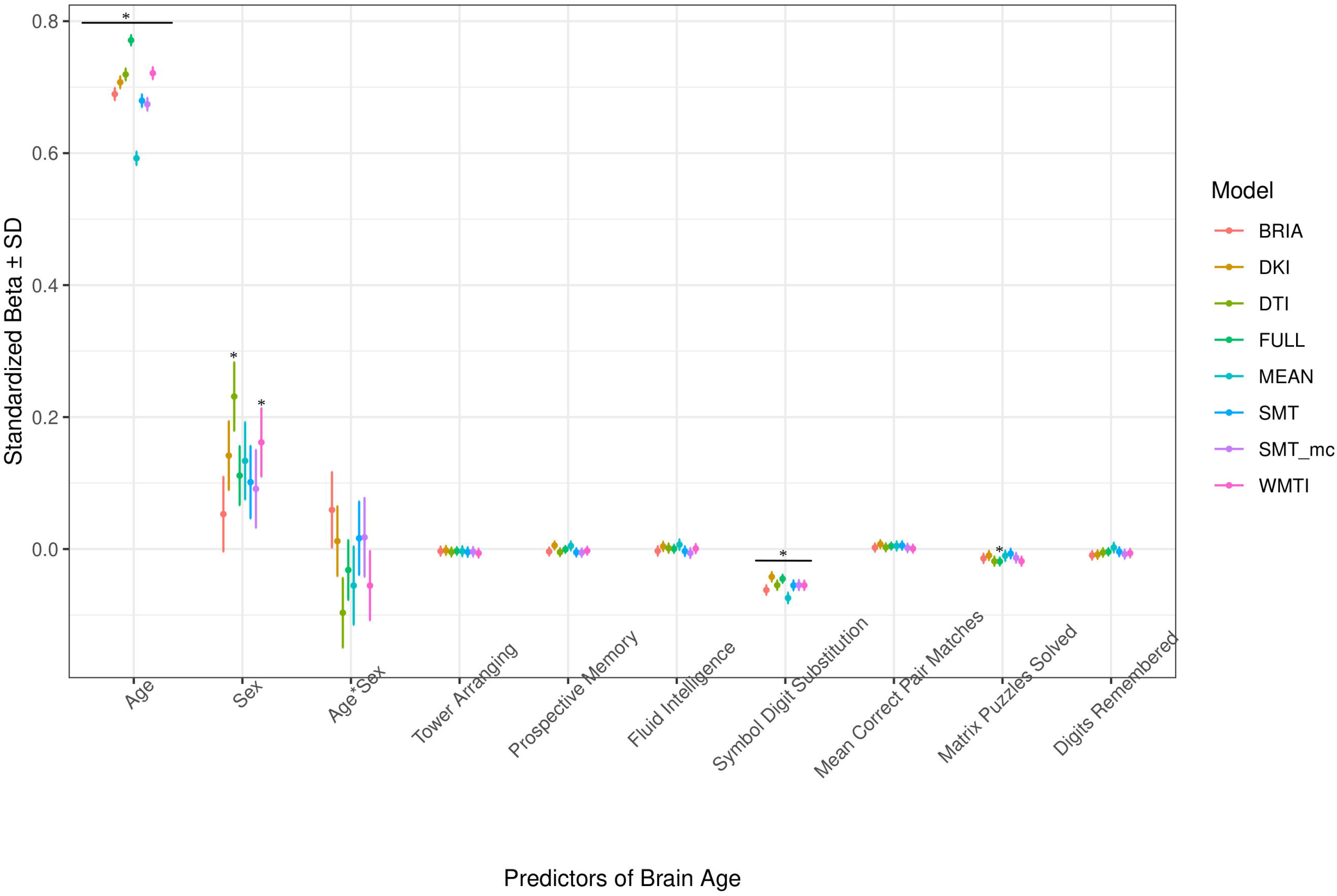
Cognition Model Predictors’ Standardized Beta-Values with Standard Deviation indicates Bonferroni-corrected *p* < .05

#### 3.1.5 Follow-up: quality control and bivariate relationships of multimodal brain age and bio-psycho-social factors

Due to the strong variability in sex *β-*values across models (**Figures 3-6**), we also ran the described analyses separately for males and females showing some differences in model performance. For example, bio-psycho-social models explained a differential of between 1%-4% of conditional variance for males (**Supplementary Table 10**) and differences in contributions of the different models’ predictors, predictors’ *β-*values being generally higher for males (**Supplementary Figure 2**). Overall, quality checks show small levels of multicollinearity, and that each predictor contributes individual to the models (**Supplementary Figures 2-10, Supplementary Tables 8-9**), supporting assumptions about the robustness of the utilized models, as well as that simply adding all variables together saturates the model leading to lower model performance than at baseline across brain ages based on different diffusion approaches with a differential in marginal R^2^ = 3.38%.

Finally, for a better understanding of bivariate relationships, **Table 2** gives an overview of brain age calculated from combined single and multi-shell diffusion data in relation to the observed bio-psycho-social factors. Strongest standardized associations when adding single factors to a model explaining brain age from age were found for WHR (*β*_*std*_ = 0.07, *p* < .001), PP (*β*_*std*_ =05, *p* < .001), and overall health rating (*β*_*std*_ = -0.04, *p* < .001), and health satisfaction (*β*_*std*_ = -0.03, *p* < .001). Strongest brain age group differences were found for sex (*β*_*std*_ = -0.09, *p* = .001), diabetes (*β*_*std*_ = 0.02, *p* < .001), and hypertension (*β*_*std*_ = 0.06, *p* < .001).

## 4 Discussion

We assessed the influence of various bio-psycho-social variables on brain age estimated from different diffusion approaches (and their combinations). As predicted, linear mixed effects models showed that bio-psycho-social variables uniquely explain a small proportion of brain age variability consistently across models and predictors’ *β-*values’ confidence intervals overlap for most predictors. Health and lifestyle factors were most indicative of brain age. However, differences in brain age variance explained between bio-psycho-social models and diffusion approaches were small. Significant predictors of brain age were job satisfaction, health satisfaction, WHR (and to a lesser extent BMI when excluding WHR as a predictor), diabetes, hypertension, any vascular diagnosis, daily coffee consumption, smoking, birth weight, matrix puzzles, and symbol digit substitution performance. Our findings indicate that brain age estimates derived from different diffusion approaches relate similarly to the examined bio-psycho-social factors. This is an important finding as it reveals that different WM characteristics share common aging associations, which are detailed by bio-psycho-social factor associations. The presented diffusion approaches are based on different theoretical assumptions for deriving a set of WM features. For example, DTI and DKI metrics are usually quite sensitive to a broad range of WM changes due to their integrative nature of the scalar metrics (Basser et al., 1994; Jensen et al., 2005), i.e. DTI’s FA or DKI’s MK allow one to detect and localize the WM changes but not to explain their origins. In turn, dMRI approaches such as SMTmc or BRIA offer several metrics potentially allowing us to bind WM architecture with their predictive power (Kaden et al., 2016a, 2016b; Reisert et al., 2017). For example, the intra-axonal water fraction appearing in both models might correlate with axonal density and axon diameter (Jelescu et al., 2020). Consequently, the metric provides information about WM maturation associated with aging leading to similar associations with age, aging and aging-related variables as DTI/DKI models. This encourages the usage of both conventional and advanced diffusion approaches when examining the relationship of bio-psycho-social factors and WM. Particularly, the application of dMRI approaches with more accurate assumptions around biophysical processes such as a ratio between intra- and extra-axonal diffusivities, permeability and other features offers various opportunities to investigate aging and associated diseases.

### 4.1 Explaining brain age from bio-psycho-social factors

Recent research has made a strong case for the conjunction effects of various bio-psycho-social factors in explaining general health (Lehman et al., 2017). Applied to brain age, for example, cardiometabolic effects have been shown to influence brain age (Beck et al., 2022a, 2022b). However, assessments of how much of the variance explained in brain age above and beyond age, sex, age-by-sex interaction, and scanner site have not been described in the literature. We find close-to-zero added brain age variance explained by models including single bio-psycho-social variables (**Figure 2**). Comparably more, yet still a small proportion of brain age variance (R^2^ < 4%) is uniquely explained by bio-psycho-social variable groups of socio-demographics, health and lifestyle, life satisfaction, and cognitive ability (**Figure 3, Supplementary Table 7**). A potential reason for small R^2^ values might lay in multiple confounder effects and heterogeneity in effects across covariate levels (table 2 fallacy, Westreich & Greenland, 2013). Importantly, the added brain age variance explained is not just an effect of adding predictors randomly to the model, which rather decreases the variance explained, as shown when adding all bio-psycho-social variables to the model. Hence, it seems more sensible to employ models incorporating several compared to single domain-specific variables to explain brain age. However, our results also indicate that a large part of the variance in brain age cannot be explained by our proposed bio-psycho-social models. Whether this unexplained variance is due to actual biologically founded individual differences, or the characteristics of brain age, for example, how the metric is being estimated (de Lange et al., 2022), remains unclear. BAG might also be rather static and indicated by constants such as genetic architecture and birth weight (Vidal-Pineiro et al., 2021). This would explain the smaller influence of more variable bio-psycho-social variables. Strong deviations from the norm, for example, due to atrophy will also have a strong influence on brain age (Kaufmann et al., 2019). Hence, for diseases impacting brain structure, brain age can be a useful indicator of health status (Kaufmann et al., 2019). Potentially, the health and lifestyle factors which are most likely to impact brain structure are therefore also more predictive of brain age than other bio-psycho-social variables (**Figure 4-7**). While our models failed to explain larger proportions of the variance of brain age, there are various interesting phenotype associations within these models which will be discussed in the following.

#### 4.1.1 The importance of age, sex, and ethnicity

Usually, age, sex, and at times, scanner site, are used as covariates for brain age-phenotype associations as they are expected to influence various phenotypes (Jirsaraie et al., 2022). As brain age reflects chronological age, age also explains most of the brain age variance (**Figure 4-7**). We also find that the effects of sex and the sex-age interaction were highly variable across diffusion models predicting brain age with sex and the sex-age interaction being mostly non-significant predictors across diffusion models (**Figure 4-7**). Nevertheless, brain age does significantly differ between sexes (Sanford et al., 2022; Subramaniapillai et al., 2022), and we cannot exclude sex difference in WM microstructure. These relationships might also lead to differences in WM brain ages between sexes. Furthermore, models were more predictive of bio-psycho-social factors in males than females (**Supplementary Table 2, Supplementary Figure 2**). Where the influence of sex changes based on the model construction, while potentially also influencing the model (**Figure 4-7**; **Supplementary Figure 5-8**). Some of the observed sex differences might be based on anatomical features, such as higher intracranial volume in males and different sex-specific aging (Eikenes et al., 2022). Brain age was differentially sensitive to ethnicity dependent on the approach it was calculated on (**Figure 4-7**), with these differences being influenced by sex (**Supplementary Figure 2**). A previous study showed that being a UK immigrant might influence brain age estimates (Leonardsen et al., 2022). Potentially, genetic contributions to brain age both estimated from T1-weighted (Ning et al., 2020; Vidal-Pineiro et al., 2021) and dMRI data (Salih et al., 2021) also have a connection with the mentioned brain age differences by sex and ethnicity. However, the causal structure of sex and ethnicity differences in brain age estimates requires further investigation.

Previous research has shown the effects of sex on metrics derived from conventional and advanced diffusion approaches, such as BRIA, DKI, DTI, NODDI, RSI, SMT, SMT mc, and WMTI (Beck et al., 2021; Eikenes et al., 2022). While a systematic assessment of sex-related effects on diffusion metrics from both conventional and advanced dMRI approaches from voxel-to-whole-brain averages over the lifespan is yet to be established, different studies presented sex-related developmental trajectories in the structural connectome in children (Ingalhalikar et al., 2013), and sex related WM changes during aging (Hsu et al., 2008). Furthermore, sex differences in aging reflected in WM microstructure can be expected due to menopause and cascading biological processes, affecting both brain and body systems in various ways (Barth & de Lange, 2020; Lohner et al., 2022; Mosconi et al., 2021). Hence, developmental trajectories differing between males and females can be expected which makes sex-separated analyses useful to providing important additional information (e.g., as in Subramaniapillai et al., 2022). To which extend this applies to ethnicity requires further research. Hence, further research is required to delineate the underlying causal structure of sex and ethnicity to explain their highly variable associations with brain age.

#### 4.1.2 Health and lifestyle factors

Interestingly, while the health and lifestyle factors models explained only a small proportion of the brain age variance, most of its predictors were significant. Furthermore, these predictors are generally only weakly correlated (**Supplementary Figure 5**), but when added in conjunction explaining more variability in brain age than on their own (compare **Table 2** and **Figure 5**). To a certain degree, this is not surprising, due to dependencies between these predictors. For example, WHR, being the strongest predictor of brain age (see **Figure 5**), shows a clear relationship with pulse pressure (**Supplementary Figure 5**). For the extreme cases, this is expressed in a well-established relationship between obesity and hypertension (Kotsis et al., 2010) or any vascular diagnosis (Mathew et al., 2008). This is reflected in brain age, where minimum and maximum values show that there is an expected difference of up to 4 years in brain age between those with lowest compared to highest WHR, or a 2.4-years brain age difference between mean and maximum WHR. Interestingly, blood pressure is expected to increase with age, and higher blood pressure is positively associated with BAG (Cherbuin et al., 2021). However, these effects were not exclusively driven by hypertension but across the spectrum of measured blood pressure values (Cherbuin et al., 2021). This was supported by our findings showing both an effect of pulse pressure and hypertension on brain age. These effects are not surprising, as hypertension has been suggested as one of the most important risk factors for various cerebrovascular complications such as cerebral small vessel disease and resulting cognitive impairments (Forte et al., 2020; Meissner, 2016).

Another aspect of high WHR and BMI is obesity increasing diabetes risk (Kahn et al., 2006). While the evidence for the direction of the effect of diabetes is mixed (Cole et al., 2018; Franke et al., 2013; Sone et al., 2022), we find participants with diabetes to show higher brain age than those without diabetes (**Table 1, Figure 5**). Several complications within the central nervous system have been associated with diabetes, including morphological, electrophysiological, and cognitive changes, often in the hippocampus (Wrighten et al., 2009), just as WM lesions and altered metabolite ratios (Biessels & Reijmer, 2014, van der Harten et al., 2006), supporting the idea of higher brain age among those with diabetes. But also generally, the increase in risk of cardiovascular disease by WHR is mediated by BMI, systolic blood pressure, diabetes, lipids, and smoking (Gill et al., 2021). In relation to the brain, higher WHR has been generally associated with lower gray matter volume (Gurholt et al., 2021; Hamer & Batty, 2019), and higher WM brain age (Beck et al., 2022a, 2022b; Subramaniapillai et al., 2022). Hence, to which extent high WHR accelerates brain aging requires further investigation, which might be particularly informative when observed in combination with other health and lifestyle variables (Hamer & Batty, 2019) and sex (Subramaniapillai et al., 2022).

Negative health consequences of smoking (Erhardt, 2009) are reflected in smoker’s cortex being thinner (Gurholt et al., 2021), and smokers’ brains being 1.5 years older on average than non-smokers’ brains (**Table 2**). Smoking is a known risk factor for cardiovascular health significantly increasing its mortality and inducing various negative downstream effects on health (Erhardt, 2009), with negative impacts on the reward system (Le Foll et al., 2022), repeatedly shown in rats (e.g., Cao et al., 2013; Gozzi et al., 2006; Kenny & Markou, 2006). It can hence be expected that both general and brain health are influenced by smoking, making it an important control variable in assessing brain age.

The findings for coffee on the other hand are mixed, suggesting coffee consumption to be generally positive for cardiovascular health and decreasing the risk of Parkinson’s disease, stroke, and Alzheimer’s (Nehling, 2016). The consumption of higher doses of caffeine is, however, associated with smaller brain volume and an increased risk of dementia (Pham et al., 2021). Practically, the direct effect of the number of daily cups of coffee consumed is small in our study. It would require on average 10 cups of coffee daily for an increase of 0.6 years of brain age, fitting the observations made by Pham et al. (2021). It also remains unclear whether the effect of coffee consumption on brain age is rather mediated by third variables such as poor sleep and mental health downstream effects which show direct negative effects on health (Distelberg et al., 2017). Additionally, there are vulnerable groups in which caffeine can cause adverse effects such as people with hypertension (Higdon & Frei, 2007). We conclude that health and lifestyle factors function in synergy in influencing brain age.

#### 4.1.3 Health perception and satisfaction, and job satisfaction

We find significant assignations of self-rated health, friendship/relationship satisfaction, and job satisfaction with brain age. Self-assessments and self-rated scores are some of the fastest and easiest assessments. Yet, their reliability is under constant scrutiny, particularly when assessing health outcomes (e.g., Crossley & Kennedy, 2002; Reychav et al., 2019). In our study, self-rated overall health was a significant predictor of brain age, suggesting that asking participants about their health can be a useful preliminary assessment of different aspects of health. Self-rated health was additionally moderately correlated with health perception (**Supplementary Figure 4**), indicating both variables measure, to a certain degree, the same underlying phenomenon. However, self-rated health brain age associations were stronger and more variable across diffusion approaches’ brain ages (Figure 6). These associations support the idea of brain age is not only indicative of brain health, but also overall health (Kaufmann et al., 2019).

Lastly, there was a trend of individuals’ job satisfaction being associated with brain age (**Figure 6**). Conceptually, this would not be surprising as associations between wealth and health (e.g., Adler & Ostrove, 1999) as well as job (e.g., Faragher et al., 2013) and financial satisfaction and health (e.g., Hsieh, 2001) have already been investigated. However, in the case of our study, higher job satisfaction was also indicative of higher brain age. Potential reasons are speculative but might reflect the tendency of people engaged in their jobs to work long hours which has previously been related with various negative mental and physical health outcomes (Bannai & Akiko, 2014; Lim et al., 2011). Nevertheless, the underlying mechanisms of the associations between these single items in their relationship with brain age require further investigation.

#### 4.1.4 Cognitive scores

Cognitive scores’ impact on brain age might be small in the current study, yet still important in general (**Table 2**). This might be due to the selection of the observed cognitive test scores, with many more possible tests to be included which are potentially more indicative of brain age, such as IQ (Elliott et al., 2021). Another opportunity lies in assessing associations of cognitive performance and brain age in clinical groups. For example, brain age has been found to be explanatory of symbol digit modality test scores in multiple sclerosis suggesting brain age as a biomarker for cognitive dysfunction (Denissen et al., 2022). Similar to such findings, we find a similarly sized effect of symbol digit substitution test scores in our healthy aging data (**Figure 7**). Associations of cognitive performance and brain age are also sensitive to sex. For example, the number of solved matrix puzzles showing an effect when analyzing males and females data together seemed to be a predictor of brain age only in females when analyzing females from males data separately (**Supplementary Figure 2**). The quality of these differences requires further investigation.

### 4.2 Variability in brain age-phenotype relationships

Imaging phenotypes derived from diffusion UKB data contribute to a small additional proportion of the variability in the obtained results. However, the presented comparison of R^2^ differences (**Figure 2**) underestimates the effects of single bio-psycho-social factors, and has to be interpreted with care, with cognitive function, life satisfaction, and health and lifestyle factors significantly adding to the baseline model (**Figure 2**). Yet, the used brain age estimation model might also introduce variability in brain age phenotype associations. Problematically, model evaluation metrics such as R^2^, MAE, or RMSE depend additionally on cohort- and study-specific data characteristics making brain age model comparison across the literature not straightforward (de Lange et al., 2022). Additionally, there are differences between models trained on voxel-level compared to region-averaged data. Deep learning models using voxel-level data reach age predictions errors as low as MAE = 2.14 years in midlife to late adulthood (Peng et al., 2021) or MAE = 3.90 years across the lifespan (Leonardsen et al., 2022) while explaining large proportions of variance in age (R^2^ > 0.90), whereas models trained on regional and global average measures predict age usually with larger error, MAE > 3.6 years, and/or lower variances explained R^2^ < 0.75 (Beck et al., 2021, 2022; de Lange et al., 2020a, 2020b; Korbmacher et al., 2022; Rokicki et al., 2021). However, Niu et al. (2019) showed that with different shallow and deep machine learning algorithms (ridge regression, support vector regressor, Gaussian process regressor, deep neural networks) high prediction accuracies (R^2^ > 0.75, MAE < 1.43) could be reached when using multimodal regional average data using a young sample with narrow age range. Nonetheless, when comparing brain age retrieved from the same database (UKB), but using different samples, modalities, and methods to calculate brain age, similar patterns of associations between brain age and phenotypes can be observed in this study. For example, diabetes diagnosis, diagnosed vascular problems or place of birth (see Figure 4 in Leonardsen et al., 2022), hip circumference, diabetes, trail-making tasks (here symbol digit substitution), and matrix pattern completion were significantly associated with brain age (see Table 5 in Cole, 2020). Nevertheless, it remains unclear whether the differences in the findings are due to analyst degrees of freedom, sample characteristics, or actual bio-physical manifestations. For example, the underlying data used to estimate brain age, being dMRI in this study were T1-weighted images in Cole (2020) and Leonardsen et al. (2022). From our findings, we can at least assume that phenotype-WM brain age associations are relatively stable across diffusion approaches. Furthermore, phenotype data processing, such as dummy coding, differed, as well as models to predict brain age. Here we used four mixed models grouping a) demographics, b) cognitive, c) life satisfaction, and d) health and lifestyle variables to predict brain age. In contrast, Cole (2020) predicted bias-adjusted brain age from simple linear models with sex, age, and age^2^ as covariates, and Leonardsen et al. (2022) observed similar associations for uncorrected brain age predicted from the respective phenotype and age and sex as covariates. However, bio-psycho-social variables are likely to interact forming a complex pattern when explaining variables such as brain age. When adding only single bio-psycho-social variables to a baseline model and then comparing the two models, the differences in variance explained are smaller than when adding blocks of meaningfully related variables (compare **Table 2** and **Supplementary Table 7**). The composition of variables forming such blocks/models requires further investigation. In sum, there are various sources introducing variability in brain age phenotype associations encompassing not only the underlying data but also researchers’ degrees of freedom such as data selection, processing, and analysis.

### 4.3 Conclusion and future directions

Bio-psycho-social factors contribute similarly to explaining WM brain age across conventional and advanced diffusion MRI approaches when arranged as cognitve scores, life satisfaction, health and lifestyle factors, but not socio-demographics. Focussing on single predictors, health and lifestyle factors, WHR, birth weight, diabetes, hypertension, and related diagnoses, as well as smoking status and coffee consumption, were more predictive of brain age than cognitive and life satisfaction measures. Apart from health satisfaction and self-ratings, we found relationships of life satisfaction variables with brain age to be non-significant. Of the cognitive scores, only the digit substitution task performance was a significant predictor, which might be relevant in samples from midlife to old age. Furthermore, the influence of sex and ethnicity is largely variable suggesting the usage of sensible control mechanisms, such as separate analyses or exclusions in case of strongly imbalanced samples. We recommend future study designs taking observable interactions between the different bio-psycho-social effects into account. A potentially helpful guiding principle in the search for bio-psycho-social variables affecting brain age could be to focus on measures which are directly or indirectly related to or reflect pathology.

## Data Availability

All data produced are available online after obtaining permission from the UK Biobank: https://www.ukbiobank.ac.uk/

## 8 Author Contributions

M.K.: Study design, Software, Formal analysis, Visualizations, Project administration, Writing – original draft, Writing – review & editing

T.P.G. Writing – review & editing

A.M.d.L.: Software, Writing – review & editing

D.v.d.M.: Software, Writing – review & editing

A.L.: Writing – review & editing, Funding acquisition

E.E.: Writing – review & editing, Funding acquisition

D.B.: Writing – review & editing

O.A.A.: Writing – review & editing, Funding acquisition

L.T.W.: Writing – review & editing, Funding acquisition

I.I.M. supervision, Study design, Data pre-processing and quality control, Writing – review & editing, Funding acquisition

## 10 Acknowledgments

This study has been conducted using UKB data under Application 27412. UKB has received ethics approval from the National Health Service National Research Ethics Service (ref 11/NW/0382). The work was performed on the Service for Sensitive Data (TSD) platform, owned by the University of Oslo, operated and developed by the TSD service group at the University of Oslo IT-Department (USIT). Computations were performed using resources provided by UNINETT Sigma2 – the National Infrastructure for High Performance Computing and Data Storage in Norway. Finally, we want to thank all UKB participants and facilitators who made this research possible.

## 11 Data Availability Statement

All raw data are available from the UK Biobank (www.ukbiobank.ac.uk). Synthetic datasets of the diffusion MRI data were created with the synthpop R package (Nowok et al., 2016) based on the original data for all six diffusion approaches (resulting in six datasets) which are openly available at the Open Science Framework: (https://osf.io/nv8ea/). Furthermore, a synthetic version of the data used to analyze group differences and associations of brain age with bio-psycho-social factors can be found here: https://osf.io/nk43a/. Synthetic datasets are simulated datasets closely mimicking the statistical characteristics of the original data while protecting data privacy and anonymity. Analysis code can be found on GitHub (https://github.com/MaxKorbmacher/Brain-Age-Variability).

## References

Adler, N. E., & Ostrove, J. M. (1999). Socioeconomic status and health: what we know and what we don’t. Annals of the New York academy of Sciences, 896(1), 3–15. https://doi.org/10.1111/j.1749-6632.1999.tb08101.x

Alfaro-Almagro, F., Jenkinson, M., Bangerter, N. K., Andersson, J. L., Griffanti, L., Douaud, G., … & Smith, S. M. (2018). Image processing and Quality Control for the first 10,000 brain imaging datasets from UK Biobank. Neuroimage, 166, 400–424. https://doi.org/10.1016/j.neuroimage.2017.10.034

Andersson, J. L., & Sotiropoulos, S. N. (2016). An integrated approach to correction for off-resonance effects and subject movement in diffusion MR imaging. Neuroimage, 125, 1063–1078. https://doi.org/10.1016/j.neuroimage.2015.10.019

Bannai, A., & Tamakoshi, A. (2014). The association between long working hours and health: a systematic review of epidemiological evidence. Scandinavian journal of work, environment & health, 5–18. https://doi.org/10.5271/sjweh.3388

Barth, C., & de Lange, A. M. G. (2020). Towards an understanding of women’s brain aging: the immunology of pregnancy and menopause. Frontiers in Neuroendocrinology, 58, 100850. https://doi.org/10.1016/j.yfrne.2020.100850

Beck, D., de Lange, A. M. G., Maximov, I. I., Richard, G., Andreassen, O. A., Nordvik, J. E., & Westlye, L. T. (2021). White matter microstructure across the adult lifespan: A mixed longitudinal and cross-sectional study using advanced diffusion models and brain-age prediction. NeuroImage, 224, 117441. https://doi.org/10.1016/j.neuroimage.2020.117441

Beck, D., de Lange, A. M. G., Alnæs, D., Maximov, I. I., Pedersen, M. L., Leinhard, O. D., … & Westlye, L. T. (2022a). Adipose tissue distribution from body MRI is associated with cross-sectional and longitudinal brain age in adults. NeuroImage: Clinical, 33, 102949. https://doi.org/10.1016/j.nicl.2022.102949

Beck, D., de Lange, A. M. G., Pedersen, M. L., Alnæs, D., Maximov, I. I., Voldsbekk, I., … & Westlye, L. T. (2022b). Cardiometabolic risk factors associated with brain age and accelerate brain ageing. Human brain mapping, 43(2), 700–720. https://doi.org/10.1002/hbm.25680

Bentler, P. M. (1980). Multivariate analysis with latent variables: Causal modeling. Annual review of psychology, 31(1), 419–456.

Biessels, G. J., & Reijmer, Y. D. (2014). Brain changes underlying cognitive dysfunction in diabetes: what can we learn from MRI?. Diabetes, 63(7), 2244–2252. https://doi.org/10.2337/db14-0348

Billiet, T., Vandenbulcke, M., Mädler, B., Peeters, R., Dhollander, T., Zhang, H., … & Emsell, L. (2015). Age-related microstructural differences quantified using myelin water imaging and advanced diffusion MRI. Neurobiology of aging, 36(6), 2107–2121. https://doi.org/10.1016/j.neurobiolaging.2015.02.029

Bollen, K. A. (1989). Structural equations with latent variables. New York: John Wiley & Sons

Cao, J., Wang, J., Dwyer, J. B., Gautier, N. M., Wang, S., Leslie, F. M., & Li, M. D. (2013). Gestational nicotine exposure modifies myelin gene expression in the brains of adolescent rats with sex differences. Translational psychiatry, 3(4), e247–e247. https://doi.org/10.1038/tp.2013.21

Chen, T., & Guestrin, C. (2016, August). Xgboost: A scalable tree boosting system. In Proceedings of the 22nd acm sigkdd international conference on knowledge discovery and data mining (pp. 785–794). https://doi.org/10.1145/2939672.2939785

Chen, C. L., Kuo, M. C., Chen, P. Y., Tung, Y. H., Hsu, Y. C., Huang, C. W. C., … & Tseng, W. Y. I. (2022). Validation of neuroimaging-based brain age gap as a mediator between modifiable risk factors and cognition. Neurobiology of Aging, 114, 61–72. https://doi.org/10.1016/j.neurobiolaging.2022.03.006

Cherbuin, N., Walsh, E. I., Shaw, M., Luders, E., Anstey, K. J., Sachdev, P. S., … & Gaser, C. (2021). Optimal blood pressure keeps our brains younger. Frontiers in Aging Neuroscience, 529. https://doi.org/10.3389/fnagi.2021.694982

Cetin-Karayumak, S., Di Biase, M. A., Chunga, N., Reid, B., Somes, N., Lyall, A. E., … & Kubicki, M. (2020). White matter abnormalities across the lifespan of schizophrenia: a harmonized multi-site diffusion MRI study. Molecular psychiatry, 25(12), 3208–3219. https://doi.org/10.1038/s41380-019-0509-y

Cole, J. H. (2020). Multimodality neuroimaging brain-age in UK biobank: relationship to biomedical, lifestyle, and cognitive factors. Neurobiology of aging, 92, 34–42. https://doi.org/10.1016/j.neurobiolaging.2020.03.014

Cole, J. H., & Franke, K. (2017). Predicting age using neuroimaging: innovative brain ageing biomarkers. Trends in neurosciences, 40(12), 681–690. https://doi.org/10.1016/j.tins.2017.10.001

Cole, J. H., Ritchie, S. J., Bastin, M. E., Hernández, V., Muñoz Maniega, S., Royle, N., … & Deary, I. J. (2018). Brain age predicts mortality. Molecular psychiatry, 23(5), 1385–1392. https://doi.org/10.1038/mp.2017.62

Cox, S. R., Ritchie, S. J., Tucker-Drob, E. M., Liewald, D. C., Hagenaars, S. P., Davies, G., … & Deary, I. J. (2016). Ageing and brain white matter structure in 3,513 UK Biobank participants. Nature communications, 7(1), 1–13. https://doi.org/10.1038/ncomms13629

Crossley, T. F., & Kennedy, S. (2002). The reliability of self-assessed health status. Journal of health economics, 21(4), 643–658. https://doi.org/10.1016/S0167-6296(02)00007-3

de Lange, A. M. G., Anatürk, M., Suri, S., Kaufmann, T., Cole, J. H., Griffanti, L., … & Ebmeier, K. P. (2020a). Multimodal brain-age prediction and cardiovascular risk: The Whitehall II MRI sub-study. NeuroImage, 222, 117292. https://doi.org/10.1016/j.neuroimage.2020.117292

de Lange, A. M. G., Barth, C., Kaufmann, T., Maximov, I. I., van der Meer, D., Agartz, I., & Westlye, L. T. (2020b). Women’s brain aging: Effects of sex-hormone exposure, pregnancies, and genetic risk for Alzheimer’s disease. Human brain mapping, 41(18), 5141–5150. https://doi.org/10.1002/hbm.25180

de Lange, A. M. G., & Cole, J. H. (2020). Commentary: Correction procedures in brain-age prediction. NeuroImage: Clinical, 26. https://doi.org/10.1016%2Fj.nicl.2020.102229

de Lange, A. M. G., Kaufmann, T., Quintana, D. S., Winterton, A., Andreassen, O. A., Westlye, L. T., & Ebmeier, K. P. (2021). Prominent health problems, socioeconomic deprivation, and higher brain age in lonely and isolated individuals: A population-based study. Behavioural Brain Research, 414, 113510. https://doi.org/10.1016/j.bbr.2021.113510

de Lange, A. M. G., Anatürk, M., Rokicki, J., Han, L. K., Franke, K., Alnæs, D., … & Cole, J. H. (2022). Mind the gap: Performance metric evaluation in brain-age prediction. Human Brain Mapping. https://doi.org/10.1002/hbm.25837

Denissen, S., Engemann, D. A., De Cock, A., Costers, L., Baijot, J., Laton, J., … & Nagels, G. (2022). Brain age as a surrogate marker for cognitive performance in multiple sclerosis. European journal of neurology, 29(10), 3039–3049. https://doi.org/10.1111/ene.15473

Distelberg, B. J., Staack, A., Elsen, K. D. D., & Sabaté, J. (2017). The effect of coffee and caffeine on mood, sleep, and health-related quality of life. Journal of Caffeine Research, 7(2), 59–70. https://doi.org/10.1089/jcr.2016.0023

Eikenes, L., Visser, E., Vangberg, T., & Håberg, A. K. (2022). Both brain size and biological sex contribute to variation in white matter microstructure in middle-aged healthy adults. Human Brain Mapping. https://doi.org/10.1002/hbm.26093

Elliott, L. T., Sharp, K., Alfaro-Almagro, F., Shi, S., Miller, K. L., Douaud, G., … & Smith, S. M. (2018). Genome-wide association studies of brain imaging phenotypes in UK Biobank. Nature, 562(7726), 210–216. https://doi.org/10.1038/s41586-018-0571-7

Elliott, M. L., Belsky, D. W., Knodt, A. R., Ireland, D., Melzer, T. R., Poulton, R., … & Hariri, A. R. (2021). Brain-age in midlife is associated with accelerated biological aging and cognitive decline in a longitudinal birth cohort. Molecular psychiatry, 26(8), 3829–3838. https://doi.org/10.1038/s41380-019-0626-7

Engel, G. L. (1977). The need for a new medical model: a challenge for biomedicine. Science, 196(4286), 129–136. https://doi.org/10.1126/science.847460

Erhardt, L. (2009). Cigarette smoking: an undertreated risk factor for cardiovascular disease. Atherosclerosis, 205(1), 23–32. https://doi.org/10.1016/j.atherosclerosis.2009.01.007

Faragher, E. B., Cass, M., & Cooper, C. L. (2013). The relationship between job satisfaction and health: a meta-analysis. From stress to wellbeing Volume 1, 254–271.

Fieremans, E., Jensen, J. H., & Helpern, J. A. (2011). White matter characterization with diffusional kurtosis imaging. Neuroimage, 58(1), 177–188. https://doi.org/10.1016/j.neuroimage.2011.06.006

Kenny, P. J., & Markou, A. (2006). Nicotine self-administration acutely activates brain reward systems and induces a long-lasting increase in reward sensitivity. Neuropsychopharmacology, 31(6), 1203–1211. https://doi.org/10.1038/sj.npp.1300905

Le Foll, B., Piper, M. E., Fowler, C. D., Tonstad, S., Bierut, L., Lu, L., … & Hall, W. D. (2022). Tobacco and nicotine use. Nature Reviews Disease Primers, 8(1), 1–16. https://doi.org/10.1038/s41572-022-00346-w

Fawns-Ritchie, C., & Deary, I. J. (2020). Reliability and validity of the UK Biobank cognitive tests. PloS one, 15(4), e0231627. https://doi.org/10.1371/journal.pone.0231627

Forte, G., De Pascalis, V., Favieri, F., & Casagrande, M. (2019). Effects of blood pressure on cognitive performance: A systematic review. Journal of clinical medicine, 9(1), 34. https://doi.org/10.3390/jcm9010034

Franke, K., Gaser, C., Manor, B., & Novak, V. (2013). Advanced BrainAGE in older adults with type 2 diabetes mellitus. Frontiers in aging neuroscience, 5, 90. https://doi.org/10.3389/fnagi.2013.00090

Ghaemi, S. N. (2009). The rise and fall of the biopsychosocial model. The British Journal of Psychiatry, 195(1), 3–4. https://doi.org/10.1192/bjp.bp.109.063859

Gill, D., Zuber, V., Dawson, J., Pearson-Stuttard, J., Carter, A. R., Sanderson, E., … & Elliott, P. (2021). Risk factors mediating the effect of body mass index and waist-to-hip ratio on cardiovascular outcomes: Mendelian randomization analysis. International Journal of Obesity, 45(7), 1428–1438. https://doi.org/10.1038/s41366-021-00807-4

Gozzi, A., Schwarz, A., Reese, T., Bertani, S., Crestan, V., & Bifone, A. (2006). Region-specific effects of nicotine on brain activity: a pharmacological MRI study in the drug-naive rat. Neuropsychopharmacology, 31(8), 1690–1703. https://doi.org/10.1038/sj.npp.1300955

Gurholt, T. P., Kaufmann, T., Frei, O., Alnæs, D., Haukvik, U. K., van der Meer, D., … & Andreassen, O. A. (2021). Population-based body–brain mapping links brain morphology with anthropometrics and body composition. Translational psychiatry, 11(1), 1–12. https://doi.org/10.1038/s41398-021-01414-7

Hamer, M., & Batty, G. D. (2019). Association of body mass index and waist-to-hip ratio with brain structure: UK Biobank study. Neurology, 92(6), e594–e600. https://doi.org/10.1212/WNL.0000000000006879

Higdon, J. V., & Frei, B. (2006). Coffee and health: a review of recent human research. Critical reviews in food science and nutrition, 46(2), 101–123. https://doi.org/10.1080/10408390500400009

Houenou, J., Wessa, M., Douaud, G., Leboyer, M., Chanraud, S., Perrin, M., … & Paillere-Martinot, M. L. (2007). Increased white matter connectivity in euthymic bipolar patients: diffusion tensor tractography between the subgenual cingulate and the amygdalo-hippocampal complex. Molecular psychiatry, 12(11), 1001–1010. https://doi.org/10.1038/sj.mp.4002010

Hsieh, C. M. (2001). Correlates of financial satisfaction. The International Journal of Aging and Human Development, 52(2), 135–153. https://doi.org/10.2190/9YDE-46PA-MV9C-2JRB

Ingalhalikar, M., Smith, A., Parker, D., Satterthwaite, T. D., Elliott, M. A., Ruparel, K., … & Verma, R. (2014). Sex differences in the structural connectome of the human brain. Proceedings of the National Academy of Sciences, 111(2), 823–828. https://doi.org/10.1073/pnas.1316909110

Jelescu, I. O., Palombo, M., Bagnato, F., & Schilling, K. G. (2020). Challenges for biophysical modeling of microstructure. Journal of Neuroscience Methods, 344, 108861. https://doi.org/10.1016/j.jneumeth.2020.108861

Jenkinson, M., Beckmann, C. F., Behrens, T. E., Woolrich, M. W., & Smith, S. M. (2012). Fsl. Neuroimage, 62(2), 782–790. https://doi.org/10.1016/j.neuroimage.2011.09.015

Jensen, J. H., Helpern, J. A., Ramani, A., Lu, H., & Kaczynski, K. (2005). Diffusional kurtosis imaging: the quantification of non-gaussian water diffusion by means of magnetic resonance imaging. Magnetic Resonance in Medicine: An Official Journal of the International Society for Magnetic Resonance in Medicine, 53(6), 1432–1440. https://doi.org/10.1002/mrm.20508

Jirsaraie, R. J., Kaufmann, T., Bashyam, V., Erus, G., Luby, J. L., Westlye, L. T., … & Sotiras, A. (2022). Benchmarking the generalizability of brain age models: Challenges posed by scanner variance and prediction bias. Human Brain Mapping. https://doi.org/10.1002/hbm.26144

Kaden, E., Kelm, N. D., Carson, R. P., Does, M. D., & Alexander, D. C. (2016a). Multi-compartment microscopic diffusion imaging. NeuroImage, 139, 346–359. https://doi.org/10.1016/j.neuroimage.2016.06.002

Kaden, E., Kruggel, F., & Alexander, D. C. (2016b). Quantitative mapping of the per-axon diffusion coefficients in brain white matter. Magnetic resonance in medicine, 75(4), 1752–1763. https://doi.org/10.1002/mrm.25734

Kahn, S. E., Hull, R. L., & Utzschneider, K. M. (2006). Mechanisms linking obesity to insulin resistance and type 2 diabetes. Nature, 444(7121), 840–846. https://doi.org/10.1038/nature05482

Kamagata, K., Andica, C., Hatano, T., Ogawa, T., Takeshige-Amano, H., Ogaki, K., … & Aoki, S. (2020). Advanced diffusion magnetic resonance imaging in patients with Alzheimer’s and Parkinson’s diseases. Neural regeneration research, 15(9), 1590. https://doi.org/10.4103/1673-5374.276326

Kaufmann, T., van der Meer, D., Doan, N. T., Schwarz, E., Lund, M. J., Agartz, I., … & Westlye, L. T. (2019). Common brain disorders are associated with heritable patterns of apparent aging of the brain. Nature neuroscience, 22(10), 1617–1623. https://doi.org/10.1038/s41593-019-0471-7

Kellner, E., Dhital, B., Kiselev, V. G., & Reisert, M. (2016). Gibbs-ringing artifact removal based on local subvoxel-shifts. Magnetic resonance in medicine, 76(5), 1574–1581. https://doi.org/10.1002/mrm.26054

Korbmacher, M., de Lange, A. M., van der Meer, D., Beck, D., Eikefjord, E. N., Lundervold, A., … & Maximov, I. I. (2022). Brain-wide associations between white matter and age highlight the role of fornix microstructure in brain age. bioRxiv. https://doi.org/10.1101/2022.09.29.510029

Kotsis, V., Stabouli, S., Papakatsika, S., Rizos, Z., & Parati, G. (2010). Mechanisms of obesity-induced hypertension. Hypertension research, 33(5), 386–393. https://doi.org/10.1038/hr.2010.9

Lawrence, K. E., Nabulsi, L., Santhalingam, V., Abaryan, Z., Villalon-Reina, J. E., Nir, T. M., … & Thompson, P. M. (2021). Age and sex effects on advanced white matter microstructure measures in 15,628 older adults: a UK biobank study. Brain imaging and behavior, 15(6), 2813–2823. https://doi.org/10.1007/s11682-021-00548-y

Lehman, B. J., David, D. M., & Gruber, J. A. (2017). Rethinking the biopsychosocial model of health: Understanding health as a dynamic system. Social and personality psychology compass, 11(8), e12328. https://doi.org/10.1111/spc3.12328

Leonardsen, E. H., Peng, H., Kaufmann, T., Agartz, I., Andreassen, O. A., Celius, E. G., … & Wang, Y. (2022). Deep neural networks learn general and clinically relevant representations of the ageing brain. NeuroImage, 256, 119210. https://doi.org/10.1016/j.neuroimage.2022.119210

Liem, F., Varoquaux, G., Kynast, J., Beyer, F., Masouleh, S. K., Huntenburg, J. M., … & Margulies, D. S. (2017). Predicting brain-age from multimodal imaging data captures cognitive impairment. Neuroimage, 148, 179–188. https://doi.org/10.1016/j.neuroimage.2016.11.005

Lim, N., Kim, E. K., Kim, H., Yang, E., & Lee, S. M. (2010). Individual and work-related factors influencing burnout of mental health professionals: A meta-analysis. Journal of Employment Counseling, 47(2), 86–96. https://doi.org/10.1002/j.2161-1920.2010.tb00093.x

Lohner, V., Pehlivan, G., Sanroma, G., Miloschewski, A., Schirmer, M. D., Stöcker, T., … & Breteler, M. M. (2022). Relation Between Sex, Menopause, and White Matter Hyperintensities: The Rhineland Study. Neurology, 99(9), e935–e943. https://doi.org/10.1212/WNL.0000000000200782

Marquand, A. F., Kia, S. M., Zabihi, M., Wolfers, T., Buitelaar, J. K., & Beckmann, C. F. (2019). Conceptualizing mental disorders as deviations from normative functioning. Molecular psychiatry, 24(10), 1415–1424. https://doi.org/10.1038/s41380-019-0441-1

Mathew, B., Francis, L., Kayalar, A., & Cone, J. (2008). Obesity: effects on cardiovascular disease and its diagnosis. The Journal of the American Board of Family Medicine, 21(6), 562–568. https://doi.org/10.3122/jabfm.2008.06.080080

Maximov, I. I., Alnæs, D., & Westlye, L. T. (2019). Towards an optimised processing pipeline for diffusion magnetic resonance imaging data: Effects of artefact corrections on diffusion metrics and their age associations in UK Biobank. Human Brain Mapping, 40(14), 4146–4162. https://doi.org/10.1002/hbm.24691

Maximov, I. I., van Der Meer, D., de Lange, A. M. G., Kaufmann, T., Shadrin, A., Frei, O., … & Westlye, L. T. (2021). Fast qualitY conTrol meThod foR derIved diffUsion Metrics (YTTRIUM) in big data analysis: UK Biobank 18,608 example. Human brain mapping, 42(10), 3141–3155. https://doi.org/10.1002/hbm.25424

Meissner, A. (2016). Hypertension and the brain: a risk factor for more than heart disease. Cerebrovascular diseases, 42(3-4), 255–262. https://doi.org/10.1159/000446082

Miller, K. L., Alfaro-Almagro, F., Bangerter, N. K., Thomas, D. L., Yacoub, E., Xu, J., … & Smith, S. M. (2016). Multimodal population brain imaging in the UK Biobank prospective epidemiological study. Nature neuroscience, 19(11), 1523–1536. https://doi.org/10.1038/nn.4393

Molarius, A., Seidell, J. C., Sans, S., Tuomilehto, J., & Kuulasmaa, K. (1999). Waist and hip circumferences, and waist-hip ratio in 19 populations of the WHO MONICA Project. International journal of obesity and related metabolic disorders : journal of the International Association for the Study of Obesity, 23(2), 116–125. https://doi.org/10.1038/sj.ijo.0800772

Mori, S., Wakana, S., Van Zijl, P. C., & Nagae-Poetscher, L. M. (2005). MRI atlas of human white matter. Elsevier.

Mosconi, L., Berti, V., Dyke, J., Schelbaum, E., Jett, S., Loughlin, L., … & Brinton, R. D. (2021). Menopause impacts human brain structure, connectivity, energy metabolism, and amyloid-beta deposition. Scientific reports, 11(1), 1–16. https://doi.org/10.1038/s41598-021-90084-y

Nehlig, A. (2016). Effects of coffee/caffeine on brain health and disease: What should I tell my patients?. Practical neurology, 16(2), 89–95. http://dx.doi.org/10.1136/practneurol-2015-001162

Ning, K., Zhao, L., Matloff, W., Sun, F., & Toga, A. W. (2020). Association of relative brain age with tobacco smoking, alcohol consumption, and genetic variants. Scientific reports, 10(1), 1–10. https://doi.org/10.1038/s41598-019-56089-4

Niu, X., Zhang, F., Kounios, J., & Liang, H. (2019). Improved prediction of brain age using multimodal neuroimaging data. Human brain mapping, 41(6), 1626–1643. https://doi.org/10.1002/hbm.24899

Novikov, D. S., Fieremans, E., Jespersen, S. N., & Kiselev, V. G. (2019). Quantifying brain microstructure with diffusion MRI: Theory and parameter estimation. NMR in Biomedicine, 32(4), e3998. https://doi.org/10.1002/nbm.3998

Nowok, B., Raab, G. M., & Dibben, C. (2016). synthpop: Bespoke creation of synthetic data in R. Journal of statistical software, 74, 1–26. https://doi.org/10.18637/jss.v074.i11

Pavlakis, A. E., Noble, K., Pavlakis, S. G., Ali, N., & Frank, Y. (2015). Brain imaging and electrophysiology biomarkers: is there a role in poverty and education outcome research?. Pediatric Neurology, 52(4), 383–388. https://doi.org/10.1016/j.pediatrneurol.2014.11.005

Pham, K., Mulugeta, A., Zhou, A., O’Brien, J. T., Llewellyn, D. J., & Hyppönen, E. (2022). High coffee consumption, brain volume and risk of dementia and stroke. Nutritional Neuroscience, 25(10), 2111–2122. https://doi.org/10.1080/1028415X.2021.1945858

Pines, A. R., Cieslak, M., Larsen, B., Baum, G. L., Cook, P. A., Adebimpe, A., … & Satterthwaite, T. D. (2020). Leveraging multi-shell diffusion for studies of brain development in youth and young adulthood. Developmental cognitive neuroscience, 43, 100788. https://doi.org/10.1016/j.dcn.2020.100788

Peng, H., Gong, W., Beckmann, C. F., Vedaldi, A., & Smith, S. M. (2021). Accurate brain age prediction with lightweight deep neural networks. Medical image analysis, 68, 101871. https://doi.org/10.1016/j.media.2020.101871

Reisert, M., Kellner, E., Dhital, B., Hennig, J., & Kiselev, V. G. (2017). Disentangling micro from mesostructure by diffusion MRI: a Bayesian approach. Neuroimage, 147, 964–975. https://doi.org/10.1016/j.neuroimage.2016.09.058

Remiszewski, N., Bryant, J. E., Rutherford, S. E., Marquand, A. F., Nelson, E., Askar, I., … & Kraguljac, N. V. (2022). Contrasting Case-Control and Normative Reference Approaches to Capture Clinically Relevant Structural Brain Abnormalities in Patients With First-Episode Psychosis Who Are Antipsychotic Naive. JAMA psychiatry. https://doi.org/10.1001/jamapsychiatry.2022.3010

Reychav, I., Beeri, R., Balapour, A., Raban, D. R., Sabherwal, R., & Azuri, J. (2019). How reliable are self-assessments using mobile technology in healthcare? The effects of technology identity and self-efficacy. Computers in Human Behavior, 91, 52–61. https://doi.org/10.1016/j.chb.2018.09.024

Rokicki, J., Wolfers, T., Nordhøy, W., Tesli, N., Quintana, D. S., Alnæs, D., … & Westlye, L. T. (2021). Multimodal imaging improves brain age prediction and reveals distinct abnormalities in patients with psychiatric and neurological disorders. Human brain mapping, 42(6), 1714–1726. https://doi.org/10.1002/hbm.25323

Sanford, N., Ge, R., Antoniades, M., Modabbernia, A., Haas, S. S., Whalley, H. C., … & Frangou, S. (2022). Sex differences in predictors and regional patterns of brain age gap estimates. Human Brain Mapping, 43(15), 4689–4698. https://doi.org/10.1002/hbm.25983

Salih, A., Boscolo Galazzo, I., Raisi-Estabragh, Z., Rauseo, E., Gkontra, P., Petersen, S. E., … & Menegaz, G. (2021). Brain age estimation at tract group level and its association with daily life measures, cardiac risk factors and genetic variants. Scientific reports, 11(1), 1–14. https://doi.org/10.1038/s41598-021-99153-8

Shaked, D., Leibel, D. K., Katzel, L. I., Davatzikos, C., Gullapalli, R. P., Seliger, S. L., … & Waldstein, S. R. (2019). Disparities in diffuse cortical white matter integrity between socioeconomic groups. Frontiers in Human Neuroscience, 13, 198. https://doi.org/10.3389/fnhum.2019.00198

Smith, S. M. (2002). Fast robust automated brain extraction. Human brain mapping, 17(3), 143–155. https://doi.org/10.1002/hbm.10062

Smith, S. M., Vidaurre, D., Alfaro-Almagro, F., Nichols, T. E., & Miller, K. L. (2019). Estimation of brain age delta from brain imaging. Neuroimage, 200, 528–539. https://doi.org/10.1016/j.neuroimage.2019.06.017

Smith, S. M., Jenkinson, M., Johansen-Berg, H., Rueckert, D., Nichols, T. E., Mackay, C. E., … & Behrens, T. E. (2006). Tract-based spatial statistics: voxelwise analysis of multi-subject diffusion data. Neuroimage, 31(4), 1487–1505. https://doi.org/10.1016/j.neuroimage.2006.02.024

Smith, S. M., Jenkinson, M., Woolrich, M. W., Beckmann, C. F., Behrens, T. E., Johansen-Berg, H., … & Matthews, P. M. (2004). Advances in functional and structural MR image analysis and implementation as FSL. Neuroimage, 23, S208–S219. https://doi.org/10.1016/j.neuroimage.2004.07.051

Sone, D., Beheshti, I., Shinagawa, S., Niimura, H., Kobayashi, N., Kida, H., … & Shigeta, M. (2022). Neuroimaging-derived brain age is associated with life satisfaction in cognitively unimpaired elderly: A community-based study. Translational psychiatry, 12(1), 1–6. https://doi.org/10.1038/s41398-022-01793-5

Subramaniapillai, S., Suri, S., Barth, C., Maximov, I. I., Voldsbekk, I., van der Meer, D., … & de Lange, A. M. G. (2022). Sex-and age-specific associations between cardiometabolic risk and white matter brain age in the UK Biobank cohort. Human brain mapping, 43(12), 3759–3774. https://doi.org/10.1002/hbm.25882

Sudlow, C., Gallacher, J., Allen, N., Beral, V., Burton, P., Danesh, J., … & Collins, R. (2015). UK biobank: an open access resource for identifying the causes of a wide range of complex diseases of middle and old age. PLoS medicine, 12(3), e1001779. https://doi.org/10.1371/journal.pmed.1001779

van der Harten, B., de Leeuw, F. E., Weinstein, H. C., Scheltens, P., & Biessels, G. J. (2006). Brain imaging in patients with diabetes: a systematic review. Diabetes care, 29(11), 2539–2548. https://doi.org/10.2337/dc06-1637

Veraart, J., Fieremans, E., & Novikov, D. S. (2016). Diffusion MRI noise mapping using random matrix theory. Magnetic resonance in medicine, 76(5), 1582–1593. https://doi.org/10.1002/mrm.26059

Vidal-Pineiro, D., Wang, Y., Krogsrud, S. K., Amlien, I. K., Baaré, W. F., Bartres-Faz, D., … & Fjell, A. (2021). Individual variations in ‘brain age‘relate to early-life factors more than to longitudinal brain change. Elife, 10, e69995. https://doi.org/10.7554/eLife.69995

Wade, D. T., & Halligan, P. W. (2017). The biopsychosocial model of illness: a model whose time has come. Clinical rehabilitation, 31(8), 995–1004. https://doi.org/10.1177/0269215517709890

Wang, Z., Bovik, A. C., Sheikh, H. R., & Simoncelli, E. P. (2004). Image quality assessment: from error visibility to structural similarity. IEEE transactions on image processing, 13(4), 600–612. https://doi.org/10.1109/TIP.2003.819861

Westlye, L. T., Walhovd, K. B., Dale, A. M., Bjørnerud, A., Due-Tønnessen, P., Engvig, A., … & Fjell, A. M. (2010). Lifespan changes of the human brain white matter: diffusion tensor imaging (DTI) and volumetry. Cerebral cortex, 20(9), 2055–2068. https://doi.org/10.1093/cercor/bhp280

Westreich, D., & Greenland, S. (2013). The table 2 fallacy: presenting and interpreting confounder and modifier coefficients. American journal of epidemiology, 177(4), 292–298. https://doi.org/10.1093/aje/kws412

Wrighten, S. A., Piroli, G. G., Grillo, C. A., & Reagan, L. P. (2009). A look inside the diabetic brain: Contributors to diabetes-induced brain aging. Biochimica et Biophysica Acta (BBA)-Molecular Basis of Disease, 1792(5), 444–453. https://doi.org/10.1016/j.bbadis.2008.10.013

Wood, D. A., Kafiabadi, S., Al Busaidi, A., Guilhem, E., Montvila, A., Lynch, J., … & Booth, T. C. (2022). Accurate brain-age models for routine clinical MRI examinations. NeuroImage, 249, 118871. https://doi.org/10.1016/j.neuroimage.2022.118871

Yap, Q. J., Teh, I., Fusar-Poli, P., Sum, M. Y., Kuswanto, C., & Sim, K. (2013). Tracking cerebral white matter changes across the lifespan: insights from diffusion tensor imaging studies. Journal of neural transmission, 120, 1369–1395. https://doi.org/10.1007/s00702-013-0971-7

Zhao, B., Zhang, J., Ibrahim, J. G., Luo, T., Santelli, R. C., Li, Y., … & Zhu, H. (2021). Large-scale GWAS reveals genetic architecture of brain white matter microstructure and genetic overlap with cognitive and mental health traits (n= 17,706). Molecular psychiatry, 26(8), 3943–3955. https://doi.org/10.1038/s41380-019-0569-z

