## Supplement for "Bio-psycho-social factors’ associations with brain age: a large-scale UK Biobank diffusion study of 35,749 participants"

### 1 Supplementary Figures and Tables

#### 1.1 Supplementary Tables

**Supplementary Table 1.** Overview of diffusion metrics by diffusion approach

| **Diffusion Approach** | **Metrics** |
| --- | --- |
| Bayesian Rotationally Invariant Approach (BRIA) | intra-axonal axial diffusivity (DAX intra) |
|  | extra-axonal radial diffusivity (DRAD extra) |
|  | microscopic fractional anisotropy (micro FA) |
|  | extra-axonal axial diffusivity (DAX extra) |
|  | intra-axonal water fraction (V intra) |
|  | extra-axonal water fraction (V extra) |
|  | cerebrospinal fluid fraction (vCSF) |
|  | microscopical axial diffusivity (micro AX) |
|  | microscopic radial diffusivity (micro RD) |
|  | microscopical apparent diffusion coefficient (micro ADC) |
| Diffusion Kurtosis Imaging (DKI) | mean kurtosis (MK) |
|  | radial kurtosis (RK) |
|  | axial kurtosis (AK) |
| Diffusion Tensor Imaging (DTI) | fractional anisotropy (FA) |
|  | axial diffusivity (AD) |
|  | mean diffusivity (MD) |
|  | radial diffusivity (RD) |
| Spherical Mean Technique (SMT) | fractional anisotropy (SMT FA) |
|  | mean diffusivity (SMT md) |
|  | transverse diffusion coefficient (SMT trans) |
|  | longitudinal diffusion coefficient (SMT long) |
| Multi-compartment Spherical Mean Technique (mcSMT) | extra-neurite microscopic mean diffusivity (mcSMT extra md) |
|  | extra-neurite transverse microscopic diffusivity (mcSMT extra trans) |
|  | mc SMTdiffusion coefficient (mcSMT diff) |
|  | intra-neurite volume fraction (mcSMT intra) |
| White Matter Tract Integrity (WMTI) | axonal water fraction (AWF) |
|  | radial extra-axonal diffusivity (radEAD) |
|  | axial extra-axonal diffusivity (axEAD) |

**Supplementary Table 2.** White matter tracts and regions

| **Regions of Interest** | | | |
| --- | --- | --- | --- |
| **#** | **Label** | **#** | **Label** |
| 1 | middle cerebellar peduncle | 25 | superior corona radiata r |
| 2 | pontine crossing tract | 26 | superior corona radiata l |
| 3 | genu of corpus callosum | 27 | posterior corona radiata r |
| 4 | body of corpus callosum | 28 | posterior corona radiata l |
| 5 | splenium of corpus callosum | 29 | posterior thalamic radiation (include optic radiation) R |
| 6 | fornix (column and body of fornix) | 30 | posterior thalamic radiation (include optic radiation) L |
| 7 | corticospinal tract r | 31 | sagittal stratum r |
| 8 | corticospinal tract l | 32 | sagittal stratum l |
| 9 | medial lemniscus r | 33 | external capsule r |
| 10 | medial lemniscus l | 34 | external capsule l |
| 11 | inferior cerebellar peduncle r | 35 | cingulum (cingulate gyrus) R |
| 12 | inferior cerebellar peduncle l | 36 | cingulum (cingulate gyrus) L |
| 13 | superior cerebellar peduncle r | 37 | cingulum (hippocampus) R |
| 14 | superior cerebellar peduncle l | 38 | cingulum (hippocampus) L |
| 15 | cerebral peduncle r | 39 | fornix r |
| 16 | cerebral peduncle l | 40 | fornix l |
| 17 | anterior limb of internal capsule r | 41 | superior longitudinal fasciculus r |
| 18 | anterior limb of internal capsule l | 42 | superior longitudinal fasciculus l |
| 19 | posterior limb of internal capsule r | 43 | superior fronto-occipital fasciculus |
| 20 | posterior limb of internal capsule l | 44 | superior fronto-occipital fasciculus |
| 21 | retrolenticular part of internal capsule r | 45 | uncinate fasciculus r |
| 22 | retrolenticular part of internal capsule l | 46 | uncinate fasciculus l |
| 23 | anterior corona radiata r | 47 | tapetum r |
| 24 | anterior corona radiata l | 48 | tapetum l |
| **Tracts** | | | |
| **#** | **Tract Name** | **#** | **Tract Name** |
| 1 | Anterior thalamic radiation l | 11 | Inferior fronto-occipital fasciculus l |
| 2 | Anterior thalamic radiation r | 12 | Inferior fronto-occipital fasciculus r |
| 3 | Corticospinal tracts l | 13 | Inferior longitudinal fasciculus l |
| 4 | Corticospinal tracts r | 14 | Inferior longitudinal fasciculus r |
| 5 | Cingulum - cingulate gyrus l | 15 | Superior longitudinal fasciculus l |
| 6 | Cingulum - cingulate gyrus r | 16 | Superior longitudinal fasciculus r |
| 7 | Cingulum – hippocampus l | 17 | Uncinate fasciculus l |
| 8 | Cingulum – hippocampus r | 18 | Uncinate fasciculus r |
| 9 | Forceps major | 19 | Superior longitudinal fasciculus - temporal part l |
| 10 | Forceps minor | 20 | Superior longitudinal fasciculus - temporal part r |

**Supplementary Table 3.** Overview of the utilized bio-psycho-social variables

| **Variable Name** | **UKB Fied Code(s)** | **Processing** |
| --- | --- | --- |
| Age | 33, 53 | Differences between birth date and day of visit at the scanner site |
| Sex | 31 |  |
| Ethnicity | 20115 | Dummy coded as European and non-European |
| Income | 738 | Coded as numeric in regression model from lowest to highest income groups. |
| Education | 845 | Dummy coded as obtained higher education and did not obtain higher education |
| Number of social visits | 1031 |  |
| BMI | 21001 |  |
| WHR | 48, 49 | Calculate WHR from waist and hip circumference |
| Pulse pressure | 4079, 94, 4080, 93 | Average of manual and automated systolic and diastolic readings subtracted from each other |
| Diabetes | 2443 |  |
| Smoking | 20116 | Dummy coded to currently smoking or not |
| High Cholesterol | 26037 | Dummy coded as cholesterol levels of 240 or higher being high |
| Diagnosed vascular disease | 6150 | Dummy coded as any or none |
| Birth weight | 20022 |  |
| Sleeping hours | 1160 |  |
| Daily coffee intake | 1498 |  |
| Prospective Memory | 20018 |  |
| Fluid Intelligence | 20016 |  |
| Symbol Digit Substitution | 20245 |  |
| Matrix Puzzles Solved | 6373 |  |
| Tower Arranging | 21004 |  |
| Mean Correct pair matches | 397, 398 | Average of both rounds |
| Digit Memorization | 4259 |  |
| Financial Satisfaction | 4581 |  |
| Job Satisfaction | 4537 |  |
| Health Satisfaction | 4548 |  |
| Self-Rated Health | 2178 |  |
| Relationship Satisfaction | 4570 |  |
| Family Relationship Satisfaction | 4559 |  |
| Happiness | 4526 |  |

**Supplementary Table 4.** Sample characteristics with variables grouped by ethnicity (European vs non-European)

| **Variable** | **Level or Metric** | **European** | **Non-European** | **N_European_** | **N_non-Eur_** | **Test** | **Test Statistic** | ***p*** | **Effect Size** |
| --- | --- | --- | --- | --- | --- | --- | --- | --- | --- |
| Ethnicity | % | 0.968 | 0.0297 | 31160 | 956 |  |  |  |  |
| **Demographics** | | | | | | | | | |
| Scanner Site | % Cheadle | 0.558 | 0.0163 | 17964 | 524 | CS | 355.11 | <.001 |  |
|  | % Newcastle | 0.2608 | 0.0028 | 8392 | 89 |  |  |  |  |
|  | % Reading | 0.1481 | 0.0106 | 4765 | 342 |  |  |  |  |
|  | % Bristol | 0.0012 | 3x10^-05^ | 39 | 1 |  |  |  |  |
| Sex | % Male | 14656 | 469 | 0.456 | 0.015 | CS | 1.45 | .229 |  |
|  | % Female | 16504 | 487 | 0.513 | 0.015 | MW | 18598765 | <.001 | 0.0732 |
| Age | Mean ± SD | 64.6±7.59 | 61.2±7.78 | 31160 | 956 |  |  |  |  |
| Brain age | Mean ± SD | 64.5±5.94 | 62.3±5.88 | 31160 | 956 | MW | 18140728 | <.001 | 0.0642 |
| **Socio-demographics** | | | | | | | | | |
| Income5 | % less £18k | 0.0989 | 0.0038 | 3182 | 121 | CS | 35.853 | <.001 |  |
|  | % £18k-£30 | 0.2348 | 0.0057 | 7553 | 182 |  |  |  |  |
|  | % £30k-£52k | 0.2678 | 0.0069 | 8615 | 223 |  |  |  |  |
|  | % £52k-100k | 0.2068 | 0.0007 | 6653 | 232 |  |  |  |  |
|  | % > £100k | 0.0661 | 0.0002 | 2127 | 91 |  |  |  |  |
|  | Don’t know | 0.0067 | 0.0004 | 217 | 14 |  |  |  |  |
|  | Prefer not to say | 0.0874 | 0.0029 | 2813 | 93 |  |  |  |  |
| Higher education | % Yes | 0.474 | 0.018 | 15253 | 589 | CS | 58.973 | <.001 |  |
|  | % No | 0.494 | 0.011 | 15904 | 367 |  |  |  |  |
| **Cognitive test scores** | | | | | | | | | |
| Matrix puzzles solved | Mean ± SD | 8.04±2.11 | 6.94±2.41 | 21098 | 612 | MW | 8139553 | <.001 | 0.0756 |
| Tower rearranging correct attempts | Mean ± SD | 9.97±3.21 | 8.31±3.45 | 20946 | 598 | MW | 7982748 | <.001 | 0.0785 |
| Prospective memory | Mean ± SD | 1.07±0.39 | 1.11±0.60 | 29393 | 855 | MW | 12014138 | <.001 | 0.0197 |
| Fluid intelligence | Mean ± SD | 6.67±2.04 | 5.19±2.03 | 28902 | 815 | MW | 16422576 | <.001 | 0.113 |
| Digits remembered | Mean ± SD | 6.68±1.53 | 6.33±1.98 | 22362 | 663 | MW | 8208465 | <.001 | 0.0321 |
| Mean number of incorrect pair matches across trials | Mean ± SD | 2.21±1.27 | 2.31±1.44 | 20198 | 541 | MW | 5268132 | .153 | 0.0099 |
| **Life satisfaction** | | | | | | | | | |
| Job satisfaction | Mean ± SD | 4.51±0.86 | 4.45±0.89 | 17611 | 758 | MW | 6921274 | .063 | 0.0137 |
| Financial satisfaction | Mean ± SD | 4.72±0.82 | 4.43±0.89 | 30918 | 934 | MW | 17125664 | <.001 | 0.0587 |
| Health satisfaction | Mean ± SD | 4.47±0.76 | 4.36±0.83 | 30917 | 937 | MW | 155766270 | <.001 | 0.0241 |
| Overall health rating | Mean ± SD | 3.03±0.63 | 2.90±0.68 | 30939 | 938 | MW | 16019589 | <.001 | 0.0358 |
| Family relation satisfaction | Mean ± SD | 4.82±0.84 | 4.60±0.92 | 30759 | 923 | MW | 16102889 | <.001 | 0.0422 |
| Friend relationship satisfaction | Mean ± SD | 4.79±0.72 | 4.64±0.75 | 30667 | 917 | MW | 15513145 | <.001 | 0.0333 |
| Happiness | Mean ± SD | 4.55±0.69 | 4.42±0.71 | 30897 | 931 | MW | 15677286 | <.001 | 0.0291 |
| **Health and lifestyle factors** | | | | | | | | | |
| BMI | Mean ± SD | 26.3±4.26 | 26.4±4.38 | 30083 | 912 | MW | 13474126 | .36 | 0.0052 |
| Pulse pressure | Mean ± SD | 60.1±14.5 | 56.4±14.1 | 27327 | 806 | MW | 12721786 | <.001 | 0.0448 |
| WHR | Mean ± SD | 0.871±0.0878 | 0.875±0.0823 | 30167 | 914 | MW | 13433908 | .187 | 0.0075 |
| Smoking | % Yes | 0.0249 | 0.0011 | 800 | 34 | CS | 3.3461 | .067 |  |
|  | % No | 0.935 | 0.0282 | 30073 | 907 |  |  |  |  |
| Diabetes | % Yes | 0.015 | 0.0016 | 491 | 52 | CS | 80.992 | <.001 |  |
|  | % No | 0.953 | 0.0281 | 305669 | 904 |  |  |  |  |
| Hypertension | % Yes | 0.19 | 0.006 | 6115 | 206 | CS | 2.0511 | .152 |  |
|  | % No | 0.778 | 0.023 | 25045 | 750 |  |  |  |  |
| High Cholesterol | % Yes | 0.118 | 0.002 | 3808 | 113 | CS | 0.1041 | .747 |  |
|  | % No | 0.850 | 0.004 | 27352 | 843 |  |  |  |  |
| Vascular Diagnosis | % Yes | 0.220 | 0.007 | 7078 | 219 | CS | 0.0102 | .920 |  |
|  | % No | 0.748 | 0.022 | 24082 | 737 |  |  |  |  |
| Birth weight (in kg) | Mean ± SD | 3.36±0.616 | 3.19±0.709 | 18986 | 399 | MW | 4477581 | <.001 | 0.0448 |
| Daily coffee intake (cups) | Mean ± SD | 2.08±1.82 | 1.44±1.52 | 30973 | 943 | MW | 18057209 | <.001 | 0.0709 |

Note: MW indicates Man-Whitney tests, CS indicates Chi-squared tests.

**Supplementary Table 5.** Model metrics for linear mixed effect models explaining brain age from covariates (baseline model) and principal components of bio-psycho-social factors

| **R^2^_M_** | **R^2^_C_** | **AIC** | **σ^2^** | **logLik** | **Regression Model** | **Diffusion Model** |
| --- | --- | --- | --- | --- | --- | --- |
| 0.63 | 0.64 | 89380.68 | 12.17 | -44684.34 | Baseline Health Model | FULL |
| 0.37 | 0.38 | 89980.55 | 12.61 | -44984.28 | Baseline Health Model | MEAN |
| 0.52 | 0.53 | 81081.70 | 12.45 | -40534.85 | Baseline Health Model | BRIA |
| 0.55 | 0.56 | 80942.78 | 12.34 | -40465.39 | Baseline Health Model | DKI |
| 0.56 | 0.56 | 81672.27 | 12.95 | -40830.13 | Baseline Health Model | DTI |
| 0.51 | 0.51 | 81832.93 | 13.09 | -40910.47 | Baseline Health Model | SMT |
| 0.49 | 0.50 | 81314.76 | 12.65 | -40651.38 | Baseline Health Model | SMT_mc |
| 0.56 | 0.56 | 81583.93 | 12.87 | -40785.97 | Baseline Health Model | WMTI |
| 0.64 | 0.64 | 89230.94 | 12.06 | -44608.47 | Principal Health Component | FULL |
| 0.38 | 0.38 | 89884.26 | 12.54 | -44935.13 | Principal Health Component | MEAN |
| 0.53 | 0.54 | 80800.87 | 12.22 | -40393.43 | Principal Health Component | BRIA |
| 0.56 | 0.56 | 80823.77 | 12.24 | -40404.88 | Principal Health Component | DKI |
| 0.57 | 0.57 | 81495.51 | 12.80 | -40740.76 | Principal Health Component | DTI |
| 0.51 | 0.52 | 81616.47 | 12.90 | -40801.24 | Principal Health Component | SMT |
| 0.50 | 0.50 | 81083.27 | 12.45 | -40534.63 | Principal Health Component | SMT_mc |
| 0.57 | 0.57 | 81402.56 | 12.71 | -40694.28 | Principal Health Component | WMTI |
| 0.65 | 0.65 | 69957.11 | 12.01 | -34972.56 | Baseline Cognitive Model | FULL |
| 0.40 | 0.40 | 69888.30 | 11.95 | -34938.15 | Baseline Cognitive Model | MEAN |
| 0.55 | 0.56 | 63295.54 | 12.26 | -31641.77 | Baseline Cognitive Model | BRIA |
| 0.57 | 0.57 | 63193.20 | 12.16 | -31590.60 | Baseline Cognitive Model | DKI |
| 0.58 | 0.58 | 63683.61 | 12.67 | -31835.81 | Baseline Cognitive Model | DTI |
| 0.53 | 0.53 | 63802.48 | 12.80 | -31895.24 | Baseline Cognitive Model | SMT |
| 0.52 | 0.52 | 63682.63 | 12.67 | -31835.32 | Baseline Cognitive Model | SMT_mc |
| 0.58 | 0.58 | 63627.19 | 12.61 | -31807.59 | Baseline Cognitive Model | WMTI |
| 0.65 | 0.65 | 69900.08 | 11.95 | -34943.04 | Principal Cognitive Component | FULL |
| 0.40 | 0.40 | 69853.34 | 11.91 | -34919.67 | Principal Cognitive Component | MEAN |
| 0.56 | 0.56 | 63231.97 | 12.19 | -31608.98 | Principal Cognitive Component | BRIA |
| 0.58 | 0.58 | 63167.35 | 12.13 | -31576.68 | Principal Cognitive Component | DKI |
| 0.58 | 0.58 | 63632.15 | 12.61 | -31809.08 | Principal Cognitive Component | DTI |
| 0.53 | 0.53 | 63765.58 | 12.75 | -31875.79 | Principal Cognitive Component | SMT |
| 0.52 | 0.52 | 63631.53 | 12.61 | -31808.77 | Principal Cognitive Component | SMT_mc |
| 0.58 | 0.58 | 63570.60 | 12.54 | -31778.30 | Principal Cognitive Component | WMTI |
| 0.64 | 0.64 | 96494.06 | 12.12 | -48241.03 | Baseline Satisfaction Model | FULL |
| 0.39 | 0.39 | 96617.52 | 12.20 | -48302.76 | Baseline Satisfaction Model | MEAN |
| 0.54 | 0.54 | 87030.99 | 12.18 | -43509.49 | Baseline Satisfaction Model | BRIA |
| 0.56 | 0.57 | 86910.71 | 12.10 | -43449.36 | Baseline Satisfaction Model | DKI |
| 0.57 | 0.57 | 87740.25 | 12.73 | -43864.12 | Baseline Satisfaction Model | DTI |
| 0.52 | 0.52 | 87940.12 | 12.88 | -43964.06 | Baseline Satisfaction Model | SMT |
| 0.51 | 0.51 | 87256.18 | 12.36 | -43622.09 | Baseline Satisfaction Model | SMT_mc |
| 0.57 | 0.57 | 87692.55 | 12.69 | -43840.28 | Baseline Satisfaction Model | WMTI |
| 0.64 | 0.64 | 96484.40 | 12.11 | -48235.20 | Principal Satisfaction Component | FULL |
| 0.39 | 0.39 | 96599.98 | 12.19 | -48292.99 | Principal Satisfaction Component | MEAN |
| 0.54 | 0.54 | 87003.42 | 12.16 | -43494.71 | Principal Satisfaction Component | BRIA |
| 0.56 | 0.57 | 86901.99 | 12.08 | -43444.00 | Principal Satisfaction Component | DKI |
| 0.57 | 0.57 | 87717.59 | 12.71 | -43851.80 | Principal Satisfaction Component | DTI |
| 0.52 | 0.52 | 87908.04 | 12.85 | -43947.02 | Principal Satisfaction Component | SMT |
| 0.51 | 0.51 | 87231.82 | 12.33 | -43608.91 | Principal Satisfaction Component | SMT_mc |
| 0.57 | 0.57 | 87671.73 | 12.67 | -43828.86 | Principal Satisfaction Component | WMTI |

R^2^_M_ refers to marginal variance explained, R^2^_C_ refers to conditional variance explained. The cognitive component explained 33.14%, the health risk component 25.08% and the life-satisfaction component 41.95% of the variance of the respective specific domain variables. Due to missingness in the data, the cognitive model was run on N = 13,134, the health risk factor model on N = 16,740, and the life-satisfaction model on N = 18,087.

**Supplementary Table 6.** Model comparisons of linear mixed effect models explaining brain age from covariates (baseline model) and principal components of bio-psycho-social factors

| **AIC** | **BIC** | **logLik** | **Deviance** | **χ^2^** | ***p*-value** | **Regression Model** | **Diffusion Model** |
| --- | --- | --- | --- | --- | --- | --- | --- |
| 89358.44 | 89404.79 | -44673.22 | 89346.44 |  |  | Health Risk Factors | FULL |
| 89203.14 | 89257.22 | -44594.57 | 89189.14 | 157.29 | <.001 | Health Risk Factors | FULL |
| 89957.51 | 90003.86 | -44972.76 | 89945.51 |  |  | Health Risk Factors | MEAN |
| 89855.56 | 89909.64 | -44920.78 | 89841.56 | 103.95 | <.001 | Health Risk Factors | MEAN |
| 81058.38 | 81104.12 | -40523.19 | 81046.38 |  |  | Health Risk Factors | BRIA |
| 80772.70 | 80826.07 | -40379.35 | 80758.70 | 287.68 | <.001 | Health Risk Factors | BRIA |
| 80919.84 | 80965.59 | -40453.92 | 80907.84 |  |  | Health Risk Factors | DKI |
| 80795.30 | 80848.67 | -40390.65 | 80781.30 | 126.55 | <.001 | Health Risk Factors | DKI |
| 81647.28 | 81693.02 | -40817.64 | 81635.28 |  |  | Health Risk Factors | DTI |
| 81465.00 | 81518.37 | -40725.50 | 81451.00 | 184.28 | <.001 | Health Risk Factors | DTI |
| 81810.16 | 81855.91 | -40899.08 | 81798.16 |  |  | Health Risk Factors | SMT |
| 81588.61 | 81641.98 | -40787.31 | 81574.61 | 223.55 | <.001 | Health Risk Factors | SMT |
| 81290.91 | 81336.65 | -40639.45 | 81278.91 |  |  | Health Risk Factors | SMT_mc |
| 81054.72 | 81108.09 | -40520.36 | 81040.72 | 238.19 | <.001 | Health Risk Factors | SMT_mc |
| 81560.89 | 81606.63 | -40774.44 | 81548.89 |  |  | Health Risk Factors | WMTI |
| 81374.38 | 81427.75 | -40680.19 | 81360.38 | 188.50 | <.001 | Health Risk Factors | WMTI |
| 69934.27 | 69979.16 | -34961.13 | 69922.27 |  |  | Cognitive Factors | FULL |
| 69871.39 | 69923.77 | -34928.70 | 69857.39 | 64.88 | <.001 | Cognitive Factors | FULL |
| 69863.66 | 69908.56 | -34925.83 | 69851.66 |  |  | Cognitive Factors | MEAN |
| 69822.55 | 69874.93 | -34904.28 | 69808.55 | 43.10 | <.001 | Cognitive Factors | MEAN |
| 63273.57 | 63317.85 | -31630.79 | 63261.57 |  |  | Cognitive Factors | BRIA |
| 63204.31 | 63255.96 | -31595.15 | 63190.31 | 71.27 | <.001 | Cognitive Factors | BRIA |
| 63168.11 | 63212.39 | -31578.06 | 63156.11 |  |  | Cognitive Factors | DKI |
| 63136.42 | 63188.07 | -31561.21 | 63122.42 | 33.70 | <.001 | Cognitive Factors | DKI |
| 63660.36 | 63704.63 | -31824.18 | 63648.36 |  |  | Cognitive Factors | DTI |
| 63603.15 | 63654.80 | -31794.58 | 63589.15 | 59.21 | <.001 | Cognitive Factors | DTI |
| 63780.08 | 63824.35 | -31884.04 | 63768.08 |  |  | Cognitive Factors | SMT |
| 63737.56 | 63789.21 | -31861.78 | 63723.56 | 44.52 | <.001 | Cognitive Factors | SMT |
| 63660.26 | 63704.54 | -31824.13 | 63648.26 |  |  | Cognitive Factors | SMT_mc |
| 63603.52 | 63655.17 | -31794.76 | 63589.52 | 58.75 | <.001 | Cognitive Factors | SMT_mc |
| 63603.85 | 63648.12 | -31795.92 | 63591.85 |  |  | Cognitive Factors | WMTI |
| 63541.70 | 63593.35 | -31763.85 | 63527.70 | 64.15 | <.001 | Cognitive Factors | WMTI |
| 96471.25 | 96518.07 | -48229.63 | 96459.25 |  |  | Life Satisfaction Factors | FULL |
| 96455.05 | 96509.67 | -48220.53 | 96441.05 | 18.20 | <.001 | Life Satisfaction Factors | FULL |
| 96593.87 | 96640.69 | -48290.94 | 96581.87 |  |  | Life Satisfaction Factors | MEAN |
| 96569.82 | 96624.44 | -48277.91 | 96555.82 | 26.06 | <.001 | Life Satisfaction Factors | MEAN |
| 87007.91 | 87054.10 | -43497.96 | 86995.91 |  |  | Life Satisfaction Factors | BRIA |
| 86973.91 | 87027.81 | -43479.96 | 86959.91 | 36.00 | <.001 | Life Satisfaction Factors | BRIA |
| 86887.28 | 86933.47 | -43437.64 | 86875.28 |  |  | Life Satisfaction Factors | DKI |
| 86872.13 | 86926.02 | -43429.06 | 86858.13 | 17.15 | <.001 | Life Satisfaction Factors | DKI |
| 87715.16 | 87761.35 | -43851.58 | 87703.16 |  |  | Life Satisfaction Factors | DTI |
| 87686.05 | 87739.94 | -43836.02 | 87672.05 | 31.11 | <.001 | Life Satisfaction Factors | DTI |
| 87917.13 | 87963.33 | -43952.57 | 87905.13 |  |  | Life Satisfaction Factors | SMT |
| 87878.66 | 87932.55 | -43932.33 | 87864.66 | 40.47 | <.001 | Life Satisfaction Factors | SMT |
| 87232.61 | 87278.80 | -43610.31 | 87220.61 |  |  | Life Satisfaction Factors | SMT_mc |
| 87201.84 | 87255.73 | -43593.92 | 87187.84 | 32.78 | <.001 | Life Satisfaction Factors | SMT_mc |
| 87669.36 | 87715.55 | -43828.68 | 87657.36 |  |  | Life Satisfaction Factors | WMTI |
| 87642.13 | 87696.02 | -43814.07 | 87628.13 | 29.23 | <.001 | Life Satisfaction Factors | WMTI |

**Supplementary Table 7.** Model metrics for linear mixed effect models explaining brain age

| **Null model 1: random effect only (scanner site)** | | | | | |
| --- | --- | --- | --- | --- | --- |
| Approach^§^ | Marginal R^2^ | Conditional R^2^ | AIC | σ^2^ | log-Likelihood |
| Full | 0.00 | 0.05 | 205109.61 | 34.34 | -102551.80 |
| Mean | 0.00 | 0.01 | 205727.02 | 35.01 | -102860.51 |
| BRIA | 0.00 | 0.00 | 176485.61 | 25.82 | -88239.81 |
| DKI | 0.00 | 0.01 | 176125.82 | 25.50 | -88059.91 |
| DTI | 0.00 | 0.01 | 175721.29 | 25.14 | -87857.65 |
| SMT | 0.00 | 0.01 | 178437.09 | 27.61 | -89215.55 |
| mcSMT | 0.00 | 0.00 | 178236.46 | 27.43 | -89115.23 |
| WMTI | 0.00 | 0.01 | 175204.13 | 24.70 | -87599.06 |
| **Null model 2: age** | | | | | |
| Approach^§^ | Marginal R^2^ | Conditional R^2^ | AIC | σ^2^ | log-Likelihood |
| Full | 0.02 | 0.07 | 204405.54 | 33.59 | -102198.77 |
| Mean | 0.00 | 0.01 | 205730.16 | 35.01 | -102861.08 |
| BRIA | 0.00 | 0.01 | 176390.44 | 25.73 | -88191.22 |
| DKI | 0.01 | 0.02 | 175823.07 | 25.23 | -87907.53 |
| DTI | 0.01 | 0.02 | 175510.89 | 24.96 | -87751.45 |
| SMT | 0.00 | 0.01 | 178357.89 | 27.53 | -89174.94 |
| mcSMT | 0.00 | 0.00 | 178180.66 | 27.37 | -89086.33 |
| WMTI | 0.00 | 0.01 | 175103.60 | 24.61 | -87547.80 |
| **Baseline: sex and age** | | | | | |
| Approach^§^ | Marginal R^2^ | Conditional R^2^ | AIC | σ^2^ | log-Likelihood |
| Full | 0.65 | 0.65 | 171684.22 | 12.14 | 0.65 |
| Mean | 0.64 | 0.65 | 172828.97 | 12.58 | 0.64 |
| BRIA | 0.52 | 0.52 | 155275.00 | 12.41 | 0.52 |
| DKI | 0.52 | 0.53 | 154853.72 | 12.23 | 0.52 |
| DTI | 0.49 | 0.49 | 156431.15 | 12.91 | 0.49 |
| SMT | 0.53 | 0.53 | 156756.87 | 13.06 | 0.53 |
| mcSMT | 0.54 | 0.54 | 155738.67 | 12.61 | 0.54 |
| WMTI | 0.49 | 0.49 | 156182.66 | 12.80 | 0.49 |
| **Social factors model** | | | | | |
| Approach^§^ | Marginal R^2^ | Conditional R^2^ | AIC | σ^2^ | log-Likelihood |
| Full | 0.65 | 0.65 | 155552.26 | 12.12 | -77766.13 |
| Mean | 0.65 | 0.65 | 156558.06 | 12.55 | -78269.03 |
| BRIA | 0.52 | 0.52 | 140834.07 | 12.39 | -70407.03 |
| DKI | 0.52 | 0.53 | 140414.59 | 12.19 | -70197.30 |
| DTI | 0.49 | 0.49 | 141884.51 | 12.89 | -70932.26 |
| SMT | 0.53 | 0.54 | 142118.11 | 13.01 | -71049.06 |
| mcSMT | 0.54 | 0.54 | 141187.45 | 12.55 | -70583.72 |
| WMTI | 0.49 | 0.49 | 141592.31 | 12.75 | -70786.16 |
| **Cognitive factors model** | | | | | |
| Approach^§^ | Marginal R^2^ | Conditional R^2^ | AIC | σ^2^ | log-Likelihood |
| Full | 0.65 | 0.65 | 69913.91 | 11.92 | -34943.96 |
| Mean | 0.65 | 0.65 | 69838.38 | 11.86 | -34906.19 |
| BRIA | 0.51 | 0.51 | 63234.80 | 12.15 | -31604.40 |
| DKI | 0.51 | 0.51 | 63192.51 | 12.11 | -31583.26 |
| DTI | 0.48 | 0.49 | 63640.14 | 12.57 | -31807.07 |
| SMT | 0.52 | 0.53 | 63778.38 | 12.72 | -31876.19 |
| mcSMT | 0.52 | 0.53 | 63647.84 | 12.58 | -31810.92 |
| WMTI | 0.48 | 0.48 | 63579.72 | 12.51 | -31776.86 |
| **Life satisfaction model** | | | | | |
| Approach^§^ | Marginal R^2^ | Conditional R^2^ | AIC | σ^2^ | log-Likelihood |
| Full | 0.64 | 0.64 | 96457.05 | 12.07 | -48215.53 |
| Mean | 0.66 | 0.66 | 96619.27 | 12.18 | -48296.64 |
| BRIA | 0.53 | 0.54 | 86959.91 | 12.10 | -43466.95 |
| DKI | 0.54 | 0.54 | 86908.39 | 12.06 | -43441.20 |
| DTI | 0.50 | 0.50 | 87698.04 | 12.67 | -43836.02 |
| SMT | 0.54 | 0.55 | 87893.91 | 12.82 | -43933.95 |
| mcSMT | 0.55 | 0.56 | 87209.40 | 12.29 | -43591.70 |
| WMTI | 0.50 | 0.50 | 87648.19 | 12.63 | -43811.09 |
| **Health and lifestyle model** | | | | | |
| Approach^§^ | Marginal R^2^ | Conditional R^2^ | AIC | σ^2^ | log-Likelihood |
| Full | 0.64 | 0.65 | 88833.15 | 11.89 | -44398.58 |
| Mean | 0.65 | 0.65 | 89475.66 | 12.35 | -44719.83 |
| BRIA | 0.54 | 0.55 | 80358.89 | 11.97 | -40161.44 |
| DKI | 0.53 | 0.54 | 80570.27 | 12.14 | -40267.13 |
| DTI | 0.50 | 0.50 | 81152.24 | 12.62 | -40558.12 |
| SMT | 0.55 | 0.55 | 81237.26 | 12.69 | -40600.63 |
| mcSMT | 0.56 | 0.56 | 80666.80 | 12.22 | -40315.40 |
| WMTI | 0.50 | 0.50 | 81053.74 | 12.54 | -40508.87 |

**Supplementary Table 8.** Comparisons of model metrics of linear mixed effect models explaining brain age from all bio-psycho-social variables and reduced models

| **R^2^_C_** | **AIC** | **σ^2^** | **logLik** | **Diffusion Model** | **Statistical Model** |
| --- | --- | --- | --- | --- | --- |
| -0.0016 | 88.0922 | 0.0555 | -45.0461 | FULL | WHR Reduced |
| -0.0017 | 57.3767 | 0.0347 | -29.6883 | MEAN | WHR Reduced |
| -0.0039 | 107.2195 | 0.0821 | -54.6098 | BRIA | WHR Reduced |
| -0.0022 | 69.1367 | 0.0515 | -35.5683 | DKI | WHR Reduced |
| -0.0021 | 61.3670 | 0.0468 | -31.6835 | DTI | WHR Reduced |
| -0.0033 | 84.9249 | 0.0676 | -43.4624 | SMT | WHR Reduced |
| -0.0034 | 89.6927 | 0.0694 | -45.8463 | SMT_mc | WHR Reduced |
| -0.0026 | 76.1471 | 0.0594 | -39.0736 | WMTI | WHR Reduced |
| -0.0012 | 55.6408 | 0.0425 | -28.8204 | FULL | Hypertension Reduced |
| -0.0015 | 36.5504 | 0.0300 | -19.2752 | MEAN | Hypertension Reduced |
| -0.0018 | 53.4141 | 0.0456 | -27.7071 | BRIA | Hypertension Reduced |
| -0.0007 | 18.5097 | 0.0180 | -10.2548 | DKI | Hypertension Reduced |
| -0.0015 | 45.3233 | 0.0411 | -23.6617 | DTI | Hypertension Reduced |
| -0.0018 | 51.7974 | 0.0469 | -26.8987 | SMT | Hypertension Reduced |
| -0.0016 | 44.4942 | 0.0393 | -23.2471 | SMT_mc | Hypertension Reduced |
| -0.0014 | 42.3575 | 0.0384 | -22.1787 | WMTI | Hypertension Reduced |
| -0.0002 | 66.4775 | 0.0121 | -34.2387 | FULL | Self-Rated Health Reduced |
| -0.0001 | 59.5867 | 0.0075 | -30.7934 | MEAN | Self-Rated Health Reduced |
| -0.0010 | 84.9284 | 0.0272 | -43.4642 | BRIA | Self-Rated Health Reduced |
| 0.0001 | 51.0026 | 0.0019 | -26.5013 | DKI | Self-Rated Health Reduced |
| -0.0003 | 70.9981 | 0.0173 | -36.4990 | DTI | Self-Rated Health Reduced |
| -0.0005 | 70.2562 | 0.0168 | -36.1281 | SMT | Self-Rated Health Reduced |
| -0.0006 | 72.2845 | 0.0180 | -37.1422 | SMT_mc | Self-Rated Health Reduced |
| -0.0005 | 71.6833 | 0.0177 | -36.8416 | WMTI | Self-Rated Health Reduced |
| 0.0000 | 141.6590 | 0.0056 | -71.8295 | FULL | Happiness Reduced |
| 0.0000 | 136.8463 | 0.0022 | -69.4232 | MEAN | Happiness Reduced |
| 0.0000 | 106.3310 | -0.0044 | -54.1655 | BRIA | Happiness Reduced |
| 0.0001 | 111.5459 | -0.0004 | -56.7730 | DKI | Happiness Reduced |
| -0.0001 | 114.4043 | 0.0009 | -58.2021 | DTI | Happiness Reduced |
| 0.0000 | 112.6028 | -0.0007 | -57.3014 | SMT | Happiness Reduced |
| -0.0001 | 110.0182 | -0.0019 | -56.0091 | SMT_mc | Happiness Reduced |
| 0.0000 | 112.7427 | -0.0003 | -57.3713 | WMTI | Happiness Reduced |
| -0.0002 | 23.1137 | 0.0084 | -12.5569 | FULL | Matrix Puzzles Reduced |
| 0.0000 | 14.2695 | 0.0002 | -8.1348 | MEAN | Matrix Puzzles Reduced |
| -0.0001 | 15.9629 | 0.0019 | -8.9814 | BRIA | Matrix Puzzles Reduced |
| 0.0001 | 11.9709 | -0.0022 | -6.9855 | DKI | Matrix Puzzles Reduced |
| -0.0001 | 18.2088 | 0.0041 | -10.1044 | DTI | Matrix Puzzles Reduced |
| 0.0001 | 11.9069 | -0.0026 | -6.9534 | SMT | Matrix Puzzles Reduced |
| -0.0001 | 16.4788 | 0.0024 | -9.2394 | SMT_mc | Matrix Puzzles Reduced |
| -0.0002 | 19.8911 | 0.0061 | -10.9455 | WMTI | Matrix Puzzles Reduced |

R^2^_C_ refers to the difference in conditional variance explained (not including variance exlpained by the random factor) between full and reduced models.

**Supplementary Table 9.** Statistical model Comparison of linear mixed effect models explaining brain age from all bio-psycho-social variables and reduced models

| **AIC** | **BIC** | **logLik** | **Deviance** | **χ^2^** | ***p*-value** | **Regression Model** | **Diffusion Model** |
| --- | --- | --- | --- | --- | --- | --- | --- |
| 88838.61 | 88969.90 | -44402.30 | 88804.61 |  |  | FULL | Full Health Model |
| 88759.79 | 88898.81 | -44361.90 | 88723.79 | 80.82 | 0.00 | FULL | WHR Reduced |
| 89448.50 | 89579.80 | -44707.25 | 89414.50 |  |  | MEAN | Full Health Model |
| 89401.82 | 89540.83 | -44682.91 | 89365.82 | 48.69 | 0.00 | MEAN | WHR Reduced |
| 80388.81 | 80518.38 | -40177.41 | 80354.81 |  |  | BRIA | Full Health Model |
| 80286.51 | 80423.69 | -40125.25 | 80250.51 | 104.30 | 0.00 | BRIA | WHR Reduced |
| 80561.92 | 80691.48 | -40263.96 | 80527.92 |  |  | DKI | Full Health Model |
| 80497.53 | 80634.71 | -40230.76 | 80461.53 | 66.39 | 0.00 | DKI | WHR Reduced |
| 81134.46 | 81264.02 | -40550.23 | 81100.46 |  |  | DTI | Full Health Model |
| 81078.15 | 81215.33 | -40521.07 | 81042.15 | 58.31 | 0.00 | DTI | WHR Reduced |
| 81245.90 | 81375.46 | -40605.95 | 81211.90 |  |  | SMT | Full Health Model |
| 81165.77 | 81302.96 | -40564.89 | 81129.77 | 82.13 | 0.00 | SMT | WHR Reduced |
| 80678.92 | 80808.48 | -40322.46 | 80644.92 |  |  | SMT_mc | Full Health Model |
| 80594.30 | 80731.48 | -40279.15 | 80558.30 | 86.62 | 0.00 | SMT_mc | WHR Reduced |
| 81053.11 | 81182.68 | -40509.56 | 81019.11 |  |  | WMTI | Full Health Model |
| 80981.83 | 81119.01 | -40472.91 | 80945.83 | 73.29 | 0.00 | WMTI | WHR Reduced |
| 88818.50 | 88949.79 | -44392.25 | 88784.50 |  |  | FULL | Full Health Model |
| 88759.79 | 88898.81 | -44361.90 | 88723.79 | 60.71 | 0.00 | FULL | Hypertension Reduced |
| 89441.37 | 89572.66 | -44703.69 | 89407.37 |  |  | MEAN | Full Health Model |
| 89401.82 | 89540.83 | -44682.91 | 89365.82 | 41.55 | 0.00 | MEAN | Hypertension Reduced |
| 80342.82 | 80472.39 | -40154.41 | 80308.82 |  |  | BRIA | Full Health Model |
| 80286.51 | 80423.69 | -40125.25 | 80250.51 | 58.31 | 0.00 | BRIA | Hypertension Reduced |
| 80518.92 | 80648.49 | -40242.46 | 80484.92 |  |  | DKI | Full Health Model |
| 80497.53 | 80634.71 | -40230.76 | 80461.53 | 23.39 | 0.00 | DKI | Hypertension Reduced |
| 81126.41 | 81255.97 | -40546.20 | 81092.41 |  |  | DTI | Full Health Model |
| 81078.15 | 81215.33 | -40521.07 | 81042.15 | 50.26 | 0.00 | DTI | Hypertension Reduced |
| 81220.43 | 81349.99 | -40593.22 | 81186.43 |  |  | SMT | Full Health Model |
| 81165.77 | 81302.96 | -40564.89 | 81129.77 | 56.66 | 0.00 | SMT | Hypertension Reduced |
| 80641.66 | 80771.23 | -40303.83 | 80607.66 |  |  | SMT_mc | Full Health Model |
| 80594.30 | 80731.48 | -40279.15 | 80558.30 | 49.37 | 0.00 | SMT_mc | Hypertension Reduced |
| 81027.05 | 81156.61 | -40496.52 | 80993.05 |  |  | WMTI | Full Health Model |
| 80981.83 | 81119.01 | -40472.91 | 80945.83 | 47.22 | 0.00 | WMTI | Hypertension Reduced |
| 96416.23 | 96509.86 | -48196.11 | 96392.23 |  |  | FULL | Full Health Model |
| 96401.54 | 96502.98 | -48187.77 | 96375.54 | 16.69 | 0.00 | FULL | Self-Rated Health Reduced |
| 96570.51 | 96664.14 | -48273.25 | 96546.51 |  |  | MEAN | Full Health Model |
| 96562.99 | 96664.43 | -48268.49 | 96536.99 | 9.52 | 0.00 | MEAN | Self-Rated Health Reduced |
| 86937.52 | 87029.91 | -43456.76 | 86913.52 |  |  | BRIA | Full Health Model |
| 86904.90 | 87004.98 | -43439.45 | 86878.90 | 34.62 | 0.00 | BRIA | Self-Rated Health Reduced |
| 86856.20 | 86948.59 | -43416.10 | 86832.20 |  |  | DKI | Full Health Model |
| 86852.97 | 86953.05 | -43413.49 | 86826.97 | 5.23 | 0.02 | DKI | Self-Rated Health Reduced |
| 87657.83 | 87750.22 | -43816.92 | 87633.83 |  |  | DTI | Full Health Model |
| 87641.39 | 87741.48 | -43807.70 | 87615.39 | 18.44 | 0.00 | DTI | Self-Rated Health Reduced |
| 87859.55 | 87951.94 | -43917.78 | 87835.55 |  |  | SMT | Full Health Model |
| 87839.38 | 87939.46 | -43906.69 | 87813.38 | 22.17 | 0.00 | SMT | Self-Rated Health Reduced |
| 87175.47 | 87267.85 | -43575.74 | 87151.47 |  |  | SMT_mc | Full Health Model |
| 87154.02 | 87254.11 | -43564.01 | 87128.02 | 23.45 | 0.00 | SMT_mc | Self-Rated Health Reduced |
| 87613.39 | 87705.78 | -43794.70 | 87589.39 |  |  | WMTI | Full Health Model |
| 87593.37 | 87693.45 | -43783.68 | 87567.37 | 22.02 | 0.00 | WMTI | Self-Rated Health Reduced |
| 96400.94 | 96494.58 | -48188.47 | 96376.94 |  |  | FULL | Full Life Satisfaction Model |
| 96401.54 | 96502.98 | -48187.77 | 96375.54 | 1.41 | 0.24 | FULL | Happiness Reduced |
| 96561.03 | 96654.66 | -48268.51 | 96537.03 |  |  | MEAN | Full Life Satisfaction Model |
| 96562.99 | 96664.43 | -48268.49 | 96536.99 | 0.04 | 0.84 | MEAN | Happiness Reduced |
| 86903.40 | 86995.79 | -43439.70 | 86879.40 |  |  | BRIA | Full Life Satisfaction Model |
| 86904.90 | 87004.98 | -43439.45 | 86878.90 | 0.50 | 0.48 | BRIA | Happiness Reduced |
| 86851.19 | 86943.57 | -43413.59 | 86827.19 |  |  | DKI | Full Life Satisfaction Model |
| 86852.97 | 86953.05 | -43413.49 | 86826.97 | 0.22 | 0.64 | DKI | Happiness Reduced |
| 87639.77 | 87732.16 | -43807.89 | 87615.77 |  |  | DTI | Full Life Satisfaction Model |
| 87641.39 | 87741.48 | -43807.70 | 87615.39 | 0.38 | 0.54 | DTI | Happiness Reduced |
| 87837.40 | 87929.78 | -43906.70 | 87813.40 |  |  | SMT | Full Life Satisfaction Model |
| 87839.38 | 87939.46 | -43906.69 | 87813.38 | 0.01 | 0.91 | SMT | Happiness Reduced |
| 87152.34 | 87244.72 | -43564.17 | 87128.34 |  |  | SMT_mc | Full Life Satisfaction Model |
| 87154.02 | 87254.11 | -43564.01 | 87128.02 | 0.31 | 0.58 | SMT_mc | Happiness Reduced |
| 87591.97 | 87684.35 | -43783.98 | 87567.97 |  |  | WMTI | Full Life Satisfaction Model |
| 87593.37 | 87693.45 | -43783.68 | 87567.37 | 0.60 | 0.44 | WMTI | Happiness Reduced |
| 69855.96 | 69945.75 | -34915.98 | 69831.96 |  |  | FULL | Full Life Satisfaction Model |
| 69848.20 | 69945.48 | -34911.10 | 69822.20 | 9.76 | 0.00 | FULL | Matrix Puzzles Reduced |
| 69770.21 | 69860.00 | -34873.10 | 69746.21 |  |  | MEAN | Full Cognitive Model |
| 69770.47 | 69867.75 | -34872.24 | 69744.47 | 1.74 | 0.19 | MEAN | Matrix Puzzles Reduced |
| 63172.76 | 63261.30 | -31574.38 | 63148.76 |  |  | BRIA | Full Cognitive Model |
| 63170.87 | 63266.80 | -31572.44 | 63144.87 | 3.89 | 0.05 | BRIA | Matrix Puzzles Reduced |
| 63125.15 | 63213.70 | -31550.58 | 63101.15 |  |  | DKI | Full Cognitive Model |
| 63125.22 | 63221.15 | -31549.61 | 63099.22 | 1.93 | 0.16 | DKI | Matrix Puzzles Reduced |
| 63580.08 | 63668.63 | -31778.04 | 63556.08 |  |  | DTI | Full Cognitive Model |
| 63575.08 | 63671.00 | -31774.54 | 63549.08 | 7.00 | 0.01 | DTI | Matrix Puzzles Reduced |
| 63713.21 | 63801.76 | -31844.61 | 63689.21 |  |  | SMT | Full Cognitive Model |
| 63714.37 | 63810.30 | -31844.18 | 63688.37 | 0.84 | 0.36 | SMT | Matrix Puzzles Reduced |
| 63584.87 | 63673.42 | -31780.43 | 63560.87 |  |  | SMT_mc | Full Cognitive Model |
| 63583.77 | 63679.70 | -31778.89 | 63557.77 | 3.10 | 0.08 | SMT_mc | Matrix Puzzles Reduced |
| 63519.69 | 63608.23 | -31747.84 | 63495.69 |  |  | WMTI | Full Cognitive Model |
| 63514.76 | 63610.68 | -31744.38 | 63488.76 | 6.93 | 0.01 | WMTI | Matrix Puzzles Reduced |

**Supplementary Table 10.** Model metrics for linear mixed effect models explaining brain age by sex

| **FEMALES** | | | | | | **MALES** | | | | | |
| --- | --- | --- | --- | --- | --- | --- | --- | --- | --- | --- | --- |
| **Baseline model** | | | | | | **Baseline model** | | | | | |
| Approach | Marginal R^2^ | Conditional R^2^ | AIC | σ^2^ | log-Likelihood | Approach | Marginal R^2^ | Conditional R^2^ | AIC | σ^2^ | log-Likelihood |
| Full | 0.63 | 0.64 | 90953.56 | 12.26 | -45472.78 | Full | 0.65 | 0.66 | 80735 | 12.01 | -40363.5 |
| Mean | 0.63 | 0.63 | 91786.45 | 12.88 | -45889.23 | Mean | 0.66 | 0.66 | 81039.21 | 12.25 | -40515.61 |
| BRIA | 0.51 | 0.51 | 82506.57 | 12.61 | -41249.29 | BRIA | 0.53 | 0.53 | 72763.64 | 12.17 | -36377.82 |
| DKI | 0.5 | 0.5 | 82577.64 | 12.67 | -41284.82 | DKI | 0.54 | 0.54 | 72261.14 | 11.72 | -36126.57 |
| DTI | 0.47 | 0.47 | 83289.84 | 13.28 | -41640.92 | DTI | 0.51 | 0.51 | 73131.95 | 12.5 | -36561.97 |
| SMT | 0.51 | 0.51 | 83692.58 | 13.62 | -41842.29 | SMT | 0.55 | 0.56 | 73028.83 | 12.41 | -36510.41 |
| mcSMT | 0.52 | 0.53 | 82999.36 | 13.02 | -41495.68 | mcSMT | 0.56 | 0.56 | 72721.97 | 12.13 | -36356.99 |
| WMTI | 0.47 | 0.47 | 83047.24 | 13.06 | -41519.62 | WMTI | 0.5 | 0.5 | 73132.36 | 12.5 | -36562.18 |
| **Social factors model** | | | | | | **Social factors model** | | | | | |
| Approach | Marginal R^2^ | Conditional R^2^ | AIC | σ^2^ | log-Likelihood | Approach | Marginal R^2^ | Conditional R^2^ | AIC | σ^2^ | log-Likelihood |
| Full | 0.63 | 0.63 | 79642.98 | 12.24 | -39813.49 | Full | 0.65 | 0.66 | 75922.85 | 11.98 | -37953.42 |
| Mean | 0.63 | 0.63 | 80367.28 | 12.85 | -40175.64 | Mean | 0.66 | 0.66 | 76205.03 | 12.23 | -38094.52 |
| BRIA | 0.51 | 0.51 | 72319.9 | 12.6 | -36151.95 | BRIA | 0.53 | 0.53 | 68516.13 | 12.14 | -34250.06 |
| DKI | 0.5 | 0.51 | 72363.19 | 12.64 | -36173.6 | DKI | 0.54 | 0.54 | 68048.63 | 11.7 | -34016.32 |
| DTI | 0.47 | 0.47 | 73032.7 | 13.29 | -36508.35 | DTI | 0.51 | 0.51 | 68851.96 | 12.46 | -34417.98 |
| SMT | 0.51 | 0.52 | 73342.83 | 13.6 | -36663.41 | SMT | 0.55 | 0.56 | 68752.23 | 12.36 | -34368.12 |
| mcSMT | 0.53 | 0.53 | 72714.11 | 12.98 | -36349.06 | mcSMT | 0.56 | 0.56 | 68466.31 | 12.09 | -34225.16 |
| WMTI | 0.47 | 0.47 | 72754.14 | 13.02 | -36369.07 | WMTI | 0.5 | 0.5 | 68845.68 | 12.45 | -34414.84 |
| **Cognitive factors model** | | | | | | **Cognitive factors model** | | | | | |
| Approach | Marginal R^2^ | Conditional R^2^ | AIC | σ^2^ | log-Likelihood | Approach | Marginal R^2^ | Conditional R^2^ | AIC | σ^2^ | log-Likelihood |
| Full | 0.63 | 0.63 | 37254.37 | 11.9 | -18616.19 | Full | 0.66 | 0.66 | 32698.63 | 11.93 | -16338.32 |
| Mean | 0.63 | 0.63 | 37231.66 | 11.87 | -18604.83 | Mean | 0.67 | 0.67 | 32645.73 | 11.83 | -16311.86 |
| BRIA | 0.49 | 0.5 | 33684.02 | 12.03 | -16831.01 | BRIA | 0.51 | 0.52 | 29591.07 | 12.27 | -14784.54 |
| DKI | 0.49 | 0.49 | 33882.38 | 12.43 | -16930.19 | DKI | 0.53 | 0.53 | 29337.22 | 11.72 | -14657.61 |
| DTI | 0.45 | 0.45 | 34073.61 | 12.8 | -17025.81 | DTI | 0.51 | 0.51 | 29605.97 | 12.31 | -14791.99 |
| SMT | 0.5 | 0.51 | 34154.38 | 12.97 | -17066.19 | SMT | 0.54 | 0.55 | 29660.38 | 12.43 | -14819.19 |
| mcSMT | 0.5 | 0.51 | 34065.24 | 12.78 | -17021.62 | mcSMT | 0.54 | 0.55 | 29622.69 | 12.34 | -14800.35 |
| WMTI | 0.45 | 0.46 | 33924.27 | 12.5 | -16951.14 | WMTI | 0.5 | 0.5 | 29697.03 | 12.51 | -14837.52 |
| **Wellbeing / satisfaction model** | | | | | | **Wellbeing / satisfaction model** | | | | | |
| Approach | Marginal R^2^ | Conditional R^2^ | AIC | σ^2^ | log-Likelihood | Approach | Marginal R^2^ | Conditional R^2^ | AIC | σ^2^ | log-Likelihood |
| Full | 0.61 | 0.62 | 50349.36 | 11.95 | -25163.68 | Full | 0.65 | 0.65 | 46144.54 | 12.19 | -23061.27 |
| Mean | 0.64 | 0.64 | 50559.78 | 12.22 | -25268.89 | Mean | 0.67 | 0.67 | 46096.21 | 12.13 | -23037.1 |
| BRIA | 0.53 | 0.53 | 45586.52 | 11.98 | -22782.26 | BRIA | 0.54 | 0.54 | 41404.97 | 12.23 | -20691.49 |
| DKI | 0.52 | 0.52 | 45683.58 | 12.12 | -22830.79 | DKI | 0.55 | 0.55 | 41258.41 | 12 | -20618.21 |
| DTI | 0.48 | 0.48 | 46100.11 | 12.73 | -23039.05 | DTI | 0.52 | 0.52 | 41634.01 | 12.61 | -20806.01 |
| SMT | 0.52 | 0.53 | 46361.38 | 13.11 | -23169.69 | SMT | 0.56 | 0.57 | 41561.6 | 12.48 | -20769.8 |
| mcSMT | 0.54 | 0.55 | 45865.72 | 12.38 | -22921.86 | mcSMT | 0.57 | 0.57 | 41374.43 | 12.19 | -20676.22 |
| WMTI | 0.48 | 0.48 | 46042.74 | 12.64 | -23010.37 | WMTI | 0.51 | 0.51 | 41645.05 | 12.62 | -20811.53 |
| **Health risk model** | | | | | | **Health risk model** | | | | | |
| Approach | Marginal R^2^ | Conditional R^2^ | AIC | σ^2^ | log-Likelihood | Approach | Marginal R^2^ | Conditional R^2^ | AIC | σ^2^ | log-Likelihood |
| Full | 0.63 | 0.63 | 52562.3 | 11.92 | -26265.15 | Full | 0.65 | 0.66 | 36324.72 | 11.83 | -18146.36 |
| Mean | 0.63 | 0.64 | 53266.06 | 12.81 | -26617.03 | Mean | 0.67 | 0.68 | 36239.57 | 11.69 | -18103.78 |
| BRIA | 0.53 | 0.54 | 47815.34 | 12.19 | -23891.67 | BRIA | 0.55 | 0.55 | 32587.22 | 11.65 | -16277.61 |
| DKI | 0.52 | 0.52 | 47971.56 | 12.4 | -23969.78 | DKI | 0.54 | 0.54 | 32648.25 | 11.77 | -16308.12 |
| DTI | 0.49 | 0.49 | 48330.92 | 12.92 | -24149.46 | DTI | 0.52 | 0.52 | 32851.08 | 12.17 | -16409.54 |
| SMT | 0.53 | 0.53 | 48534.39 | 13.21 | -24251.19 | SMT | 0.57 | 0.57 | 32725.57 | 11.92 | -16346.78 |
| mcSMT | 0.54 | 0.55 | 48101.56 | 12.59 | -24034.78 | mcSMT | 0.57 | 0.58 | 32597.77 | 11.67 | -16282.89 |
| WMTI | 0.49 | 0.49 | 48221.35 | 12.75 | -24094.67 | WMTI | 0.51 | 0.51 | 32873.33 | 12.21 | -16420.66 |

#### 1.2 Supplementary Figures

**Supplementary Figure 1. Overview of brain age calculations and following modelling in bio-psycho-social models**

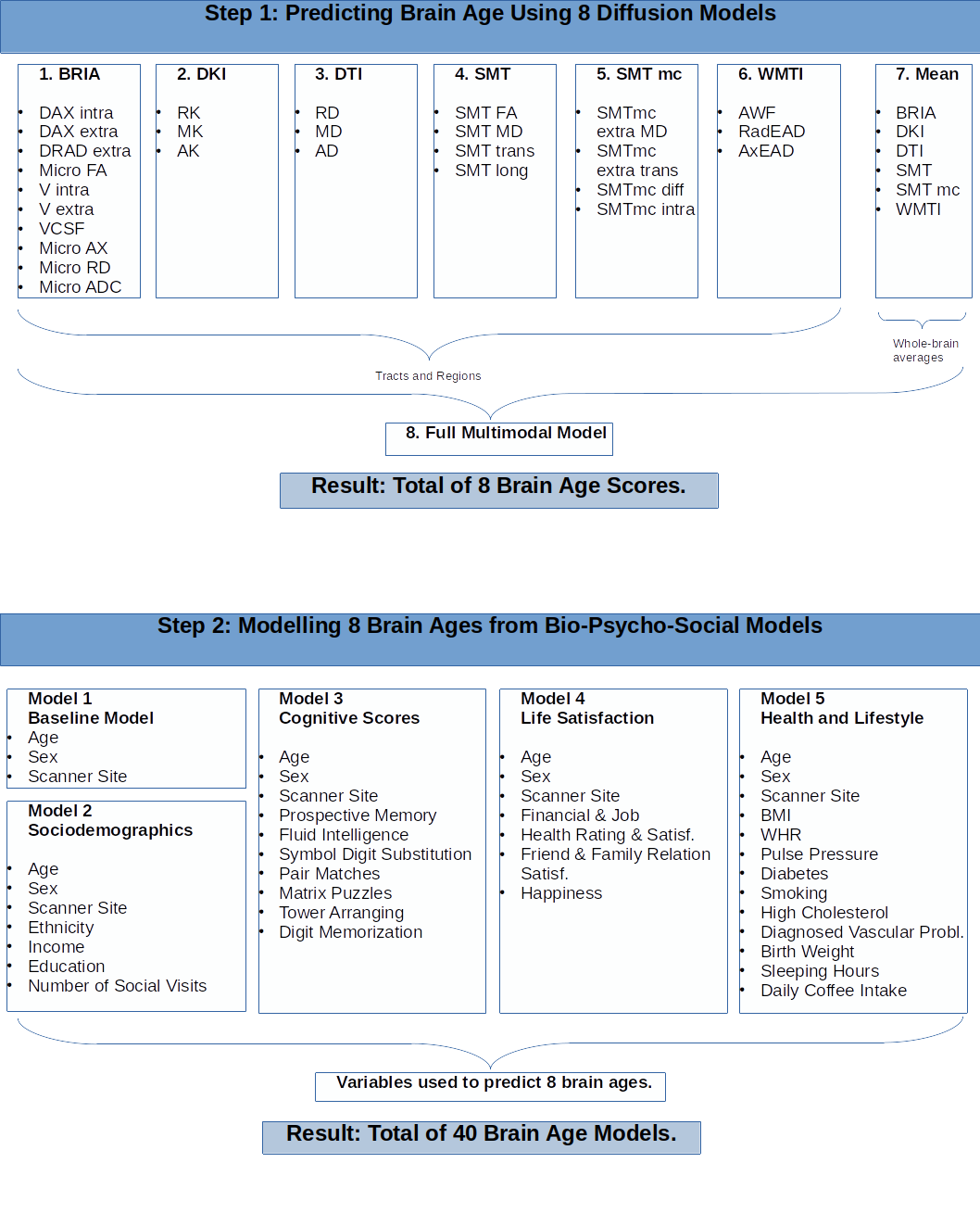

**Supplementary Figure 2.** Correlations of Cognitive Scores

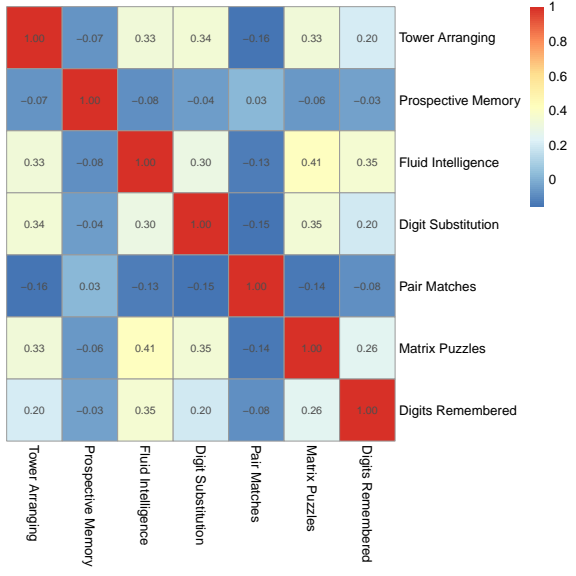

Variance inflation factor for sex VIF = 75.41, age VIF = 2.30, and the age-sex interaction VIF = 77.65. All other VIFs < 1.46 indicating small correlations.

**Supplementary Figure 3.** Correlations of Life Satisfaction Scores

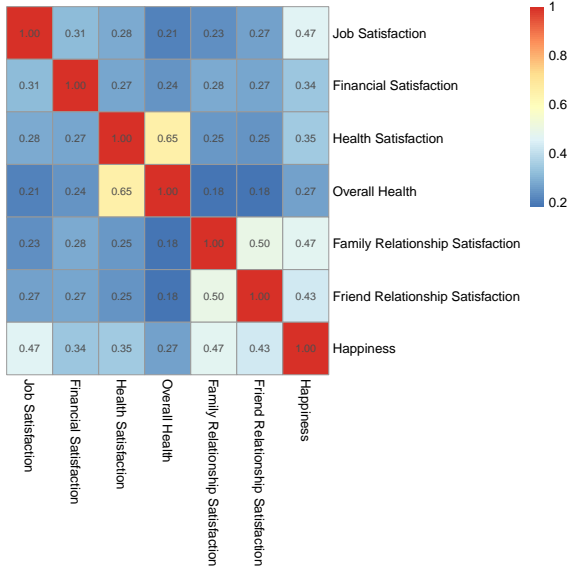

Variance inflation factor for sex VIF = 68.26, age VIF = 2.10, and the age-sex interaction VIF = 70.75. All other VIFs < 1.77 indicate small correlations.

**Supplementary Figure 4.** Correlations of Health Risk Scores

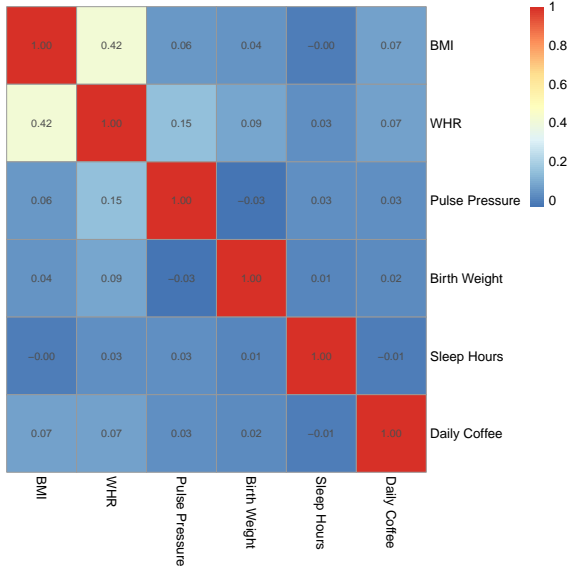
Variance inflation factor for sex VIF = 73.10, age VIF = 2.09, and the age-sex interaction VIF = 73.85. All other VIFs < 2.33 indicate small correlations with the WHR VIF = 2.32 being the strongest correlated feature (with BMI).

**Supplementary Figure 5.** Models’ Standardized *β-V*alues with Standard Deviation for Mixed Linear Models separating males and females

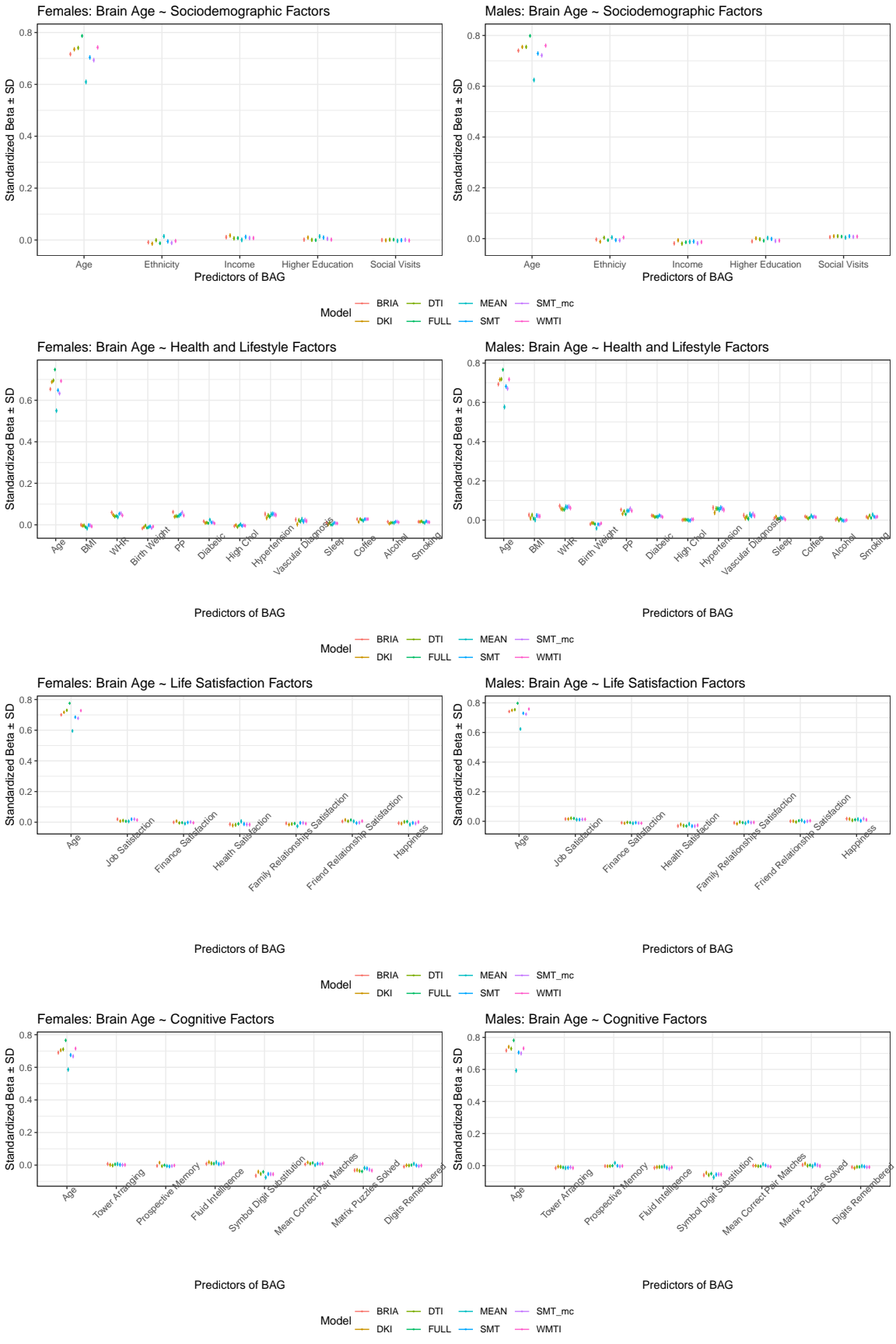

**Supplementary Figure 6.** Sociodemographic Model Predictors’ *Un*Standardized Beta-Values with 95% Confidence Interval

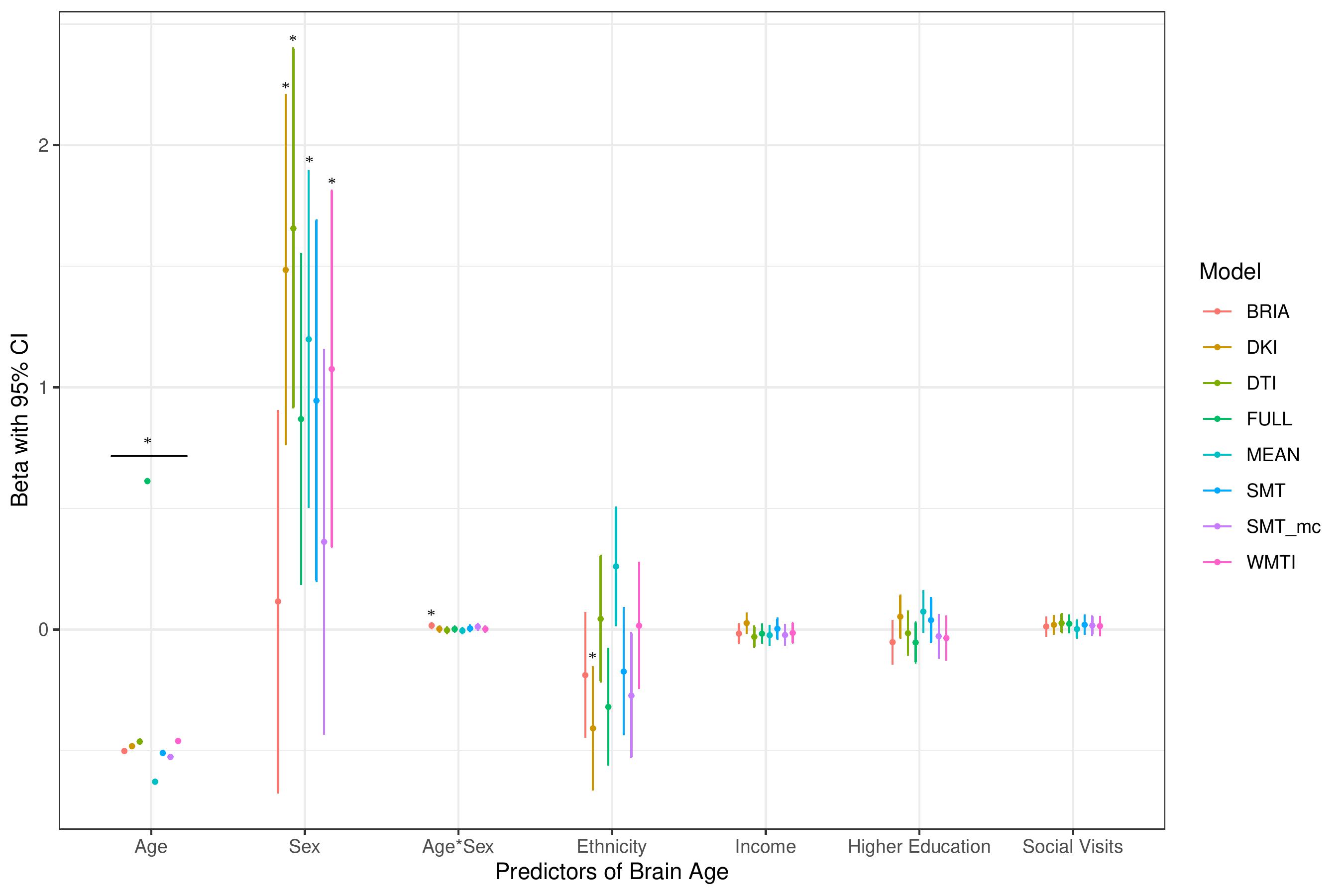

* indicates Bonferroni-corrected *p* < .05

**Supplementary Figure 7.** Health Model Predictors’ UnStandardized Beta-Values with 95% Confidence Interval

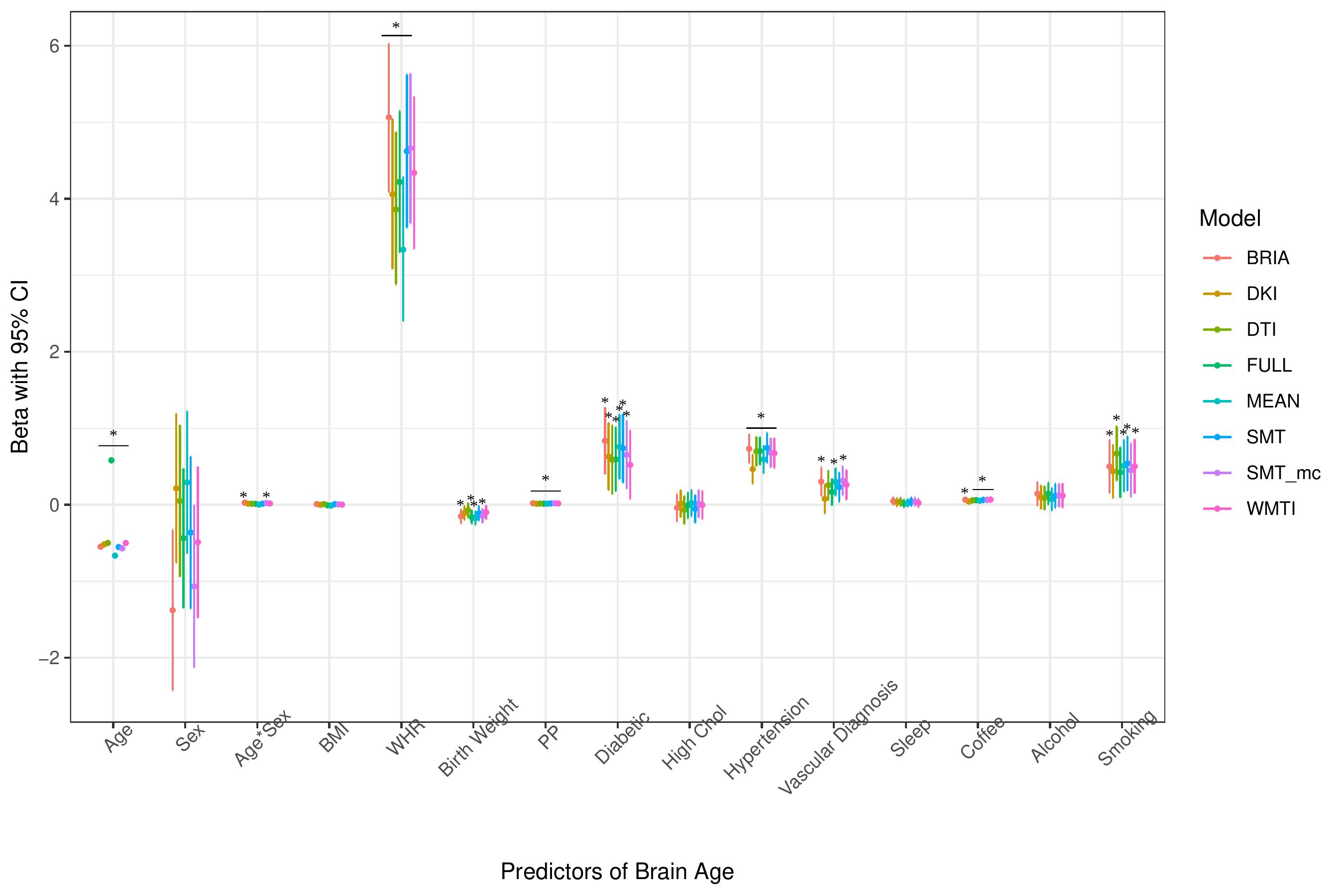
* indicates Bonferroni-corrected *p* < .05

**Supplementary Figure 8.** Well-being Model Predictors’ UnStandardized Beta-Values with 95% Confidence Interval

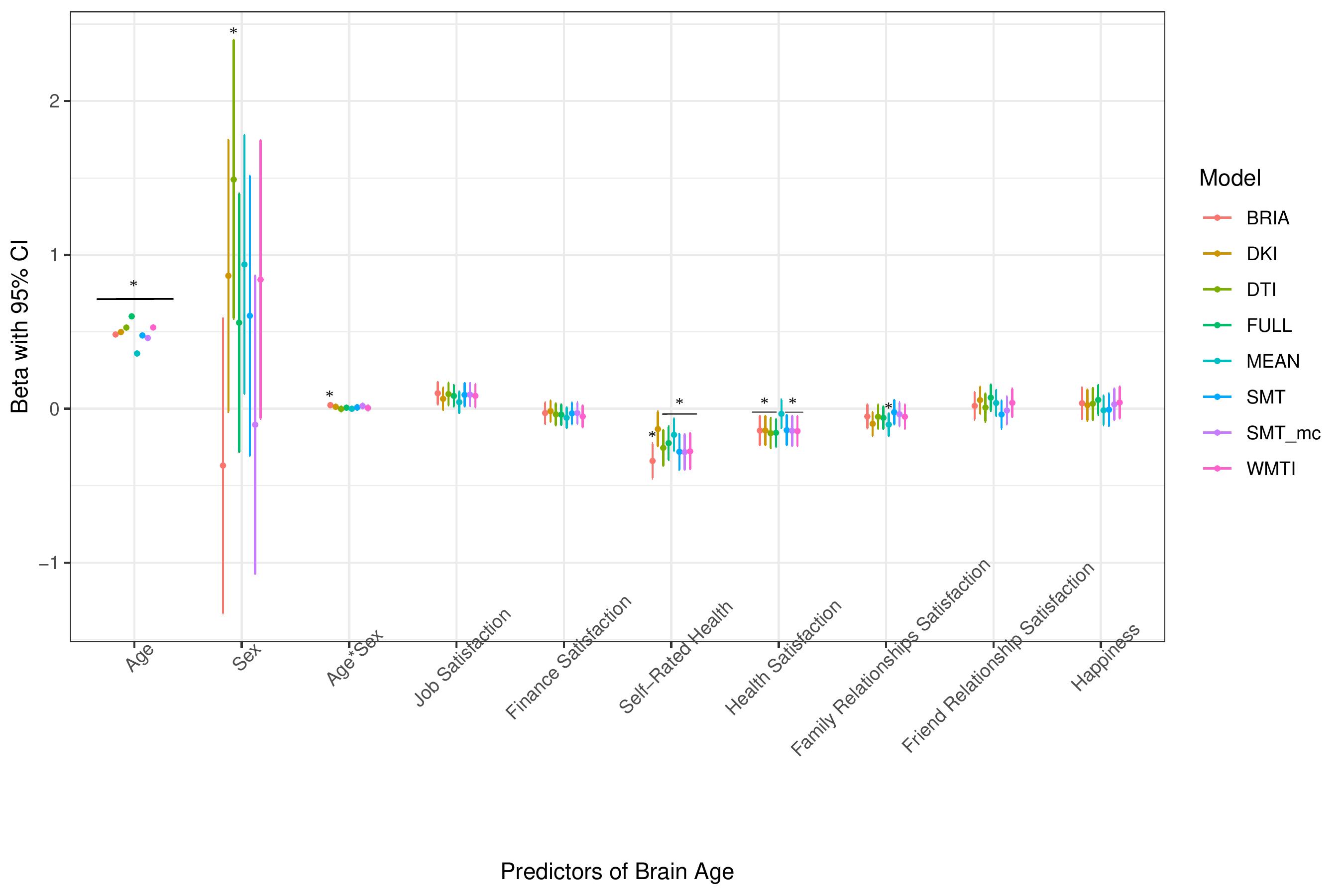
* indicates Bonferroni-corrected *p* < .05

**Supplementary Figure 9.** Cognition Model Predictors’ UnStandardized Beta-Values with 95% Confidence Interval

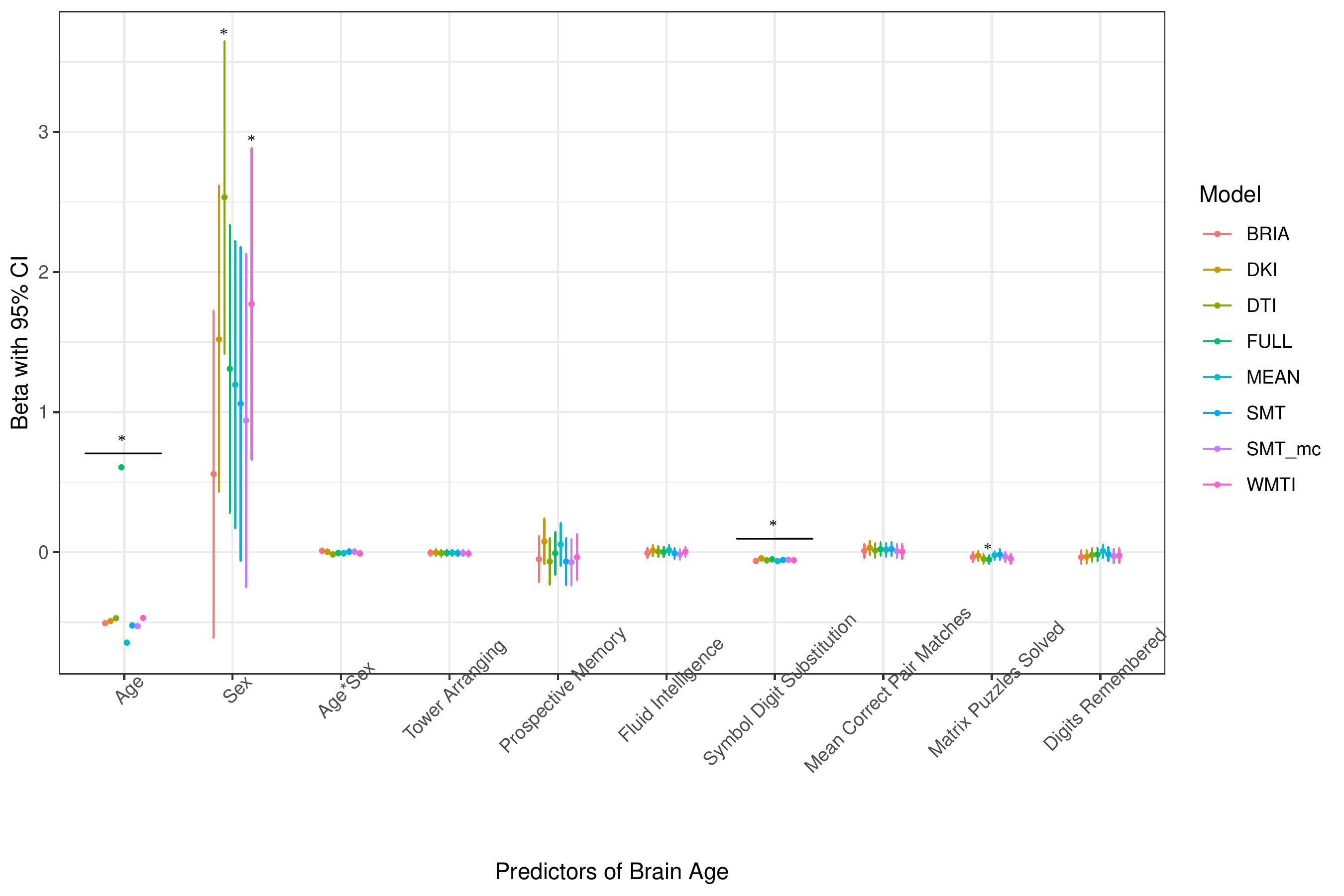
* indicates Bonferroni-corrected *p* < .05

**Supplementary Figure 10.** Health Model without WHR as predictor

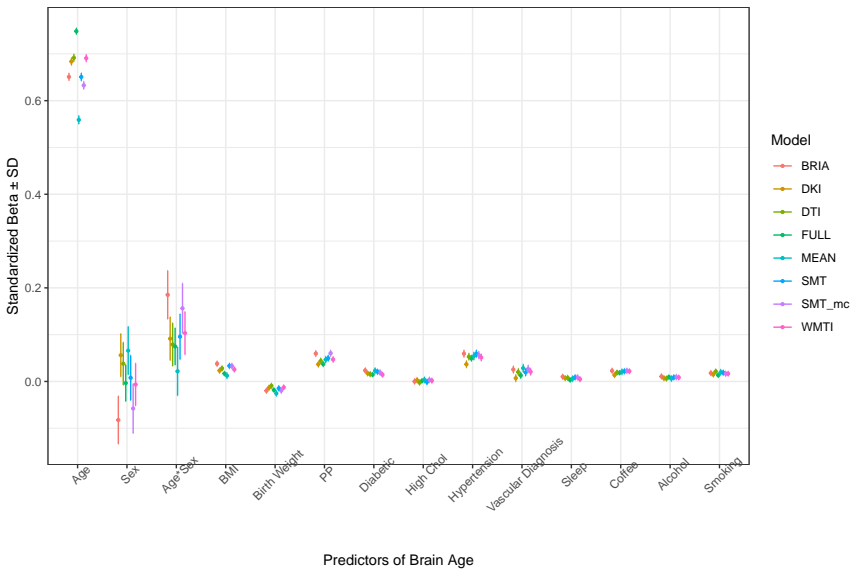

**Supplementary Figure 11.** Health Model without hypertension as predictor

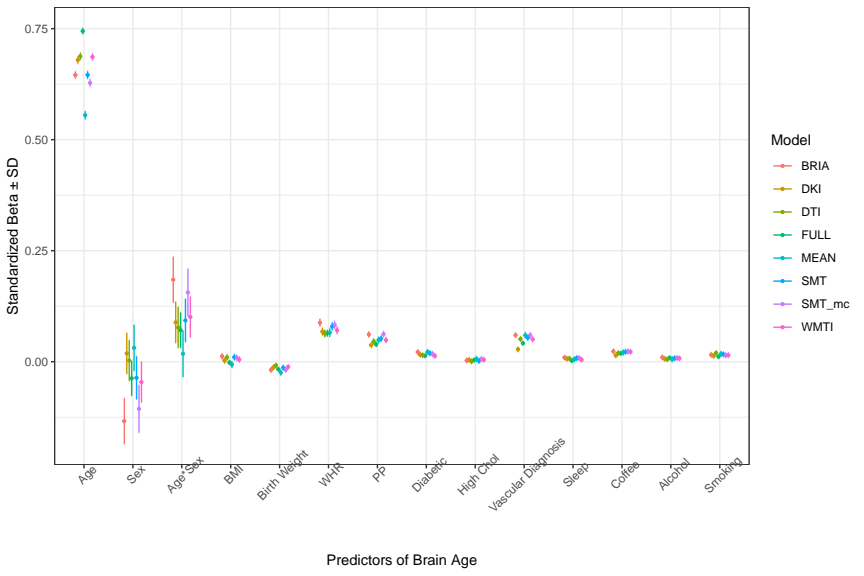

**Supplementary Figure 12.** Life-Satisfaction model without self-rated health satisfaction as predictor

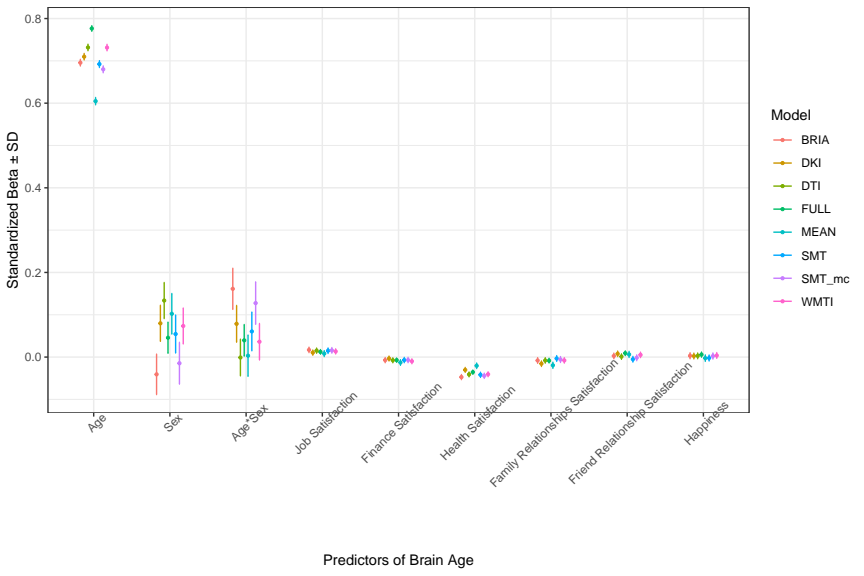

**Supplementary Figure 13.** Life satisfaction model without self-rated happiness as predictor

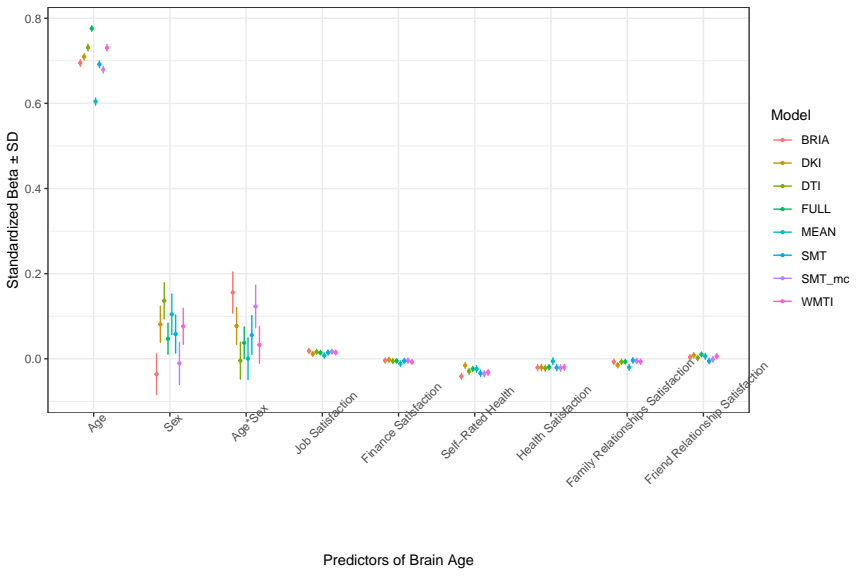

**Supplementary Figure 14.** Cognitive factor model without number of matrix puzzles as predictor

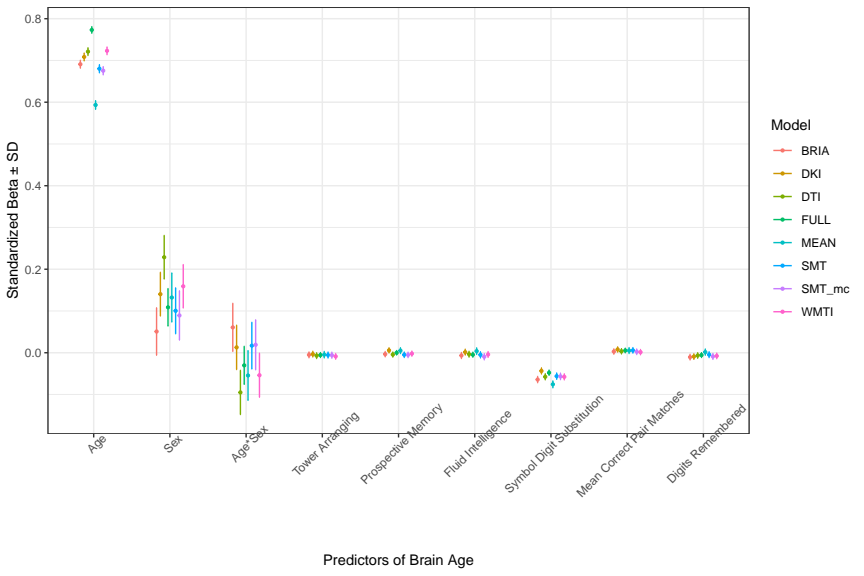
